# Non-Invasive, MultiOmic and MultiCompartmental Biomarkers of Reflux Disease: A Systematic Review

**DOI:** 10.1101/2022.06.20.22276215

**Authors:** Muhammad S. Farooqi, Sanjiti Podury, George Crowley, Sophia Kwon, Abraham R. Khan, Fritz Francois, Anna Nolan

## Abstract

**Background and Aims:** Gastroesophageal reflux disease (GERD) is a prevalent GI disorder which may complicate conditions such as obstructive airways disease (OAD). Our group has identified predictive biomarkers of GERD in particulate exposed 1^st^ responders with OAD. Additionally, GERD diagnosis and treatment is costly, and invasive. In light of these clinical concerns our aim was to systematically review studies identifying non-invasive, multiOmic and multi-compartmental biomarkers of GERD.

**Methods:** A systematic review of PubMed and EMBASE was performed on February 22, 2022 utilizing keywords focusing on reflux disease and biomarkers. The study was registered with PROSPERO (2022-CRD42022301543). We included: original human studies in English, published after December 31, 2009 focusing on non-invasive biomarkers of GERD. Reflux related conditions included Nonerosive Reflux Disease (NERD) Laryngopharyngeal Disease (LPR), Erosive Esophagitis (EE) and Barretts Esophagus (BE). Predictive measures were synthesized and bias assessed.

**Results:** Primary search identified 241 studies. After removing duplicates and applying inclusion/exclusion criteria n=15 articles were identified. Salivary pepsin was the most studied biomarker (n=5) with a significant sensitivity and specificity for GERD and LPR detection. Studies showed that for GERD diagnosis, miR-203 downregulation had the highest area under curve the receiver operator curve(ROC_AUC_) 0.94(95% CI; 0.90-0.7). An oral microbiome model including *Lautropia*, *Streptococcus* and *Bacteroidetes* showed the greatest discrimination between BE and controls vs *Lautropia* alone; sensitivity of 96.9%, specificity of 88.2% and ROC_AUC_ of 0.94(0.81-1.00).

**Conclusion:** Prior studies identified significant multiOmic, multi-compartmental non-invasive biomarker risks for GERD and its complications such as BE. However, due to study limitations and to further ascertain the reliability and accuracy of these biomarkers more studies are warranted.

**WHAT YOU NEED TO KNOW:** *BACKGROUND:* Gastroesophageal reflux disease (GERD) is a prevalent GI disorder which may complicate conditions such as obstructive airways disease (OAD). GERD diagnosis and treatment is costly, and invasive. In light of these clinical concerns our aim was to systematically review studies identifying non-invasive, multiOmic and multi-compartmental biomarkers of GERD.

*FINDINGS:* Salivary pepsin was the most studied biomarker with a significant sensitivity and specificity for GERD detection. Studies showed that for GERD diagnosis, miR-203 downregulation had the highest ROC_AUC_. An oral microbiome model including Lautropia, Streptococcus and Bacteroidetes showed the greatest discrimination between Barrette’s Esophagus and controls vs Lautropia alone.

*IMPLICATIONS FOR PATIENT CARE:* Prior studies identified significant multiOmic, multi-compartmental non-invasive biomarker risks for GERD and its complications such as BE. However, due to study limitations and to further ascertain the reliability, accuracy and clinical utility of these biomarkers more studies are warranted.

## BACKGROUND

Gastroesophageal reflux disease (GERD), is a highly prevalent disorder; has an incidence of 5/1000 person-years and costs >$9-10 billion/year.(1–5) GERD diminishes health-related quality of life (QoL), productivity, accounts for about 5% of outpatient visits and is an independent risk factor of the metaplastic changes of BE.(3, 6–8) Refractory reflux was associated with both anxiety and depression, and once weekly episodes of GERD was detrimental to QoL.(8–10) Extraesophageal manifestations, include laryngopharyngeal disease (LPR), may lead to complications such as erosive esophagitis (EE) and Barrett’s Esophagus (BE).(11–15) An estimated 15% of all visits to otolaryngologists are due to LPR.(16)

GERD can be diagnosed clinically as per the Montreal consensus or it can be defined using invasive testing as per the Porto and Lyon consensus statements.(17–21) Conclusive evidence of GERD includes endoscopic findings of erosive esophagitis Los Angeles(LA) grade C or D, a stricture or Barrett’s esophagus, as well as an esophageal acid exposure time >6% on a pH or pH impedance study in at least one full day of recording.(17–20) In the recently published American College of Gastroenterology (ACG) guidelines GERD is objectively defined by the presence of characteristic mucosal injury seen and/or an abnormal reflux monitoring study.(21)

**Diagnosing GERD may be challenging** and includes clinical symptoms of heartburn and/or regurgitation,(22, 23) endoscopy, response to PPI (24, 25), alarm symptoms, and reflux monitoring.(21) The main methods of reflux testing include a wireless telemetry capsule attached to the esophageal mucosa during endoscopy and trans-nasal catheter-based testing.(26, 27) Similarly, high resolution manometry (HRM) is recommended for patients that may need anti-reflux surgery.(28) These tests are invasive, and expensive. Several scoring systems have been developed such as the Reflux Finding Score (RFS) (29) for grading laryngoscopy findings. However, it has been found that the correlation between symptoms, laryngoscopic findings and other objective testing such as pH-impedance testing is low.(30, 31)

In addition, to these heterogeneous often invasive diagnostics there have been studies focused on pepsin and other less invasive biomarkers and associated reviews.(32, 33) Therefore, it is in the context of these prior works and clinical need that we developed our systematic review which focuses on non-invasive multicompartmental biomarkers of reflux disease and severity.

## METHODS

### Review Strategy

Our systematic review adhered to the Preferred Reporting Items for Systematic Reviews and Meta-analysis (PRISMA) guidelines.(34, 35) Our Population, Intervention, Control, Outcome **(PICO)** question was “In adult patients with diagnosed reflux disease including GERD, LPR, EE and BE **(P)**, we performed a systematic review to identify**(I)** the non-invasive multiOmic and multi-compartmental biomarkers of reflux disease in adult subjects **(O)**”. Given the design of our systematic review, no comparison control **(C)** was needed PubMed and EMBASE were searched on February 2, 2022 as per the protocol of our systematic review were registered on <u>PROSPERO</u>(2022-CRD42022301543) and can be accessed at https://www.crd.york.ac.uk/prospero/display_record.php?RecordID=301543

### Search Terms

Databases were searched for the following:

(Gastric Acid Reflux OR Gastric Acid Reflux Disease OR Gastro-Esophageal Reflux Disease OR Gastro Esophageal Reflux Disease OR Gastro-Esophageal Reflux Diseases OR Gastro-oesophageal Reflux OR Gastro oesophageal Reflux OR Gastroesophageal Reflux Disease OR GERD OR Reflux, Gastroesophageal OR Esophageal Reflux OR Gastro-Esophageal Reflux OR Gastro Esophageal Reflux) **AND** (Biological Marker OR Biologic Marker OR Biological Markers OR Biologic Markers OR Biomarker OR Immune Markers OR Immunologic Markers OR Immune Marker OR Immunologic Marker OR Serum Markers OR Serum Marker OR Surrogate Endpoints OR Surrogate End Point OR Surrogate End Points OR Surrogate Endpoint OR Clinical Markers OR Clinical Marker OR Viral Markers OR Viral Marker OR Biochemical Marker OR Biochemical Markers OR Laboratory Markers OR Laboratory Marker OR Surrogate Markers OR Surrogate Marker)

### Reference-list screening was also used

For this review, we have <u>defined</u>: reflux disease to include GERD and its presentations such as nonerosive reflux disease(NERD) with no evidence of mucosal injury, erosive reflux disease also refer to as erosive esophagitis (EE)(36), extraesophageal manifestations such as laryngopharyngeal disease (LPR) and complications such BE. Also, to best utilize the limited studies on non-invasive biomarkers we have not excluded studies which include subjects with high grade dysplasia and esophageal adenocarcinoma in the context of GERD and/or BE.

### Study Criteria

Studies were <u>included</u> if they were of: (**1**) non-invasive biomarkers of GERD in blood, serum or saliva or exhaled breath in diagnosed adult (clinically or endoscopically) reflux patients (**2**) Evaluated diagnostic tests (assessed sensitivity, specificity, positive/negative predictive values, risk and/or accuracy of these biomarkers). (**3**) were written in English and (**4**) published after December 31^st^, 2009.

Studies were excluded if they (**1**) were not original research; (**2**) not written in English; (**3**) published before January 1, 2010 (**4**) included any non-human subjects or in-vitro studies (4) were conducted in a pediatric population or (**5**) focused on biomarkers in biopsied specimens or involved invasive tissue sampling (**6**) immunohistochemistry.

### Data Extraction

Articles were reviewed and data regarding study design, patient characteristics, sample size, tools used, severity and prevalence of reflux disease was extracted. Results from each database search were filtered for human subjects, English language, and publication date and imported into (EndNote X9). The references were then screened for duplicates using RefWorks (ProQuest LLC). Original research papers were reviewed for (title, abstract and full text) to ascertain eligibility. We examined references cited in the relevant articles. All results were screened by **MSF** and **SP** and further independently evaluated by **AN**. Disagreements were resolved by consensus. Details as per 2020 Preferred Reporting Items for Systematic Reviews and Meta-Analyses (PRISMA) are found in **Figure 1** and resultant manuscripts that meet these criteria are detailed, **Supplemental Tables 1-6**.(37)

**Figure 1.**
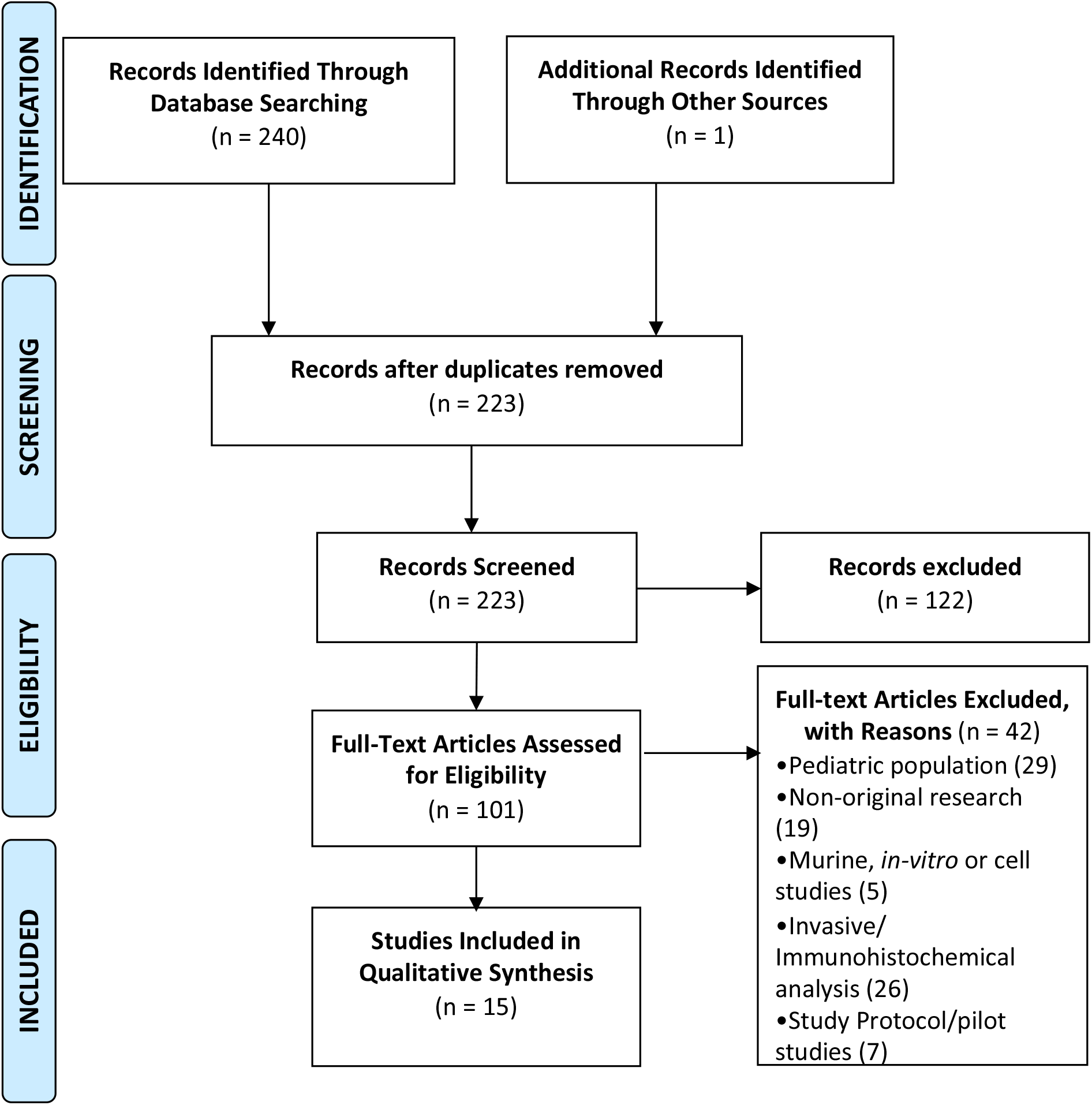
Study Design. Flow Diagram as per PRISMA Guidelines.(34, 35)

### Risk of Bias Assessment

Systematic reviews inherent biases (selection, detection, performance, and reporting) were addressed through study design/search algorithm. Selection bias was addressed by having pre-determined inclusion, exclusion criteria and distinct definitions. Detection and performance bias were addressed by having at least two rounds of screening individually performed by MF and SP. Reporting bias was minimized by using PubMed and EMBASE-search filters for peer-reviewed published articles of human subjects written in English and removing duplicates.

The Newcastle-Ottawa Scale (NOS)(38) a domain based approach was used to assess the degree of bias as in prior studies.(39) Low risk studies reflected were concordant in all domains (green); studies with at least one unclear or high-risk domain were considered as unclear or high risk of bias studies (yellow or red), respectively, **Supplemental Tables 7A-B.**

## RESULTS

### Literature Search

A total of 241 studies were identified from PubMed, EMBASE and reference-list screening, **Figure 1.** After application of selection criteria, 223 research papers were assessed for inclusion. Exclusion criteria met by n=122, **Supplemental Table 5**. Finally, n=15 original research articles were considered eligible. Data from screening and extraction are available, **Supplemental Tables 1-6**. (**40-54**)

Risk of bias using NOS was performed in case-cohort (n=6) and case control (n=9) studies. Case-cohort studies, n=3 had a high risk, n=1 had an unclear risk and n=2 had low risk of bias. Similarly for case-control studies n=8 had a high risk and n=1 had low risk of bias, **Supplemental Table 7A-B.**

### Study Characteristics

The populations of patients with reflux that were studied included: GERD (n=11), LPR (n=2), BE (n=7), EE (n=1) and EAC (n=2) while one study compared nonerosive reflux disease (NERD) vs erosive reflux disease (ERD). The biomarkers were found in biospecimens such as saliva (n=8), serum (n=4), exhaled breath (n=2) and exfoliated tongue cells (n=1), **Table 1.**

**Table 1.**
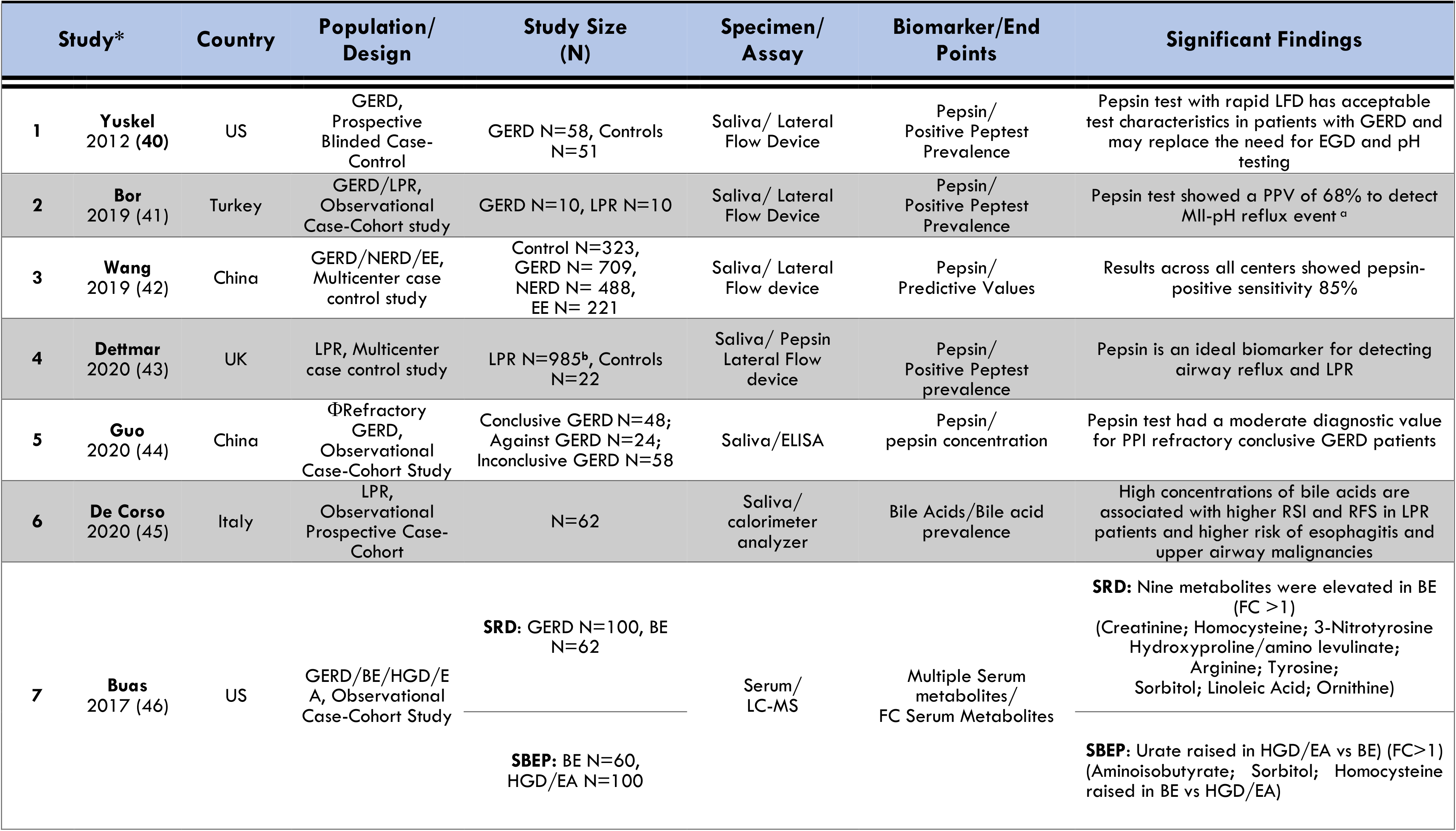
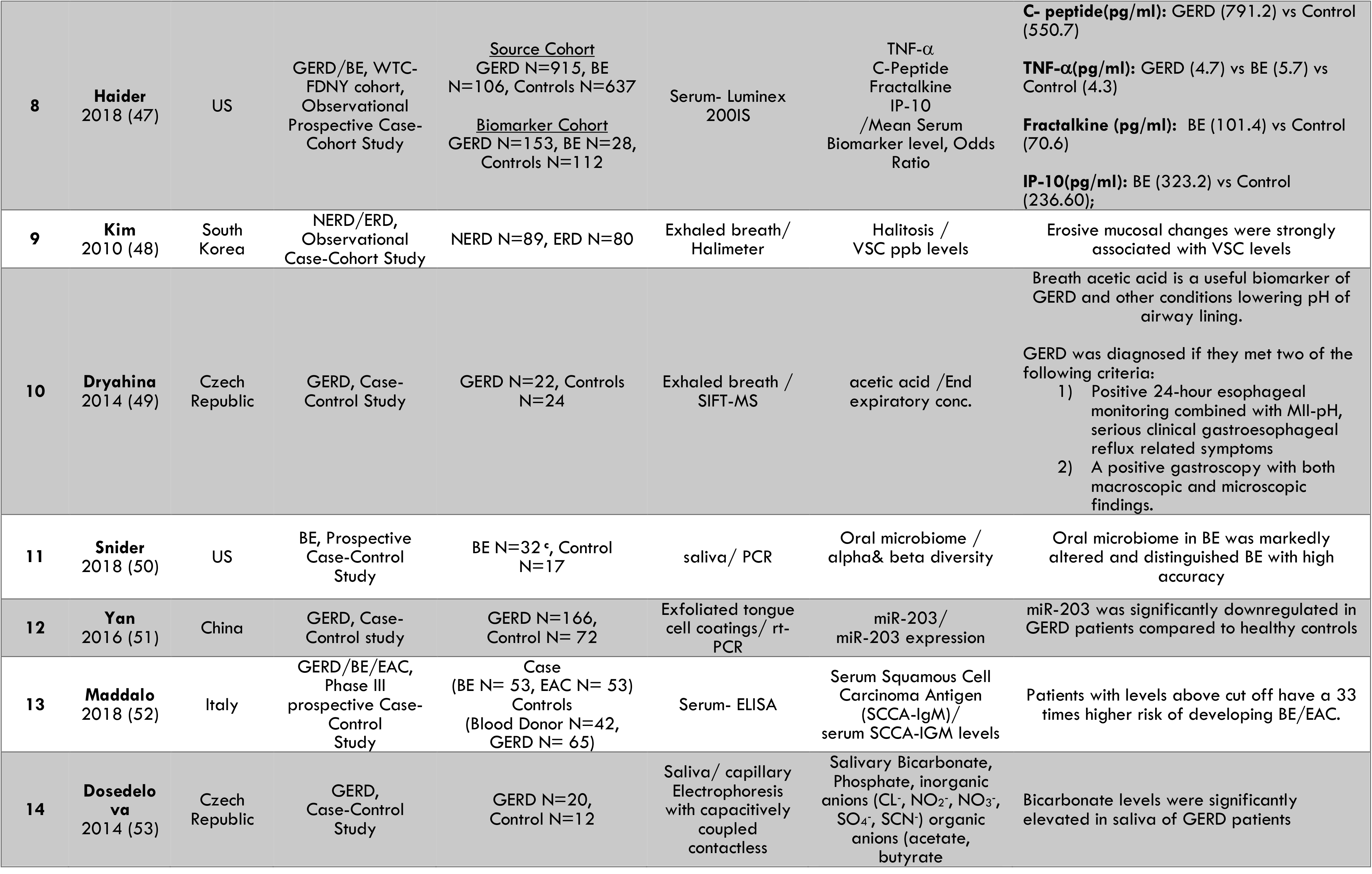
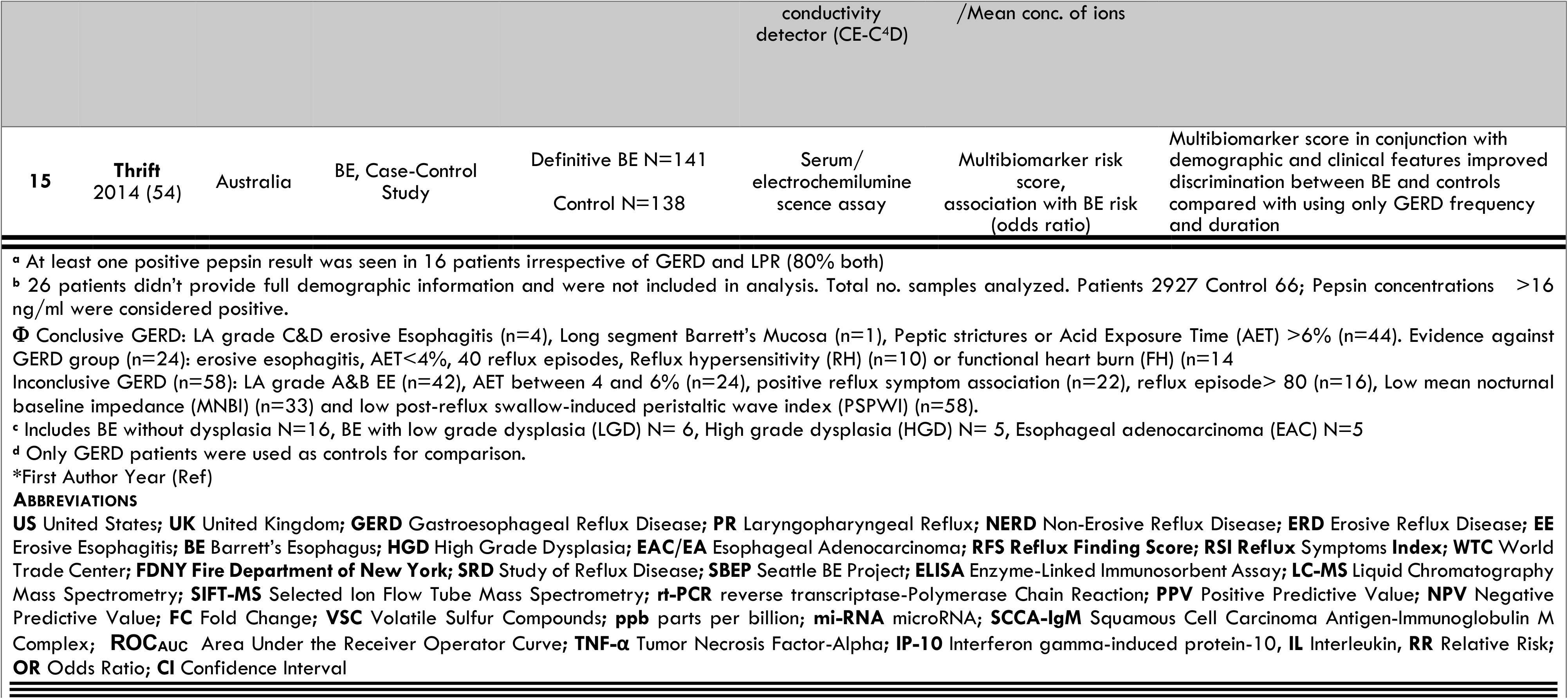
Study Characteristics (N=15)

| Study* | Country | Population/<br>Design | Study Size<br>(N) | Specimen/<br>Assay | Biomarker/End<br>Points | Significant Findings |
| --- | --- | --- | --- | --- | --- | --- |
| 1<br>**Yuskel**<br>2012 (40) | US | GERD,<br>Prospective<br>Blinded Case-Control | GERD N=58, Controls<br>N=51 | Saliva/ Lateral<br>Flow Device | Pepsin/<br>Positive Peptest<br>Prevalence | Pepsin test with rapid LFD has acceptable<br>test characteristics in patients with GERD and<br>may replace the need for EGD and pH<br>testing |
| 2<br>**Bor**<br>2019 (41) | Turkey | GERD/LPR,<br>Observational<br>Case-Cohort study | GERD N=10, LPR N=10 | Saliva/ Lateral<br>Flow Device | Pepsin/<br>Positive Peptest<br>Prevalence | Pepsin test showed a PPV of 68% to detect<br>MII-pH reflux event <sup>a</sup> |
| 3<br>**Wang**<br>2019 (42) | China | GERD/NERD/EE,<br>Multicenter case<br>control study | Control N=323,<br>GERD N= 709,<br>NERD N= 488,<br>EE N= 221 | Saliva/ Lateral<br>Flow device | Pepsin/<br>Predictive Values | Results across all centers showed pepsin-<br>positive sensitivity 85% |
| 4<br>**Dettmar**<br>2020 (43) | UK | LPR, Multicenter<br>case control study | LPR N=985<sup>b</sup>, Controls<br>N=22 | Saliva/ Pepsin<br>Lateral Flow<br>device | Pepsin/<br>Positive Peptest<br>prevalence | Pepsin is an ideal biomarker for detecting<br>airway reflux and LPR |
| 5<br>**Guo**<br>2020 (44) | China | ΦRefractory<br>GERD,<br>Observational<br>Case-Cohort Study | Conclusive GERD N=48;<br>Against GERD N=24;<br>Inconclusive GERD N=58 | Saliva/ELISA | Pepsin/<br>pepsin concentration | Pepsin test had a moderate diagnostic value<br>for PPI refractory conclusive GERD patients |
| 6<br>**De Corso**<br>2020 (45) | Italy | LPR,<br>Observational<br>Prospective Case-Cohort | N=62 | Saliva/<br>calorimeter<br>analyzer | Bile Acids/Bile acid<br>prevalence | High concentrations of bile acids are<br>associated with higher RSI and RFS in LPR<br>patients and higher risk of esophagitis and<br>upper airway malignancies |
| 7<br>**Buas**<br>2017 (46) | US | GERD/BE/HGD/E<br>A, Observational<br>Case-Cohort Study | **SRD:** GERD N=100, BE<br>N=62<br>**SBEP:** BE N=60,<br>HGD/EA N=100 | Serum/<br>LC-MS | Multiple Serum<br>metabolites/<br>FC Serum Metabolites | **SRD:** Nine metabolites were elevated in BE<br>(FC >1)<br>(Creatinine; Homocysteine; 3-Nitrotyrosine<br>Hydroxyproline/amino levulinate;<br>Arginine; Tyrosine;<br>Sorbitol; Linoleic Acid; Ornithine)<br>**SBEP:** Urate raised in HGD/EA vs BE) (FC>1)<br>(Aminoisobutyrate; Sorbitol; Homocysteine<br>raised in BE vs HGD/EA) |

|  |  |  |  |  |  |  |  |
| --- | --- | --- | --- | --- | --- | --- | --- |
| 8 | Haider<br>2018 (47) | US | GERD/BE, WTC-FDNY cohort,<br>Observational<br>Prospective Case-Cohort Study | Source Cohort<br>GERD N=915, BE<br>N=106, Controls N=637<br>Biomarker Cohort<br>GERD N=153, BE N=28,<br>Controls N=112 | Serum- Luminex<br>200IS | TNF-α<br>C-Peptide<br>Fractalkine<br>IP-10<br>/Mean Serum<br>Biomarker level, Odds<br>Ratio | C- peptide(pg/ml): GERD (791.2) vs Control<br>(550.7)<br>TNF-α(pg/ml): GERD (4.7) vs BE (5.7) vs<br>Control (4.3)<br>Fractalkine (pg/ml): BE (101.4) vs Control<br>(70.6)<br>IP-10(pg/ml): BE (323.2) vs Control<br>(236.60); |
| 9 | Kim<br>2010 (48) | South<br>Korea | NERD/ERD,<br>Observational<br>Case-Cohort Study | NERD N=89, ERD N=80 | Exhaled breath/<br>Halimeter | Halitosis /<br>VSC ppb levels | Erosive mucosal changes were strongly<br>associated with VSC levels |
| 10 | Dryahina<br>2014 (49) | Czech<br>Republic | GERD, Case-Control Study | GERD N=22, Controls<br>N=24 | Exhaled breath /<br>SIFT-MS | acetic acid /End<br>expiratory conc. | Breath acetic acid is a useful biomarker of<br>GERD and other conditions lowering pH of<br>airway lining.<br>GERD was diagnosed if they met two of the<br>following criteria:<br>1) Positive 24-hour esophageal<br>monitoring combined with MII-pH,<br>serious clinical gastroesophageal<br>reflux related symptoms<br>2) A positive gastroscopy with both<br>macroscopic and microscopic<br>findings. |
| 11 | Snider<br>2018 (50) | US | BE, Prospective<br>Case-Control<br>Study | BE N=32<sup>c</sup>, Control<br>N=17 | saliva/ PCR | Oral microbiome /<br>alpha& beta diversity | Oral microbiome in BE was markedly<br>altered and distinguished BE with high<br>accuracy |
| 12 | Yan<br>2016 (51) | China | GERD, Case-Control study | GERD N=166,<br>Control N= 72 | Exfoliated tongue<br>cell coatings/ rt-PCR | miR-203/<br>miR-203 expression | miR-203 was significantly downregulated in<br>GERD patients compared to healthy controls |
| 13 | Maddalo<br>2018 (52) | Italy | GERD/BE/EAC,<br>Phase III<br>prospective Case-Control<br>Study | Case<br>(BE N= 53, EAC N= 53)<br>Controls<br>(Blood Donor N=42,<br>GERD N= 65) | Serum- ELISA | Serum Squamous Cell<br>Carcinoma Antigen<br>(SCCA-IgM)/<br>serum SCCA-IGM levels | Patients with levels above cut off have a 33<br>times higher risk of developing BE/EAC. |
| 14 | Dosedelo<br>va<br>2014 (53) | Czech<br>Republic | GERD,<br>Case-Control<br>Study | GERD N=20,<br>Control N=12 | Saliva/ capillary<br>Electrophoresis<br>with capacitively<br>coupled<br>contactless | Salivary Bicarbonate,<br>Phosphate, inorganic<br>anions (CL<sup>-</sup>, NO<sub>2</sub><sup>-</sup>, NO<sub>3</sub><sup>-</sup>,<br>SO<sub>4</sub><sup>-</sup>, SCN<sup>-</sup>) organic<br>anions (acetate,<br>butyrate | Bicarbonate levels were significantly<br>elevated in saliva of GERD patients |

|  |  |  |  | conductivity<br>detector (CE-C<sup>4</sup>D) | /Mean conc. of ions |  |  |
| --- | --- | --- | --- | --- | --- | --- | --- |
| 15 | Thrift<br>2014 (54) | Australia | BE, Case-Control<br>Study | Definitive BE N=141<br>Control N=138 | Serum/<br>electrochemiluminescence assay | Multibiomarker risk<br>score,<br>association with BE risk<br>(odds ratio) | Multibiomarker score in conjunction with<br>demographic and clinical features improved<br>discrimination between BE and controls<br>compared with using only GERD frequency<br>and duration |
| <sup>a</sup> At least one positive pepsin result was seen in 16 patients irrespective of GERD and LPR (80% both)<br><sup>b</sup> 26 patients didn't provide full demographic information and were not included in analysis. Total no. samples analyzed. Patients 2927 Control 66; Pepsin concentrations >16 ng/ml were considered positive.<br>Φ Conclusive GERD: LA grade C&D erosive Esophagitis (n=4), Long segment Barrett's Mucosa (n=1), Peptic strictures or Acid Exposure Time (AET) >6% (n=44). Evidence against GERD group (n=24): erosive esophagitis, AET<4%, 40 reflux episodes, Reflux hypersensitivity (RH) (n=10) or functional heart burn (FH) (n=14)<br>Inconclusive GERD (n=58): LA grade A&B EE (n=42), AET between 4 and 6% (n=24), positive reflux symptom association (n=22), reflux episode> 80 (n=16), Low mean nocturnal baseline impedance (MNBI) (n=33) and low post-reflux swallow-induced peristaltic wave index (PSPWI) (n=58).<br><sup>c</sup> Includes BE without dysplasia N=16, BE with low grade dysplasia (LGD) N= 6, High grade dysplasia (HGD) N= 5, Esophageal adenocarcinoma (EAC) N=5<br><sup>d</sup> Only GERD patients were used as controls for comparison.<br>*First Author Year (Ref)<br>**ABBREVIATIONS**<br>**US** United States; **UK** United Kingdom; **GERD** Gastroesophageal Reflux Disease; **PR** Laryngopharyngeal Reflux; **NERD** Non-Erosive Reflux Disease; **ERD** Erosive Reflux Disease; **EE** Erosive Esophagitis; **BE** Barrett's Esophagus; **HGD** High Grade Dysplasia; **EAC/EA** Esophageal Adenocarcinoma; **RFS Reflux Finding Score**; **RSI Reflux Symptoms Index**; **WTC** World Trade Center; **FDNY Fire Department of New York**; **SRD** Study of Reflux Disease; **SBEP** Seattle BE Project; **ELISA** Enzyme-Linked Immunosorbent Assay; **LC-MS** Liquid Chromatography Mass Spectrometry; **SIFT-MS** Selected Ion Flow Tube Mass Spectrometry; **rt-PCR** reverse transcriptase-Polymerase Chain Reaction; **PPV** Positive Predictive Value; **NPV** Negative Predictive Value; **FC** Fold Change; **VSC** Volatile Sulfur Compounds; **ppb** parts per billion; **mi-RNA** microRNA; **SCCA-IgM** Squamous Cell Carcinoma Antigen-Immunoglobulin M Complex; **ROC<sub>AUC</sub>** Area Under the Receiver Operator Curve; **TNF-α** Tumor Necrosis Factor-Alpha; **IP-10** Interferon gamma-induced protein-10, **IL** Interleukin, **RR** Relative Risk; **OR** Odds Ratio; **CI** Confidence Interval |  |  |  |  |  |  |  |

### Pepsin a classic biomarker of reflux disease

Five of the 15 studies utilized pepsin as a biomarker. Peptest which detects salivary pepsin as low as 16ng/ml using a lateral flow device (LFD) was used in n=4 studies. An additional study used ELISA to quantify salivary pepsin, **Table 1**.(40–44)

Utilizing a prospective blinded case control study, a pepsin cut-off of 50 ng/ml was determined using area under receiver operating curve (ROC_AUC_) analysis from 52 gastric samples and 54 sterile water samples. Patients with GERD underwent endoscopy and wireless 48-hour pH monitoring. This study yielded a positive pepsin test prevalence of 22 % (GERD) vs 12% (controls). There was a stepwise increase in prevalence of positive pepsin with 24% positive with heart burn only symptoms, 43% with abnormal pH and 55% positivity with endoscopic esophagitis. The predictive values (PPV, NPV) were calculated based on the disease definition of esophagitis and/or abnormal pH monitoring; **Table 2, Figure 2A.**(40) Sensitivity ranged from 50% to 85%, with high specificity of 60 to 100%.(40-43, 55, 56)

**Figure 2.**
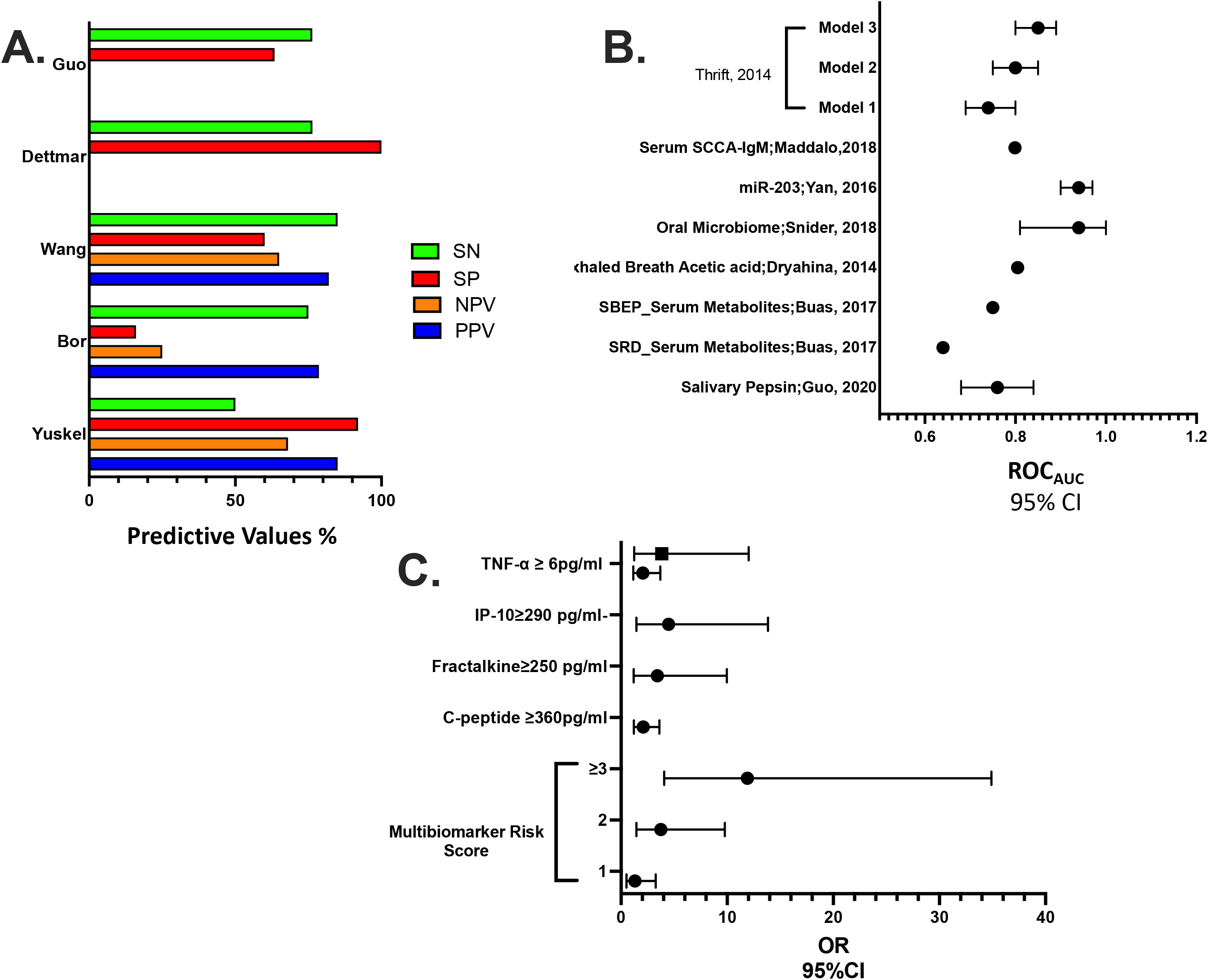
Synthesis of Biomarker Outcomes. **A.** Predictive values of Pepsin. **B.** Area Under the Receiver Operator Curve (ROC_AUC_) of biomarkers **C.** Odds ratio (OR) of biomarkers

**Table 2.**
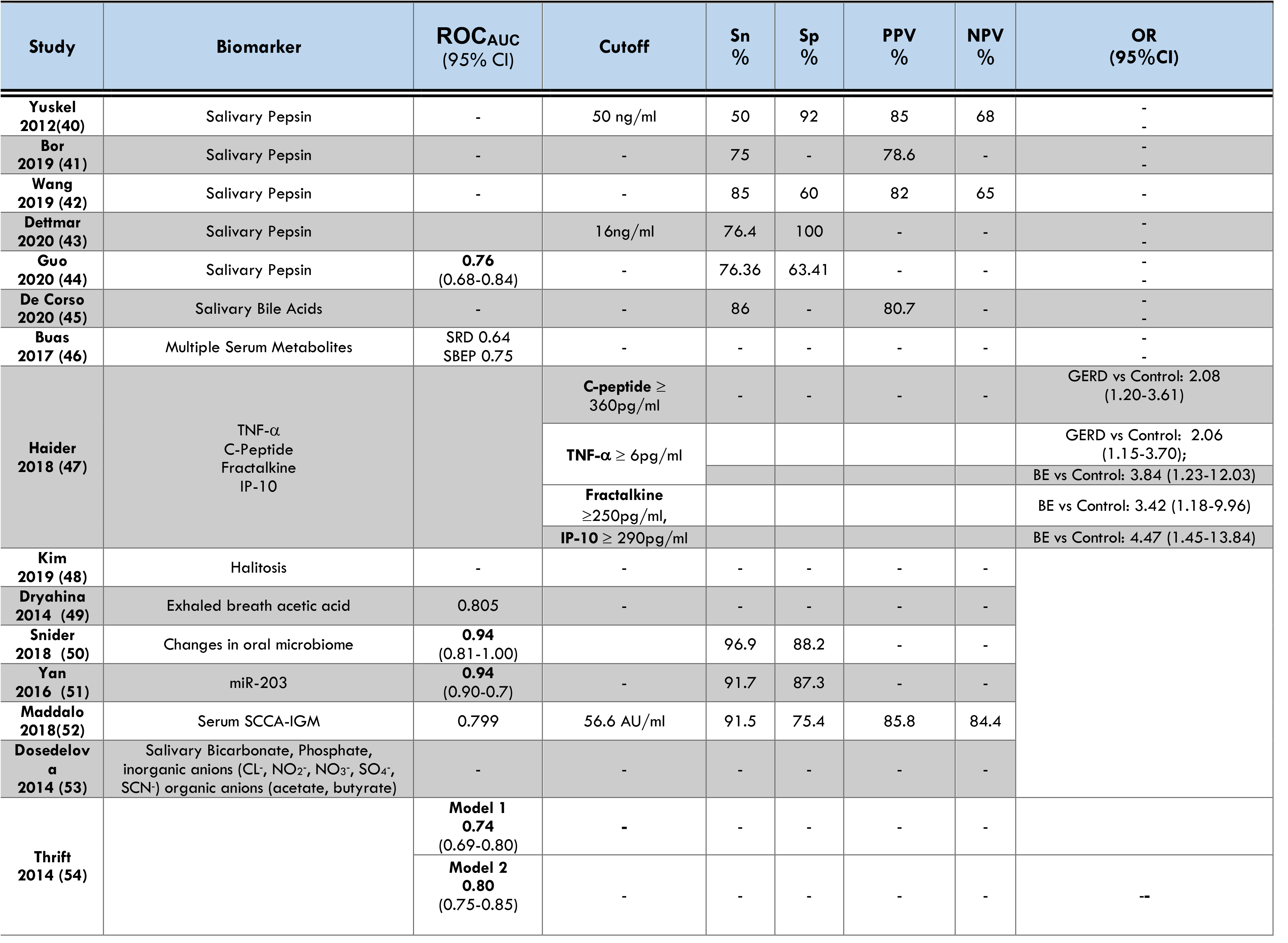
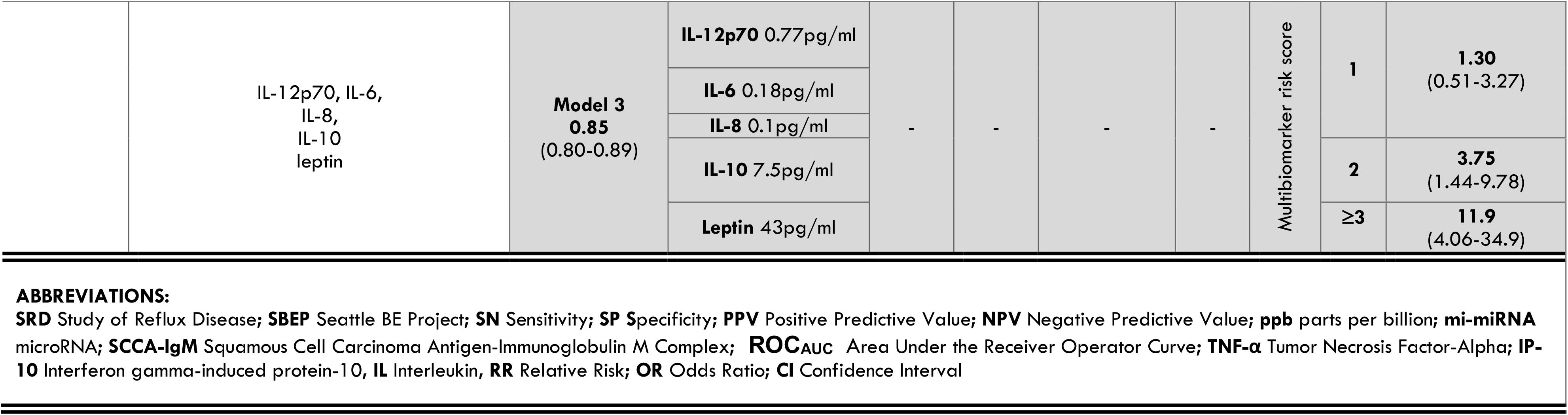
Predictive values of Biomarkers.

LPR and GERD had similar positive Peptest prevalence of 80%.(41) A multicenter case control study enrolling 1032 participants compared GERD, NERD and EE who received endoscopy and a Peptest. The results from all centers showed high sensitivity but the specificity achieved in some centers was low.(42)

In a study of LPR, Peptest prevalence was 74.3% with mean pepsin concentration of 130.9 ng/ml. The highest concentration of Pepsin was reported in post-prandial samples (154.8±4.985 ng/ml) and the lowest in morning samples (102.9± 5.187 ng/ml).(43)

Finally, salivary pepsin levels were assayed by ELISA in refractory GERD as per the Lyon Consensus. Subjects were categorized using mean nocturnal baseline impedance (MBI), post-reflux swallow-induced peristaltic wave index (PSPWI), esophagogastric junction type (EGJ) and contractile integral (EGJ-CI) into three categories (conclusive, inconclusive and against GERD), **Table 1**. The study reported highest mean pepsin concentration of 8.2ng/ml in conclusive GERD patients, 4.0ng/ml in inconclusive GERD patients and 2.4 ng/ml in “against GERD” group. Predictive values were calculated at a pepsin cutoff of 4.21 ng/ml, **Table 2 and Figure 2A**.(44)

### Salivary Bile Acids a Biomarker of LPR and Predictor of Severity

The impact of salivary bile acids was studied prospectively in severe LPR (n = 62), Table 1. Higher values of RFS and Reflux symptom Index (RSI) were observed in patients with recorded salivary bile acids. Three subgroups defined according to bile acid concentrations, Group-ac1 (<0.4μmol/L) Group-ac2 (≥0.4μmol/L and <1μmol/L) and Group-ac3 (≥1μmol/L). A cut off of >1μmol/L was observed to be a reliable indicator for severity. A significant difference was observed between mean RSI in group-ac2 and group ac-3 vs group-ac1 (19.88 vs 16.45 and 21.57 vs 16.45 [p<0.05]). A significant difference was also observed between mean RFS in group-a3 vs group-ac1 (13.52 vs 9.7 [P>0.05]). Pepsinogen I, Pepsinogen II and bilirubin were also detected in the samples but their prevalence was less (25.8%, 46.7% and 40.3% patients respectively), **Table 2**.(45)

### Serum Metabolites and other Biomarkers of GERD, BE and Esophageal Adenocarcinoma (EA)

In order to predict the risk of EA in patients with GERD, BE and high-grade dysplasia (HGD), an observational case cohort study explored the metabolome in serum. In two cohorts; Study of Reflux Disease (SRD; n=162) and Seattle BE Project (SBE; n=160), **Table 1**. Nine metabolites were elevated in BE vs the GERD comparison group (SRD) (ROC_AUC_ 0.64; p<0.05). Whereas one of four metabolites (Urate) was elevated in HGD/EA vs BE, **Table 2** and **Figure 2B**. (46)

Similarly, in a study on the WTC (World Trade Center) particulate-exposed firefighters with OAD serum biomarkers were assessed. The study showed that serum TNF-α ≥6pg/ml predicted both GERD and BE, C-peptide ≥360pg/ml predicted GERD, Fractalkine ≥ 250pg/ml and IP-10 ≥290pg/ml predicted BE. These biomarkers sampled prior to disease presentation showed strong predictive abilities**, Table 2 and Figure 2C**.(47)

### Exhaled Breath Biomarkers of NERD and ERD

In a case cohort study a Halimeter was used to measure volatile sulfur compounds in exhaled breath. Erosive mucosal changes were strongly associated with volatile sulfur compounds (VSC) levels with mean Halimeter *ppb* levels of 191.85 vs 136.43 in ERD vs NERD, **Table 1**. Differences in VSC levels halitosis can be used as a biomarker to discriminate NERD and ERD.(48)

Another case control study compared the end expiratory concentrations of acetic acid in the exhaled breath condensate (EBC) of GERD patients with controls. Mean acetic acid concentration was higher in GERD compared to controls (85 ppbv vs 48 ppbv), **Table 1**. Acetic acid as a diagnostic biomarker for GERD had an ROC_AUC_ of 0.805**. Table 2 & Figure 2B**.(49)

### Oral Microbiome: a Screening Biomarker of BE

Reflux related conditions including BE alter the esophageal microbiome.(57, 58) The oral microbiome, was compared in a case control study in BE patients (n=32) and controls (n=17). Among controls (n=10; 59%) had GERD and (n=6; 35 %) were on PPI. Alpha diversity was no different (mean Shannon index: BE 2.73 vs. Controls 2.89, p=0.10). At the phylum-level there was an increase in the relative abundance of *Firmicutes* (27.1% vs 14.6 %p=0.005) and a decreased *Proteobacteria* (23.8 %vs 34.5%, p=0.02) in BE vs controls respectively. Other notable differences included an increased relative abundance of *Streptococcus*, *Veillonella* and *Enterobacteriaceae* in BE and several taxa (*Neisseria*, *Lautropia* and *Corynebacterium)* in controls. Models with *Lautropia*, *Streptococcus* and *Bacteroidetes* showed the greatest discrimination between BE and controls vs *Lautropia* alone; ROC_AUC_ 0.94 (95% CI 0.81-1.00; p=0.04)**, Figure 2B** and **Table 2.**(50)

### Additional Biomarkers

One study determined microRNA (miRNA) expression (miR-143, −145, −192, −203 andn-205) on exfoliated tongue cells across a discovery cohort (GERD n=24, control n=24). Validated results showed significantly downregulated miR-203 in GERD (n=142) as compared to healthy controls (n=48); ROC_AUC_ 0.84(95% CI: 0.90-0.97), sensitivity 91.37% and specificity 87.3%, **Figure 2B and Table 2**.(51)

The role of immunocomplexed squamous cell carcinoma antigen (SCCA-IgM) as a screening biomarker of BE and EAC, was determined in a phase III cancer screening biomarker development study (n=213). Mean serum SCCA-IgM levels were higher in BE 90 U/mL (95%CI: 89.31-131.55; p<0.0001) and EAC 76 U/mL (95%CI: 56.63-178.87; p<0.0001) as compared to controls which included GERD patients 36.6 U/mL (95%CI: 40.32-68.287) and blood donors 41.5U/mL (95%CI: 49.37-91.33) with a relative risk (RR) 33 (95% CI: 12.66-89.46). BE patients with long segment or dysplastic BE had SCCA-IgM levels significantly higher than those with short nondysplastic BE(p=0.035), **Table 2**.(52)

Salivary anion levels were measured in GERD (n=20) vs healthy controls (n=12) using background electrolyte (BGE) with capillary electrophoresis. They found a significant difference in bicarbonate concentration in case vs controls (6.4mM vs 5.7 mM; p=0.272). No difference was reported in the concentrations of phosphate and other measured anions (6.4mM vs 5.7 mM; p=0.272) **Table 1.**(53)

### Multi-biomarker model predicts BE risk

A case control study of 279 subjects compared the accuracy of three **risk prediction** models that utilized demographic and clinical variables, **Table 1**. Model 1 included GERD frequency and duration, model 2 included GERD frequency, duration, age, sex, race, waist-to-hip (WHR) ratio and H-pylori status. Whereas Model 3 utilized multiple serum biomarkers (IL-12p70, IL6, IL8, IL10 and leptin). Model 3 had a risk score that was significantly associated with BE; (ROC_AUC_ of 0.85 (95% CI 0.80-0.89)).(54)

## DISCUSSION

### GERD diagnosis may include invasive adjunctive tests

GERD detection with endoscopy, ambulatory pH testing and other invasive testing poses rare but potential risk, and contribute to a higher economic burden.(4) Sensitivity of endoscopy may be limited.(59) Conversely, those with endoscopic evidence of reflux may be completely asymptomatic.(60, 61)

**Noninvasive biomarkers** of GERD have diagnostic utility, may direct future research into mechanisms, their downstream effects and was the focus of our review. Pepsin was the most commonly studied noninvasive biomarker of GERD, **Figure 2A**. However, recent ACJ guidelines are less enthusiastic about pepsin testing for evaluation due to its inability to distinguish between patients with laryngeal symptoms and controls.(21)

Studies identified by our systematic review, showed that EBC has the potential to address unmet medical needs by expanding the portfolio of noninvasive assays for the multiple coexisting pathological mechanisms underlying respiratory disorders. Compounds identified in EBC include histamine, adenosine, ammonia and leukotrienes.(62)

### Microbiome of the Gut/Lung Axis

Gut microbiota are linked to the regulation of the innate immune system.(63, 64) Alteration in the microbiota and bacterial products can result in the activation of pathways involved in inflammation. Changes in esophageal microbiota can lead to production of large amounts of bacterial components like LPS which can delay gastric emptying via COX1/2 and predispose to GERD. The role of lautropia is analogous to that of clostridia in the colon and a decrease in bacterial load leads to proliferation of other proinflammatory bacteria. A decrease in concentration of lautropia was seen in patients with periodontitis and successful treatment resulted in subsequent increase. This Taxonomic difference to potentially distinguish BE was utilized by (65)Snider et al using oral swabs and 16S rRNA gene sequencing, **Table 1.**

### Biomarkers of GERD and BE in the FDNY WTC Exposed Cohort

A prime example of the importance of the gut/lung axis was found in the WTC exposed 1^st^ responder cohort.(66–97) WTC-particulate matter (PM) exposure is associated with OAD, GERD and BE.(98–100) WTC-exposed firefighters with OAD had a three times higher risk of developing GERD.(101) Approximately 44% of WTC responders developed GERD symptoms by 2005, which is 8.2 times its pre-9/11 prevalence.(102) In a cohort of non-smoking firefighters with WTC exposure we identified biomarkers of GERD and BE.(73, 103–106) Greater odds of developing GERD were associated with elevated TNF-α and C-peptide, whereas BE was associated with TNF-α, Fractalkine, and IP-10, **Table 2**.(73)

### Systematic reviews by their very nature are subject to limitations and inherent biases

(**1**) the heterogeneity of baseline characteristics, diagnostic criteria, standards for comparison Multichannel intraluminal impedance-pH testing (MII-pH testing), manometry, 24-hour reflux monitoring, questionnaires endoscopy) and lack of validation limit interpretability of the investigator’s findings. (**2**) Although several studies that were a focus of our review utilized Peptest as a biomarker recent guidelines question the applicability of the Peptest for GERD diagnosis.(21) (**3**) Our review focused on non-invasive biomarkers of reflux disease however reliable diagnostic test could be an integration of invasive and noninvasive tests. (**4**) Finally, risk of bias was high in most studies included in this review. This adds additional importance to all future work, in that the clinically relevant non-invasive biomarkers of GERD and associated conditions are needed.

### Conclusion/Future plans

Studies identified include multiOmic, multi-compartmental non-invasive biomarkers for GERD and BE risk. However, due to study limitations and variable controls further validations studies are warranted to further ascertain the reliability and accuracy of these biomarkers. Our future work will focus on validating the previously discovered biomarkers of WTC-aerodigestive disease in longitudinally phenotyped WTC exposed cohort and develop novel, noninvasive disease phenotyping of premalignant diseases such as BE, and identify potential targeted therapeutics to improve care; ClinicalTrials.gov #NCT05216133.

## Data Availability

All data produced in the present work are contained in the manuscript

## LIST OF ABBREVIATIONS

ACG: American College of Gastroenterology
ROC_AUC_: Area under receiver operating curve
BE: Barrett’s Esophagus
BGE: Background electrolytes
CI: Confidence Interval
EE: Erosive Esophagitis
EAC/EA: Esophageal Adenocarcinoma
ELISA: Enzyme-Linked Immunosorbent Assay
ERD: Erosive Reflux Disease
FC: Fold Change
FDNY: Fire Department of New York
GERD: Gastroesophageal Reflux Disease
HGD: High Grade Dysplasia
HRM: Esophageal high-resolution manometry
IL: Interleukin
IP-10: Interferon gamma-induced protein-10
LC-MS: Liquid Chromatography Mass Spectrometry
LPR: Laryngopharyngeal Disease
mi-RNA: MicroRNA
NERD: Non-Erosive Reflux Disease
NPV: Negative Predictive Value
OR: Odds Ratio
*ppb*: *Parts per billion*
*ppbv*: *Parts per billion by volume*
PPV: Positive Predictive Value
PR: Laryngopharyngeal Reflux
qrt-PCR: Quantitative reverse transcription PCR
RR: Relative Risk
SBEP: Seattle BE Project
SCCA-IgM: Squamous Cell Carcinoma Antigen-Immunoglobulin-M complex
SIFT-MS: Selected Ion Flow Tube Mass Spectrometry
SN: Sensitivity
SP: Specificity
SRD: Study of Reflux Disease
TNF-α: Tumor Necrosis Factor-Alpha
UK: United Kingdom
US: United States
VSC: Volatile Sulfur Compounds
WTC: World Trade Center

---

PRISMA CHECK LIST

PRISMA CHECK LIST
| Section/topic | # | Checklist item | Reported on page # |
| --- | --- | --- | --- |
| **TITLE** |  |  |  |
| Title | 1 | Identify the report as a systematic review, meta-analysis, or both. | 1 |
| **ABSTRACT** |  |  |  |
| Structured summary | 2 | Provide a structured summary including, as applicable: background; objectives; data sources; study eligibility criteria, participants, and interventions; study appraisal and synthesis methods; results; limitations; conclusions and implications of key findings; systematic review registration number. | 4 |
| **INTRODUCTION** |  |  |  |
| Rationale | 3 | Describe the rationale for the review in the context of what is already known. | 5-6 |
| Objectives | 4 | Provide an explicit statement of questions being addressed with reference to participants, interventions, comparisons, outcomes, and study design (PICOS). | 7 |
| **METHODS** |  |  |  |
| Protocol and registration | 5 | Indicate if a review protocol exists, if and where it can be accessed (e.g., Web address), and, if available, provide registration information including registration number. | 7 |
| Eligibility criteria | 6 | Specify study characteristics (e.g., PICOS, length of follow-up) and report characteristics (e.g., years considered, language, publication status) used as criteria for eligibility, giving rationale. | 7-8 |
| Information sources | 7 | Describe all information sources (e.g., databases with dates of coverage, contact with study authors to identify additional studies) in the search and date last searched. | 7 |
| Search | 8 | Present full electronic search strategy for at least one database, including any limits used, such that it could be repeated. | 7 |
| Study selection | 9 | State the process for selecting studies (i.e., screening, eligibility, included in systematic review, and, if applicable, included in the meta-analysis). | 7-8 |
| Data collection process | 10 | Describe method of data extraction from reports (e.g., piloted forms, independently, in duplicate) and any processes for obtaining and confirming data from investigators. | 8 |
| Data items | 11 | List and define all variables for which data were sought (e.g., PICOS, funding sources) and any assumptions and simplifications made. | 8-9 |
| Risk of bias in individual studies | 12 | Describe methods used for assessing risk of bias of individual studies (including specification of whether this was done at the study or outcome level), and how this information is to be used in any data synthesis. | 9 |
| Summary measures | 13 | State the principal summary measures (e.g., risk ratio, difference in means). | 8-9 |
| Synthesis of results | 14 | Describe the methods of handling data and combining results of studies, if done, including measures of consistency (e.g., $I^2$) for each meta-analysis. | 8-9 |
| Risk of bias across studies | 15 | Specify any assessment of risk of bias that may affect the cumulative evidence (e.g., publication bias, selective reporting within studies). | 10, Supplemental table 7 |
| Additional analyses | 16 | Describe methods of additional analyses (e.g., sensitivity or subgroup analyses, meta-regression), if done, indicating which were pre-specified. | 10-15, Figure 2 |
| **RESULTS** |  |  |  |
| Study selection | 17 | Give numbers of studies screened, assessed for eligibility, and included in the review, with reasons for exclusions at each stage, ideally with a flow diagram. | 10, Figure 1 |
| Study characteristics | 18 | For each study, present characteristics for which data were extracted (e.g., study size, PICOS, follow-up period) and provide the citations. | 11-12 |
| Risk of bias within studies | 19 | Present data on risk of bias of each study and, if available, any outcome level assessment (see item 12). | 11-12, Supplemental table 7 |
| Results of individual studies | 20 | For all outcomes considered (benefits or harms), present, for each study: (a) simple summary data for each intervention group (b) effect estimates and confidence intervals, ideally with a forest plot. | 10-15. Table 1 & 2 |
| Synthesis of results | 21 | Present results of each meta-analysis done, including confidence intervals and measures of consistency. | 12-16. Table 1 & 2 |
| Risk of bias across studies | 22 | Present results of any assessment of risk of bias across studies (see Item 15). | 10, Supplemental Table 7 |
| Additional analysis | 23 | Give results of additional analyses, if done (e.g., sensitivity or subgroup analyses, meta-regression [see Item 16]). | 12, 22-23, Figure 2 |
| **DISCUSSION** |  |  |  |
| Summary of evidence | 24 | Summarize the main findings including the strength of evidence for each main outcome; consider their relevance to key groups (e.g., healthcare providers, users, and policy makers). | 16-17 |
| Limitations | 25 | Discuss limitations at study and outcome level (e.g., risk of bias), and at review-level (e.g., incomplete retrieval of identified research, reporting bias). | 17 |
| Conclusions | 26 | Provide a general interpretation of the results in the context of other evidence, and implications for future research. | 18 |
| **FUNDING** |  |  |  |
| Funding | 27 | Describe sources of funding for the systematic review and other support (e.g., supply of data); role of funders for the systematic review. | 1 |

From: Moher D, Liberati A, Tetzlaff J, Altman DG, The PRISMA Group (2009). Preferred Reporting Items for Systematic Reviews and Meta-Analyses: The PRISMA Statement. PLoS Med 6(7): e1000097. doi:10.1371/journal.pmed1000097

