## Supplemental Table 1 for "Non-Invasive, MultiOmic and MultiCompartmental Biomarkers of Reflux Disease: A Systematic Review"

**SUPPLEMENTAL TABLES**

**Supplemental Table 1. Database Search Criteria**

**Supplemental Table 2. EMBASE Identified Manuscripts (N=150)**

**Supplemental Table 3. PUBMED Identified Manuscripts (N=90)**

**Supplemental Table 4. Manuscripts Identified after Merge (N=240)**

**Supplemental Table 5. Excluded Manuscripts (N=208)**

**Supplemental Table 6. Manuscripts that met all Inclusion and Exclusion Criteria**

**Supplemental Table 7. Risk of Bias Assessment A. Cohort Studies (N=6) and B. Case-Cohort Studies (N=9)**

**Supplemental Table No.1- Database Search Criteria**

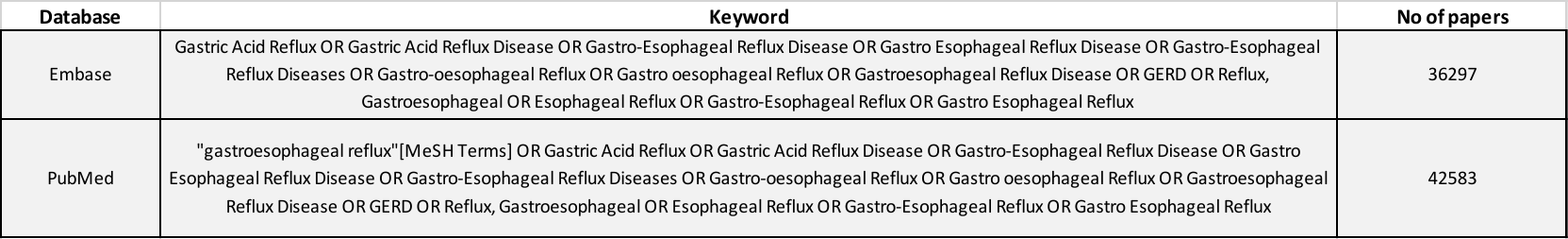

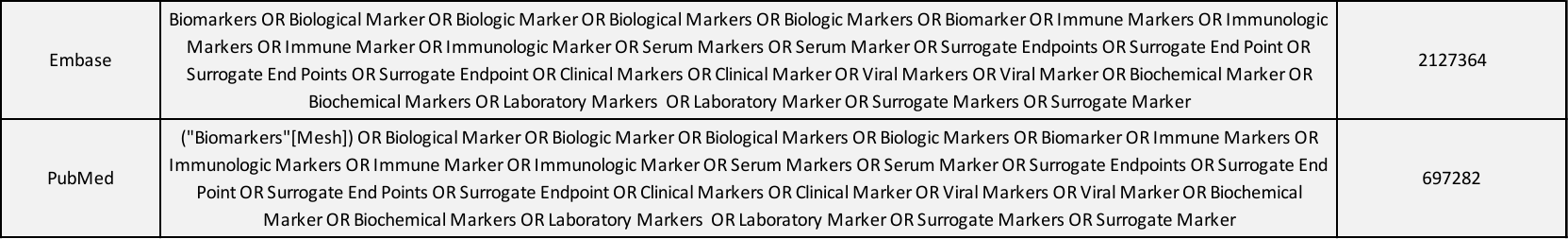

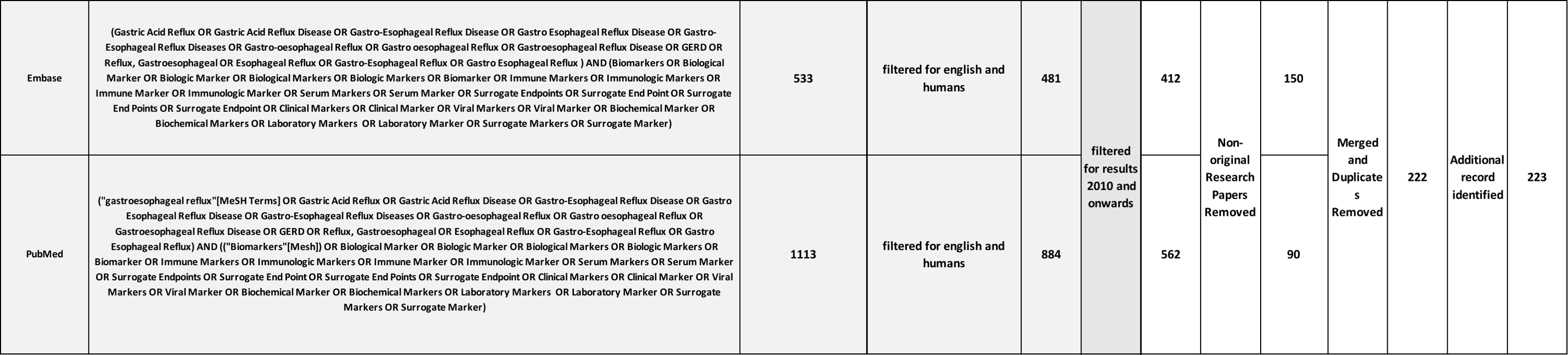

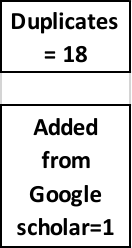
