## Supplemental Table 2 for "Non-Invasive, MultiOmic and MultiCompartmental Biomarkers of Reflux Disease: A Systematic Review"

| **Supplemental Table 2: Embase Identified Manuscripts (N= 150)** | | | | |
| --- | --- | --- | --- | --- |
| **TITLE** | **YEAR** | **AUTHOR** | **JOURNAL** | **DOI** |
| Immunohistochemical analysis of Ki-67, p53 and Bcl-2 expression related to histological features in gastroesophageal reflux disease | 2010 | Ayhan, S., Ak Nalbant, O., Isisag, A., Turkel Kucukmetin, N. and Temiz, P. | Turkish Journal of Gastroenterology | http://dx.doi.org/10.4318/tjg.2010.0088 |
| Foveolar type dysplasia in Barrett esophagus | 2010 | Brown, I. S., Whiteman, D. C. and Lauwers, G. Y. | Modern Pathology | http://dx.doi.org/10.1038/modpathol.2010.59 |
| Oil red O stain of alveolar macrophages is an effective screening test for gastroesophageal reflux disease in lung transplant recipients | 2010 | Hopkins, P. M., Kermeen, F., Duhig, E., Fletcher, L., Gradwell, J., Whitfield, L., Godinez, C., Musk, M., Chambers, D., Gotley, D. and Mcneil, K. | Journal of Heart and Lung Transplantation | http://dx.doi.org/10.1016/j.healun.2010.03.015 |
| Halimeter ppb levels as the predictor of erosive gastroesophageal reflux disease | 2010 | Kim, J. G., Kim, Y. J., Yoo, S. H., Lee, S. J., Chung, J. W., Kim, M. H., Park, D. K. and Hahm, K. B. | Gut and Liver | http://dx.doi.org/10.5009/gnl.2010.4.3.320 |
| Molecular abnormalities in pediatric barrett esophagus: Can we test for potential of neoplastic progression? | 2010 | Maltby, E. L., Dyson, M. J., Wheeler, M. R., Thomson, M., Sethuraman, C. and Cohen, M. C. | Pediatric and Developmental Pathology | http://dx.doi.org/10.2350/09-08-0700-OA.1 |
| Exercise and the Prevention of Oesophageal Cancer (EPOC) study protocol: A randomized controlled trial of exercise versus stretching in males with Barrett's oesophagus | 2010 | Winzer, B. M., Paratz, J. D., Reeves, M. M. and Whiteman, D. C. | BMC Cancer | http://dx.doi.org/10.1186/1471-2407-10-292 |
| A striking local esophageal cytokine expression profile in eosinophilic esophagitis | 2011 | Blanchard, C., Stucke, E. M., Rodriguez-Jimenez, B., Burwinkel, K., Collins, M. H., Ahrens, A., Alexander, E. S., Buckmeier Butz, B. K., Jameson, S. C., Kaul, A., Franciosi, J. P., Kushner, J. P., Putnam, P. E., Abonia, J. P. and Rothenberg, M. E. | Journal of Allergy and Clinical Immunology | http://dx.doi.org/10.1016/j.jaci.2010.10.039 |
| Distinct proteomic profiles characterise non-erosive from erosive reflux disease | 2011 | Calabrese, C., Marzano, V., Urbani, A., Lazzarini, G., Valerii, M. C., Liguori, G., Di Molfetta, S., Rizzello, F., Gionchetti, P., Campieri, M. and Spisni, E. | Alimentary Pharmacology and Therapeutics | http://dx.doi.org/10.1111/j.1365-2036.2011.04801.x |
| Barrett's esophagus: Progression to adenocarcinoma and markers | 2011 | Fang, D., Das, K. M., Cao, W., Malhotra, U., Triadafilopoulos, G., Najarian, R. M., Hardie, L. J., Lightdale, C. J., Beales, I. L., Felix, V. N., Schneider, P. M. and Bellizzi, A. M. | Annals of the New York Academy of Sciences | http://dx.doi.org/10.1111/j.1749-6632.2011.06053.x |
| Role of e-cadherin in the pathogenesis of gastroesophageal reflux disease | 2011 | Jovov, B., Que, J., Tobey, N. A., Djukic, Z., Hogan, B. L. M. and Orlando, R. C. | American Journal of Gastroenterology | http://dx.doi.org/10.1038/ajg.2011.102 |
| MUC2 is a highly specific marker of goblet cell metaplasia in the distal esophagus and gastroesophageal junction | 2011 | Mcintire, M. G., Soucy, G., Vaughan, T. L., Shahsafaei, A. and Odze, R. D. | American Journal of Surgical Pathology | http://dx.doi.org/10.1097/PAS.0b013e318218940d |
| The histological and immunohistochemical aspects of bile reflux in patients with gastroesophageal reflux disease | 2011 | Nakos, A., Kouklakis, G., Pitiakoudis, M., Zezos, P., Efraimidou, E., Giatromanolaki, A., Polychronidis, A., Liratzopoulos, N., Sivridis, E. and Simopoulos, K. | Gastroenterology Research and Practice | http://dx.doi.org/10.1155/2011/905872 |
| Correlation of serum pepsinogens and gastrin-17 with atrophic gastritis in gastroesophageal reflux patients: A matched-pairs study | 2011 | Peitz, U., Wex, T., Vieth, M., Stolte, M., Willich, S., Labenz, J., Jaspersen, D., Lind, T. and Malfertheiner, P. | Journal of Gastroenterology and Hepatology (Australia) | http://dx.doi.org/10.1111/j.1440-1746.2010.06413.x |
| Early events during neoplastic progression in Barrett's esophagus | 2011 | Reid, B. J. | Cancer Biomarkers | http://dx.doi.org/10.3233/CBM-2011-0162 |
| Barrett's esophagus: Genetic and cell changes | 2011 | Souza, R. F., Freschi, G., Taddei, A., Ringressi, M. N., Bechi, P., Castiglione, F., Degl'innocenti, D. R., Triadafilopoulos, G., Wang, J. S., Chang, A. C., Barr, H., Bajpai, M., Das, K. M., Schneider, P. M., Krishnadath, K. K., Malhotra, U. and Lynch, J. P. | Annals of the New York Academy of Sciences | http://dx.doi.org/10.1111/j.1749-6632.2011.06043.x |
| Identification of clinical and laboratory markers for predicting eosinophilic esophagitis in adults | 2011 | Von Arnim, U., Wex, T., Rohl, F. W., Neumann, H., Kuster, D., Weigt, J., Monkemuller, K. and Malfertheiner, P. | Digestion | http://dx.doi.org/10.1159/000331142 |
| Prolonged exposure to acid and bile induces chromosome abnormalities that precede malignant transformation of benign Barrett's epithelium | 2012 | Bajpai, M., Aviv, H. and Das, K. M. | Molecular Cytogenetics | http://dx.doi.org/10.1186/1755-8166-5-43 |
| Prevalence and predictors of columnar lined esophagus in Gastroesophageal Reflux Disease (GERD) patients undergoing upper endoscopy | 2012 | Balasubramanian, G., Singh, M., Gupta, N., Gaddam, S., Giacchino, M., Wani, S. B., Moloney, B., Higbee, A. D., Rastogi, A., Bansal, A. and Sharma, P. | American Journal of Gastroenterology | http://dx.doi.org/10.1038/ajg.2012.299 |
| Activation of the BMP4 Pathway and Early Expression of CDX2 Characterize Non-specialized Columnar Metaplasia in a Human Model of Barrett's Esophagus | 2012 | Castillo, D., Puig, S., Iglesias, M., Seoane, A., De Bolos, C., Munitiz, V., Parrilla, P., Comerma, L., Poulsom, R., Krishnadath, K. K., Grande, L. and Pera, M. | Journal of Gastrointestinal Surgery | http://dx.doi.org/10.1007/s11605-011-1758-5 |
| Diagnostic utility of major basic protein, eotaxin-3, and leukotriene enzyme staining in eosinophilic esophagitis | 2012 | Dellon, E. S., Chen, X., Miller, C. R., Woosley, J. T. and Shaheen, N. J. | American Journal of Gastroenterology | http://dx.doi.org/10.1038/ajg.2012.202 |
| Eosinophilic esophagitis: Epithelial mesenchymal transition contributes to esophageal remodeling and reverses with treatment | 2012 | Kagalwalla, A. F., Akhtar, N., Woodruff, S. A., Rea, B. A., Masterson, J. C., Mukkada, V., Parashette, K. R., Du, J., Fillon, S., Protheroe, C. A., Lee, J. J., Amsden, K., Melin-Aldana, H., Capocelli, K. E., Furuta, G. T. and Ackerman, S. J. | Journal of Allergy and Clinical Immunology | http://dx.doi.org/10.1016/j.jaci.2012.03.005 |
| Oxidative stress in inflammation-based gastrointestinal tract diseases: Challenges and opportunities | 2012 | Kim, Y. J., Kim, E. H. and Hahm, K. B. | Journal of Gastroenterology and Hepatology (Australia) | http://dx.doi.org/10.1111/j.1440-1746.2012.07108.x |
| COX-2 mRNA is increased in oesophageal mucosal cells by a proton pump inhibitor | 2012 | Mayne, G. C., Watson, D. I. and Hussey, D. J. | ANZ Journal of Surgery | http://dx.doi.org/10.1111/j.1445-2197.2012.06124.x |
| The presence of pepsin in the lung and its relationship to pathologic gastro-esophageal reflux | 2012 | Rosen, R., Johnston, N., Hart, K., Khatwa, U. and Nurko, S. | Neurogastroenterology and Motility | http://dx.doi.org/10.1111/j.1365-2982.2011.01826.x |
| Rapid salivary pepsin test: Blinded assessment of test performance in gastroesophageal reflux disease | 2012 | Saritas Yuksel, E., Hong, S. K. S., Strugala, V., Slaughter, J. C., Goutte, M., Garrett, C. G., Dettmar, P. W. and Vaezi, M. F. | Laryngoscope | http://dx.doi.org/10.1002/lary.23252 |
| Influence of genetics on tumoral pathologies: The example of the adenocarcinoma arising in barrett's esophagus | 2012 | Villanacci, V., Bassotti, G., Salemme, M. and Rossi, E. | Revista Espanola de Enfermedades Digestivas | http://dx.doi.org/10.4321/s1130-01082012001100007 |
| Current hurdles in the management of eosinophilic oesophagitis: The next steps | 2013 | Attwood, S. E. A. and Wilson, M. S. J. | World Journal of Gastroenterology | https://dx.doi.org/10.3748/wjg.v19.i6.790 |
| Barrett's esophagus in 2012: Updates in pathogenesis, treatment, and surveillance | 2013 | Chandra, S., Gorospe, E. C., Leggett, C. L. and Wang, K. K. | Current Gastroenterology Reports | http://dx.doi.org/10.1007/s11894-013-0322-8 |
| Immunoreactivity of p53 and Ki-67 for dysplastic changes in children with eosinophilic esophagitis | 2013 | Denning, K. L., Al-Subu, A. and Elitsur, Y. | Pediatric and Developmental Pathology | http://dx.doi.org/10.2350/13-03-1306-OA.1 |
| Research advances in esophageal diseases: Bench to bedside | 2013 | Di Pietro, M. and Fitzgerald, R. C. | F1000Prime Reports | http://dx.doi.org/10.12703/P5-44 |
| The oesophageal string test: A novel, minimally invasive method measures mucosal inflammation in eosinophilic oesophagitis | 2013 | Furuta, G. T., Kagalwalla, A. F., Lee, J. J., Alumkal, P., Maybruck, B. T., Fillon, S., Masterson, J. C., Ochkur, S., Protheroe, C., Moore, W., Pan, Z., Amsden, K., Robinson, Z., Capocelli, K., Mukkada, V., Atkins, D., Fleischer, D., Hosford, L., Kwatia, M. A., Schroeder, S., Kelly, C., Lovell, M., Melin-Aldana, H. and Ackerman, S. J. | Gut | http://dx.doi.org/10.1136/gutjnl-2012-303171 |
| Aspiration and allograft injury secondary to gastroesophageal reflux occur in the immediate post-lung transplantation period (Prospective Clinical Trial) | 2013 | Griffin, S. M., Robertson, A. G. N., Bredenoord, A. J., Brownlee, I. A., Stovold, R., Brodlie, M., Forrest, I., Dark, J. H., Pearson, J. P. and Ward, C. | Annals of Surgery | http://dx.doi.org/10.1097/SLA.0b013e3182a6589b |
| Urease and Helicobacter spp. Antigens in Pulmonary Granuloma | 2013 | Herndon, B., Quinn, T., Wasson, N., Nzabi, M. and Molteni, A. | Journal of Comparative Pathology | http://dx.doi.org/10.1016/j.jcpa.2012.06.011 |
| Markers of antigen presentation and activation on eosinophils and T cells in the esophageal tissue of patients with eosinophilic esophagitis | 2013 | Le-Carlson, M., Seki, S., Abarbanel, D., Quiros, A., Cox, K. and Nadeau, K. C. | Journal of Pediatric Gastroenterology and Nutrition | http://dx.doi.org/10.1097/MPG.0b013e3182758d49 |
| Gastroesophageal reflux and metabolic syndrome | 2013 | Mocanu, M. A., Diculescu, M. and Dumitrescu, M. | Revista medico-chirurgicala a Societatii de Medici si Naturalisti din Iasi |  |
| Reflux-Associated Oxygen Desaturations: Usefulness in Diagnosing Reflux-Related Respiratory Symptoms | 2013 | Wilshire, C. L., Salvador, R., Sepesi, B., Niebisch, S., Watson, T. J., Litle, V. R., Peyre, C. G., Jones, C. E. and Peters, J. H. | Journal of Gastrointestinal Surgery | http://dx.doi.org/10.1007/s11605-012-2065-5 |
| MicroRNA expression can be a promising strategy for the detection of barrett's esophagus: A pilot study | 2014 | Bansal, A., Hong, X., Lee, I. H., Krishnadath, K. K., Mathur, S. C., Gunewardena, S., Rastogi, A., Sharma, P. and Christenson, L. K. | Clinical and Translational Gastroenterology | http://dx.doi.org/10.1038/ctg.2014.17 |
| Immunohistochemistry for CDX2 expression in non-goblet-cell Barrett's oesophagus | 2014 | Behan, M., Gledhill, A. and Hayes, S. | British journal of biomedical science |  |
| Pharmacologic treatments for esophageal disorders | 2014 | Blackshaw, L. A., Bordin, D. S., Brock, C., Brokjaer, A., Drewes, A. M., Farmer, A. D., Krarup, A. L., Lottrup, C., Masharova, A. A., Moawad, F. J. and Olesen, A. E. | Annals of the New York Academy of Sciences | http://dx.doi.org/10.1111/nyas.12520 |
| Exhaled breath concentrations of acetic acid vapour in gastro-esophageal reflux disease | 2014 | Dryahina, K., Pospisilova, V., Sovova, K., Shestivska, V., Kubista, J., Spesyvyi, A., Pehal, F., Turzikova, J., Votruba, J. and Spanel, P. | Journal of Breath Research | http://dx.doi.org/10.1088/1752-7155/8/3/037109 |
| Assessment of cricopharyngeal muscle aging with apoptotic markers | 2014 | Gor, A. P., Alkan, Z., Yigit, O., Dursun, N., Engin, A., Akin, S. M. and Sam, B. | The Journal of craniofacial surgery | http://dx.doi.org/10.1097/SCS.0000000000000940 |
| Cost-effectiveness of endoscopic surveillance of non-dysplastic Barrett's esophagus | 2014 | Gordon, L. G., Mayne, G. C., Hirst, N. G., Bright, T., Whiteman, D. C. and Watson, D. I. | Gastrointestinal Endoscopy | http://dx.doi.org/10.1016/j.gie.2013.07.046 |
| Correlation between the serum pepsinogen I level and the symptom degree in proton pump inhibitor-users administered with a probiotic | 2014 | Igarashi, M., Nagano, J., Tsuda, A., Suzuki, T., Koike, J., Uchida, T., Matsushima, M., Mine, T. and Koga, Y. | Pharmaceuticals | http://dx.doi.org/10.3390/ph7070754 |
| Correlation of ALOX15 expression with eosinophilic or reflux esophagitis in a cohort of pediatric patients with esophageal eosinophilia | 2014 | Matoso, A., Allen, D., Herzlinger, M., Ferreira, J., Chen, S., Lu, S., Fabre, V., Monahan, R., Yang, D., Noble, L., Mangray, S. and Resnick, M. B. | Human Pathology | http://dx.doi.org/10.1016/j.humpath.2014.01.021 |
| A PCR blood test outperforms chromogranin A in carcinoid detection and is unaffected by proton pump inhibitors | 2014 | Modlin, I. M., Aslanian, H., Bodei, L., Drozdov, I. and Kidd, M. | Endocrine Connections | http://dx.doi.org/10.1530/EC-14-0100 |
| Sensiprobe - A miniature thermal device incorporating Peltier technology as a diagnostic tool for studying human oesophageal sensitivity | 2014 | Reeves, J. W., Al-Zinaty, M., Woodland, P., Sifrim, D., Aziz, Q. and Birch, M. J. | Physiological Measurement | http://dx.doi.org/10.1088/0967-3334/35/7/1265 |
| A Multibiomarker Risk Score Helps Predict Risk for Barrett's Esophagus | 2014 | Thrift, A. P., Garcia, J. M. and El-Serag, H. B. | Clinical Gastroenterology and Hepatology | http://dx.doi.org/10.1016/j.cgh.2013.12.014 |
| A Multibiomarker Risk Score Helps Predict Risk for Barrett'sEsophagus | 2014 | Thrift, A. P., Garcia, J. M. and El-Serag, H. B. | Clinical Gastroenterology and Hepatology | http://dx.doi.org/10.1016/j.cgh.2013.12.014 |
| Changes in obesity-related diseases and biochemical variables after laparoscopic sleeve gastrectomy: a two-year follow-up study | 2014 | Vage, V., Sande, V. A., Mellgren, G., Laukeland, C., Behme, J. and Andersen, J. R. | BMC surgery | http://dx.doi.org/10.1186/1471-2482-14-8 |
| Bronchoalveolar pH and inflammatory biomarkers in newly diagnosed IPF and GERD patients: A case-control study | 2014 | Vukovac, E. L., Lozo, M., Mise, K., Gudelj, I., Puljiz, Z., Jurcev-Savicevic, A., Bradaric, A., Kokeza, J. and Mise, J. | Medical Science Monitor | http://dx.doi.org/10.12659/MSM.889800 |
| Eosinophilic oesophagitis: A paediatric update | 2015 | Allen, K. J. and Heine, R. G. | Journal of Paediatrics and Child Health | http://dx.doi.org/10.1111/jpc.12757 |
| Exhaled Breath Condensate pH in Lung Cancer, the Impact of Clinical Factors | 2015 | Bikov, A., Lazar, Z., Gyulai, N., Szentkereszty, M., Losonczy, G., Horvath, I. and Galffy, G. | Lung | http://dx.doi.org/10.1007/s00408-015-9778-7 |
| Central obesity and other factors associated with uncontrolled asthma in women | 2015 | Capelo, A. V., De Fonseca, V. M., Peixoto, M. V. M., De Carvalho, S. R. and Guerino, L. G. | Allergy, Asthma and Clinical Immunology | http://dx.doi.org/10.1186/s13223-015-0076-y |
| Utility of a noninvasive serum biomarker panel for diagnosis and monitoring of eosinophilic esophagitis: A prospective study | 2015 | Dellon, E. S., Rusin, S., Gebhart, J. H., Covey, S., Higgins, L. L., Beitia, R., Speck, O., Woodward, K., Woosley, J. T. and Shaheen, N. J. | American Journal of Gastroenterology | http://dx.doi.org/10.1038/ajg.2015.57 |
| Pediatric Eosinophilic Esophagitis Symptom Scores (PEESS v2.0) identify histologic and molecular correlates of the key clinical features of disease | 2015 | Martin, L. J., Franciosi, J. P., Collins, M. H., Abonia, J. P., Lee, J. J., Hommel, K. A., Varni, J. W., Grotjan, J. T., Eby, M., He, H., Marsolo, K., Putnam, P. E., Garza, J. M., Kaul, A., Wen, T. and Rothenberg, M. E. | Journal of Allergy and Clinical Immunology | http://dx.doi.org/10.1016/j.jaci.2015.03.004 |
| Comparison of eotaxin-3 biomarker in patients with eosinophilic oesophagitis, proton pump inhibitor-responsive oesophageal eosinophilia and gastro-oesophageal reflux disease | 2015 | Moawad, F. J., Wells, J. M., Johnson, R. L., Reinhardt, B. J., Maydonovitch, C. L. and Baker, T. P. | Alimentary Pharmacology and Therapeutics | http://dx.doi.org/10.1111/apt.13258 |
| The Cologne-Bonn cohort: lessons learned | 2015 | Rockstroh, J. K. | Infection. | http://dx.doi.org/10.1007/s15010-015-0745-2 |
| Clinical and autoantibody profile in systemic sclerosis: Baseline characteristics from a West Malaysian cohort | 2015 | Sujau, I., Ng, C. T., Sthaneshwar, P., Sockalingam, S., Cheah, T. E., Yahya, F. and Jasmin, R. | International Journal of Rheumatic Diseases | http://dx.doi.org/10.1111/1756-185X.12322 |
| Pepsinogen identification in the middle ear fluid of children with otitis media with effusion | 2016 | Buyruk, A., Osma, U., Yilmaz, M. D. and Eyigor, H. | Kulak burun bogaz ihtisas dergisi : KBB = Journal of ear, nose, and throat | http://dx.doi.org/10.5606/kbbihtisas.2016.83669 |
| Proteomic Analysis in Esophageal Eosinophilia Reveals Differential Galectin-3 Expression and S-Nitrosylation | 2016 | Davis, C. M., Hiremath, G., Wiktorowicz, J. E., Soman, K. V., Straub, C., Nance, C., Quintanilla, N., Pazdrak, K., Thakkar, K., Olive, A. P. and Kurosky, A. | Digestion | http://dx.doi.org/10.1159/000444675 |
| Prospective assessment of serum periostin as a biomarker for diagnosis and monitoring of eosinophilic oesophagitis | 2016 | Dellon, E. S., Higgins, L. L., Beitia, R., Rusin, S., Woosley, J. T., Veerappan, R., Selitsky, S. R., Parker, J. S., Genta, R. M., Lash, R. H., Aranda, R., Peach, R. J. and Grimm, M. | Alimentary Pharmacology and Therapeutics | http://dx.doi.org/10.1111/apt.13672 |
| Characterization of oncocytes in deep esophageal glands | 2016 | Gonzalez, G., Huang, Q. and Mashimo, H. | Diseases of the Esophagus | http://dx.doi.org/10.1111/dote.12382 |
| Biomarkers of Reflux Disease | 2016 | Kia, L., Pandolfino, J. E. and Kahrilas, P. J. | Clinical Gastroenterology and Hepatology | http://dx.doi.org/10.1016/j.cgh.2015.09.014 |
| Differential protein expression and oncogenic gene network link tyrosine kinase ephrin B4 receptor to aggressive gastric and gastroesophageal junction cancers | 2016 | Liersch-Lohn, B., Slavova, N., Buhr, H. J. and Bennani-Baiti, I. M. | International Journal of Cancer | http://dx.doi.org/10.1002/ijc.29865 |
| A Systematic Review of Esophageal MicroRNA Markers for Diagnosis and Monitoring of Barrett's Esophagus | 2016 | Mallick, R., Patnaik, S. K., Wani, S. and Bansal, A. | Digestive Diseases and Sciences | http://dx.doi.org/10.1007/s10620-015-3959-3 |
| Epidemiology | 2016 | Navaratnam, V., Forrester, D. L. and Hubbard, R. B. | ERS Monograph | http://dx.doi.org/10.1183/2312508X.10004615 |
| Distal mean nocturnal baseline impedance on pH-impedance monitoring predicts reflux burden and symptomatic outcome in gastro-oesophageal reflux disease | 2016 | Patel, A., Wang, D., Sainani, N., Sayuk, G. S. and Gyawali, C. P. | Alimentary Pharmacology and Therapeutics | http://dx.doi.org/10.1111/apt.13777 |
| Lymphocytic Esophagitis in Nonachalasia Primary Esophageal Motility Disorders: Improved Criteria, Prevalence, Strength of Association, and Natural History | 2016 | Putra, J., Muller, K. E., Hussain, Z. H., Parker, S., Gabbard, S., Brickley, E. B., Lacy, B. E., Rothstein, R. and Lisovsky, M. | The American journal of surgical pathology |  |
| Exhaled nitric oxide in relation to asthma control: A real-life survey | 2016 | Ricciardolo, F. L. M., Sorbello, V., Bellezza Fontana, R., Schiavetti, I. and Ciprandi, G. | Allergologia et Immunopathologia | http://dx.doi.org/10.1016/j.aller.2015.05.012 |
| Leukocyte telomere length in relation to the risk of Barrett's esophagus and esophageal adenocarcinoma | 2016 | Wennerstrom, E. C. M., Risques, R. A., Prunkard, D., Giffen, C., Corley, D. A., Murray, L. J., Whiteman, D. C., Wu, A. H., Bernstein, L., Ye, W., Chow, W., Vaughan, T. L. and Liao, L. M. | Cancer Medicine | http://dx.doi.org/10.1002/cam4.810 |
| MiR-203 Expression in Exfoliated Cells of Tongue Coating Represents a Sensitive and Specific Biomarker of Gastroesophageal Reflux Disease | 2016 | Yan, X., Zhu, S. and Zhang, H. | Gastroenterology Research and Practice | http://dx.doi.org/10.1155/2016/2349453 |
| Candidate serum metabolite biomarkers for differentiating gastroesophageal reflux disease, Barrett's esophagus, and high-grade dysplasia/esophageal adenocarcinoma | 2017 | Buas, M. F., Gu, H., Djukovic, D., Zhu, J., Onstad, L., Reid, B. J., Raftery, D. and Vaughan, T. L. | Metabolomics | http://dx.doi.org/10.1007/s11306-016-1154-y |
| The diagnostic value of pepsin detection in saliva for gastro-esophageal reflux disease: A preliminary study from China | 2017 | Du, X., Wang, F., Hu, Z., Wu, J., Wang, Z., Yan, C., Zhang, C. and Tang, J. | BMC Gastroenterology | http://dx.doi.org/10.1186/s12876-017-0667-9 |
| Esomeprazole FDA Approval in Children with GERD: Exposure-Matching and Exposure-Response | 2017 | Earp, J. C., Mehrotra, N., Peters, K. E., Fiorentino, R. P., Griebel, D., Lee, S. C., Mulberg, A., Rohss, K., Sandstrom, M., Taylor, A., Tornoe, C. W., Wynn, E. L., Van Der Walt, J. S. and Garnett, C. | Journal of Pediatric Gastroenterology and Nutrition | http://dx.doi.org/10.1097/MPG.0000000000001467 |
| Pepsin in saliva as a biomarker for oropharyngeal reflux compared with 24-hour esophageal impedance/pH monitoring in pediatric patients | 2017 | Fortunato, J. E., D'agostino, R. B. and Lively, M. O. | Neurogastroenterology and Motility | http://dx.doi.org/10.1111/nmo.12936 |
| Promising Modalities to Identify and Monitor Eosinophilic Esophagitis | 2017 | Hiremath, G. and Gupta, S. K. | Clinical Gastroenterology and Hepatology | http://dx.doi.org/10.1016/j.cgh.2017.05.004 |
| Diagnostic value of a pattern of exhaled breath condensate biomarkers in asthmatic children | 2017 | Maloca Vuljanko, I., Turkalj, M., Nogalo, B., Bulat Lokas, S. and Plavec, D. | Allergologia et Immunopathologia | http://dx.doi.org/10.1016/j.aller.2016.05.001 |
| Improved Cough and Cough-Specific Quality of Life in Patients Treated for Scleroderma-Related Interstitial Lung Disease: Results of Scleroderma Lung Study II | 2017 | Tashkin, D. P., Volkmann, E. R., Tseng, C. H., Roth, M. D., Khanna, D., Furst, D. E., Clements, P. J., Theodore, A., Kafaja, S., Kim, G. H., Goldin, J., Ariolla, E. and Elashoff, R. M. | Chest | http://dx.doi.org/10.1016/j.chest.2016.11.052 |
| Clinical symptom tool that raises the index of suspicion for eosinophilic oesophagitis in adults and drives earlier biopsy for definitive diagnosis | 2017 | Von Arnim, U., Rohl, F. W., Miehlke, S., Jechorek, D., Reinhold, D., Wex, T. and Malfertheiner, P. | Alimentary Pharmacology and Therapeutics | http://dx.doi.org/10.1111/apt.13869 |
| A pilot randomized clinical trial assessing the effect of cricoid pressure on risk of aspiration | 2018 | Bohman, J. K., Kashyap, R., Lee, A., He, Z., Soundar, S., Bolling, L. L. and Kor, D. J. | Clinical Respiratory Journal | http://dx.doi.org/10.1111/crj.12508 |
| OPCML is hypermethylated in a subset of patients with metaplastic changes in their esophagus | 2018 | Castano-Rodriguez, N., Popple, G. L., Porras-Hurtado, G. L., Cardona-Deazza, J. L., Montoya-Martinez, J. J., Cadavid-Velez, A. J., Toro-Hidalgo, H. W., Cobo-Alvarado, A. R., Del Socorro Hincapie-Rincon, O., Riordan, S. M. and Kaakoush, N. O. | Biomarker Research | http://dx.doi.org/10.1186/s40364-018-0150-y |
| Methylation panel is a diagnostic biomarker for Barrett's oesophagus in endoscopic biopsies and non-endoscopic cytology specimens | 2018 | Chettouh, H., Mowforth, O., Galeano-Dalmau, N., Bezawada, N., Ross-Innes, C., Macrae, S., Debiram-Beecham, I., O'donovan, M. and Fitzgerald, R. C. | Gut | http://dx.doi.org/10.1136/gutjnl-2017-314026 |
| Short-Term Neurodevelopmental Outcome in Children Born With High-Risk Congenital Lung Lesions | 2018 | Danzer, E., Hoffman, C., D'agostino, J. A., Boelig, M. M., Gerdes, M., Bernbaum, J. C., Rosenthal, H., Waqar, L. N., Rintoul, N. E., Herkert, L. M., Kallan, M. J., Peranteau, W. H., Flake, A. W., Adzick, N. S. and Hedrick, H. L. | Annals of Thoracic Surgery | http://dx.doi.org/10.1016/j.athoracsur.2018.01.033 |
| Interchangeable Use of Proton Pump Inhibitors Based on Relative Potency | 2018 | Graham, D. Y. and Tansel, A. | Clinical Gastroenterology and Hepatology | http://dx.doi.org/10.1016/j.cgh.2017.09.033 |
| Pepsin in saliva as a diagnostic marker for gastroesophageal reflux disease: A meta-analysis | 2018 | Guo, Z., Wu, H., Jiang, J. and Zhang, C. | Medical Science Monitor | http://dx.doi.org/10.12659/MSM.913978 |
| Predictive Biomarkers of Gastroesophageal Reflux Disease and Barrett's Esophagus in World Trade Center Exposed Firefighters: a 15 Year Longitudinal Study | 2018 | Haider, S. H., Kwon, S., Lam, R., Lee, A. K., Caraher, E. J., Crowley, G., Zhang, L., Schwartz, T. M., Zeig-Owens, R., Liu, M., Prezant, D. J. and Nolan, A. | Scientific reports | http://dx.doi.org/10.1038/s41598-018-21334-9 |
| Identification of Individuals with Functional Dyspepsia With a Simple, Minimally Invasive Test: A Single Center Cohort Study of the Oral Capsaicin Test | 2018 | Hammer, J. | American Journal of Gastroenterology | http://dx.doi.org/10.1038/ajg.2018.16 |
| Pepsin: Biomarker, mediator, and therapeutic target for reflux and aspiration | 2018 | Johnston, N., Dettmar, P. W., Ondrey, F. G., Nanchal, R., Lee, S. H. and Bock, J. M. | Annals of the New York Academy of Sciences. | http://dx.doi.org/10.1111/nyas.13729 |
| Fragments of e-Cadherin as Biomarkers of Non-erosive Reflux Disease | 2018 | Jovov, B., Reed, C. C., Shaheen, N. J., Pruitt, A., Ferrell, K., Orlando, G. S., Djukic, Z. and Orlando, R. C. | Digestive Diseases and Sciences | http://dx.doi.org/10.1007/s10620-017-4815-4 |
| Nutritional State and Feeding Behaviors of Children with Eosinophilic Esophagitis and Gastroesophageal Reflux Disease | 2018 | Mehta, P., Furuta, G. T., Brennan, T., Henry, M. L., Maune, N. C., Sundaram, S. S., Menard-Katcher, C., Atkins, D., Takurukura, F., Giffen, S., Pan, Z. and Haas, A. M. | Journal of Pediatric Gastroenterology and Nutrition | http://dx.doi.org/10.1097/MPG.0000000000001741 |
| Comorbidities are associated with different features of severe asthma 11 Medical and Health Sciences 1102 Cardiorespiratory Medicine and Haematology | 2018 | Novelli, F., Bacci, E., Latorre, M., Seccia, V., Bartoli, M. L., Cianchetti, S., Dente, F. L., Franco, A. D., Celi, A. and Paggiaro, P. | Clinical and Molecular Allergy | http://dx.doi.org/10.1186/s12948-018-0103-x |
| Pediatric Eosinophilic Esophagitis | 2018 | Posten, S., Adamiak, T. and Jensen, M. | South Dakota medicine : the journal of the South Dakota State Medical Association |  |
| Immunohistochemical assessment of Survivin and Bcl3 expression as potential biomarkers for NF-kappaB activation in the Barrett metaplasia-dysplasia-adenocarcinoma sequence | 2018 | Puccio, I., Khan, S., Butt, A., Graham, D., Sehgal, V., Patel, D., Novelli, M., Lovat, L. B., Rodriguez-Justo, M. and Hamoudi, R. A. | International Journal of Experimental Pathology | http://dx.doi.org/10.1111/iep.12260 |
| Blind esophageal brushing offers a safe and accurate method to monitor inflammation in children and young adults with eosinophilic esophagitis | 2018 | Smadi, Y., Deb, C., Bornstein, J., Safder, S., Horvath, K. and Mehta, D. | Diseases of the Esophagus | http://dx.doi.org/10.1093/dote/doy056 |
| Prospective single arm study on the effect of ilaprazole in patients with heartburn but no reflux esophagitis | 2018 | Song, I. J., Kim, H. K., Lee, N. K. and Lee, S. K. | Yonsei Medical Journal | http://dx.doi.org/10.3349/ymj.2018.59.8.951 |
| Review: Helicobacter pylori and non-malignant upper gastrointestinal diseases | 2019 | Boltin, D., Niv, Y., Schutte, K. and Schulz, C. | Helicobacter | http://dx.doi.org/10.1111/hel.12637 |
| Functional dyspepsia susceptibility is associated with TGFB1 gene polymorphisms (RS4803455, RS1800469) in H pylori-negative Chinese population | 2019 | Cheung, C. K. Y., Lan, L. L., Chan, Y., Yuen, K., Cheong, P. K., Fang, F. and Wu, J. C. Y. | Neurogastroenterology and Motility | http://dx.doi.org/10.1111/nmo.13681 |
| Plasma levels of TNF-alpha, IL-6, IFN-gamma, IL-12, IL-17, IL-22, and IL-23 in achalasia, eosinophilic esophagitis (EoE), and gastroesophageal reflux disease (GERD) | 2019 | Clayton, S., Cauble, E., Kumar, A., Patil, N., Ledford, D., Kolliputi, N., Lopes-Virella, M. F., Castell, D. and Richter, J. | BMC Gastroenterology | http://dx.doi.org/10.1186/s12876-019-0937-9 |
| Overexpression of CCN1 in Het1A cells attenuates bile-induced esophageal metaplasia through suppressing non-canonical NFkappaB activation | 2019 | Dang, T., Meng, X., Modak, C., Wu, J., Chang, Z., Che, N., Narvaez, R. and Chai, J. | Cytokine | http://dx.doi.org/10.1016/j.cyto.2018.12.020 |
| Autism spectrum disorder and neurodevelopmental delays in children with giant omphalocele | 2019 | Danzer, E., Hoffman, C., Miller, J. S., D'agostino, J. A., Schindewolf, E. M., Gerdes, M., Bernbaum, J., Adams, S. E., Rintoul, N. E., Herkert, L. M., Taylor, L., Schreiber, J., Peranteau, W. H., Flake, A. W., Adzick, N. S. and Hedrick, H. L. | Journal of Pediatric Surgery | http://dx.doi.org/10.1016/j.jpedsurg.2019.05.017 |
| Omeprazole induced increase in liver markers-a case report | 2019 | Elmahdy, M. F. and Almater, J. M. | Journal of Clinical and Diagnostic Research | http://dx.doi.org/10.7860/JCDR/2019/41848.13218 |
| Developing a blood-based gene mutation assay as a novel biomarker for oesophageal adenocarcinoma | 2019 | Haboubi, H. N., Lawrence, R. L., Rees, B., Williams, L., Manson, J. M., Al-Mossawi, N., Bodger, O., Griffiths, P., Thornton, C. and Jenkins, G. J. | Scientific reports | http://dx.doi.org/10.1038/s41598-019-41490-w |
| Association between skeletal muscle attenuation and gastroesophageal reflux disease: A health check-up cohort study | 2019 | Kim, Y. M., Kim, J. H., Baik, S. J., Jung, D. H., Park, J. J., Youn, Y. H. and Park, H. | Scientific reports | http://dx.doi.org/10.1038/s41598-019-56702-6 |
| Mean platelet volume and red cell distribution width as potential new biomarkers in children with gastroesophageal reflux disease | 2019 | Sevencan, N. O., Cesur, O., Cakar, M., Dogan, E., Ozkan, A. E. and Benli, A. R. | American Journal of Translational Research |  |
| Validation in China of a non-invasive salivary pepsin biomarker containing two unique human pepsin monoclonal antibodies to diagnose gastroesophageal reflux disease | 2019 | Wang, Y. F., Yang, C. Q., Chen, Y. X., Cao, A. P., Yu, X. F., Yu, Y., Zhang, Z. Y., Shen, X. Z., Liu, F., Zhong, L., Wang, Y. X., Liu, Z. J., Shi, Y. H., Zhong, J., Li, J. N., Lan, Y., Lenham, R. K., Woodcock, A. D., Dettmar, P. W. and Fang, J. Y. | Journal of Digestive Diseases | http://dx.doi.org/10.1111/1751-2980.12783 |
| Targeting the COX1/2-Driven thromboxane A2 pathway suppresses Barrett's esophagus and esophageal adenocarcinoma development | 2019 | Zhang, T., Wang, Q., Ma, W. Y., Wang, K., Chang, X., Johnson, M. L., Bai, R., Bode, A. M., Foster, N. R., Falk, G. W., Limburg, P. J., Iyer, P. G. and Dong, Z. | EBioMedicine | http://dx.doi.org/10.1016/j.ebiom.2019.10.038 |
| Highlighted role of "IL17 signaling pathway" in gastroesophageal reflux disease | 2020 | Azodi, M. Z., Razzaghi, M., Malekpour, H., Heidari, M. H. and Rezaei-Tavirani, M. | Gastroenterology and Hepatology from Bed to Bench |  |
| The association between depression and gastroesophageal reflux based on phylogenetic analysis of mirna biomarkers | 2020 | Chen, Y. H. and Wang, H. | Current Medicinal Chemistry | http://dx.doi.org/10.2174/0929867327666200425214906 |
| Eosinophilic Esophagitis Histology Remission Score: Significant Relations to Measures of Disease Activity and Symptoms | 2020 | Collins, M. H., Martin, L. J., Wen, T., Abonia, J. P., Putnam, P. E., Mukkada, V. A. and Rothenberg, M. E. | Journal of pediatric gastroenterology and nutrition | http://dx.doi.org/10.1097/MPG.0000000000002637 |
| Differential MicroRNA signatures in the pathogenesis of barrett's esophagus | 2020 | Craig, M. P., Rajakaruna, S., Paliy, O., Sajjad, M., Madhavan, S., Reddy, N., Zhang, J., Bottomley, M., Agrawal, S. and Kadakia, M. P. | Clinical and Translational Gastroenterology | http://dx.doi.org/10.14309/ctg.0000000000000125 |
| A Multicentre Study in UK Voice Clinics Evaluating the Non-invasive Reflux Diagnostic Peptest in LPR Patients | 2020 | Dettmar, P. W., Watson, M., Mcglashan, J., Tatla, T., Nicholaides, A., Bottomley, K., Jarad, N., Stapleton, E., Lenham, R. K., Fisher, J. and Woodcock, A. D. | SN Comprehensive Clinical Medicine | http://dx.doi.org/10.1007/s42399-019-00184-0 |
| May failure to thrive in infants be a clinical marker for the early diagnosis of cow's milk allergy? | 2020 | Diaferio, L., Caimmi, D., Verga, M. C., Palladino, V., Trove, L., Giordano, P., Verduci, E. and Miniello, V. L. | Nutrients | http://dx.doi.org/10.3390/nu12020466 |
| Analysis of bicarbonate, phosphate and other anions in saliva by capillary electrophoresis with capacitively coupled contactless conductivity detection in diagnostics of gastroesophageal reflux disease | 2020 | Dosedelova, V., Durc, P., Dolina, J., Konecny, S., Foret, F. and Kuban, P. | Electrophoresis | http://dx.doi.org/10.1002/elps.201900319 |
| The role of salivary pepsin in the diagnosis of gastroesophageal reflux disease (GERD) evaluated using high-resolution manometry and 24-hour multichannel intraluminal impedance-pH monitoring | 2020 | Guo, Z., Wu, Y., Li, L., Chen, J., Zhang, S. and Zhang, C. | Medical Science Monitor | http://dx.doi.org/10.12659/MSM.927381 |
| Salivary pepsin A detection related to gastro-oesophageal reflux episodes in children undergoing impedance probe monitoring | 2020 | Haddad, H. A., He, Z., Shaffer, S. E. and Molle-Rios, Z. L. | Acta Paediatrica, International Journal of Paediatrics | http://dx.doi.org/10.1111/apa.15276 |
| Phenotypic characteristics and asthma severity in an East African cohort of adults and adolescents with asthma: Findings from the African severe asthma project | 2020 | Kirenga, B., Chakaya, J., Yimer, G., Nyale, G., Haile, T., Muttamba, W., Mugenyi, L., Katagira, W., Worodria, W., Aanyu-Tukamuhebwa, H., Lugogo, N., Joloba, M., Bekele, A., Makumbi, F., Green, C., De Jong, C., Kamya, M. and Van Der Molen, T. | BMJ Open Respiratory Research | http://dx.doi.org/10.1136/bmjresp-2019-000484 |
| Hematological indices as indicators of silent inflammation in achalasia patients: A cross-sectional study | 2020 | Lopez-Verdugo, F., Furuzawa-Carballeda, J., Romero-Hernandez, F., Coss-Adame, E., Valdovinos, M. A., Priego-Ranero, A., Olvera-Prado, H., Narvaez-Chavez, S., Peralta-Figueroa, J. and Torres-Villalobos, G. | Medicine (United States) | http://dx.doi.org/10.1097/MD.0000000000019326 |
| Cross validated serum small extracellular vesicle microRNAs for the detection of oropharyngeal squamous cell carcinoma | 2020 | Mayne, G. C., Woods, C. M., Dharmawardana, N., Wang, T., Krishnan, S., Hodge, J. C., Foreman, A., Boase, S., Carney, A. S., Sigston, E. a. W., Watson, D. I., Ooi, E. H. and Hussey, D. J. | Journal of Translational Medicine | http://dx.doi.org/10.1186/s12967-020-02446-1 |
| Current therapies for gastro-oesophageal reflux in the setting of chronic lung disease: State of the art review | 2020 | Mcdonnell, M. J., Hunt, E. B., Ward, C., Pearson, J. P., O'toole, D., Laffey, J. G., Murphy, D. M. and Rutherford, R. M. | ERJ Open Research | http://dx.doi.org/10.1183/23120541.00190-2019 |
| Implications of historical height loss for prevalent vertebral fracture, spinal osteoarthritis, and gastroesophageal reflux disease | 2020 | Nakano, M., Nakamura, Y., Suzuki, T., Kobayashi, T., Takahashi, J. and Shiraki, M. | Scientific reports | https://dx.doi.org/10.1038/s41598-020-76074-6 |
| Nissen fundoplication in Cornelia de Lange syndrome spectrum: Who are the potential candidates? | 2020 | Parma, B., Cianci, P., Mariani, M., Cereda, A., Panceri, R., Fossati, C., Maestri, L., Macchini, F., Onesimo, R., Zampino, G., Betalli, P., Cheli, M. and Selicorni, A. | American Journal of Medical Genetics, Part A | http://dx.doi.org/10.1002/ajmg.a.61625 |
| Routine Preoperative Nutritional Screening in All Primary Total Joint Arthroplasty Patients Has Little Utility | 2020 | Rao, S. S., Chaudhry, Y. P., Solano, M. A., Sterling, R. S., Oni, J. K. and Khanuja, H. S. | Journal of Arthroplasty | https://dx.doi.org/10.1016/j.arth.2020.06.073 |
| Asthma similarities across ProAR (Brazil) and U-BIOPRED (Europe) adult cohorts of contrasting locations, ethnicity and socioeconomic status | 2020 | Riley, J. H., Bansal, A. T., Almeida, P. C. A., Davis, M., Bates, S., Adcock, I. M., Chung, K. F., Alcantara-Neves, N., Amorim, L., Araujo, M. I., Barnes, K. C., Barreto, M. L., Belitardo, E., Biao-Lima, V., Cardoso, L., Camargos, P. A., Chatkin, J. M., Costa, R. S., Coelho, A. C. C., Cooper, P. J., Cruz, A. A., Cruz, C. S., Cunha, J., De Jesus, J. V., Fernandes, J., Franco, R. A., Gomes-Filho, I., Lima-Matos, A., Figueiredo, C. A., Lessa, M. A., Lins, L., Mello, L. M., Moura-Santos, P., Muniz, I. S., Paixao-Araujo, I., Pinheiro, G. P., Ponte, E. V., Rodrigues, L. C., Santana, C. V. N., Santos-Lima, G., Souza, T. M. O., Souza-Machado, A., Souza-Machado, C., Stelmach, R., Vasquez, V. S., Ahmed, H., Auffray, C., Bakke, P., Baribaud, F., Bel, E. H., Bigler, J., Bisgaard, H., Boedigheimer, M. J., Bonnelykke, K., Brandsma, J., Brinkman, P., Bucchioni, E., Burg, D., Bush, A., Caruso, M., Chaiboonchoe, A., Chanez, P., Compton, C. H., Corfield, J., D'amico, A., Dahlen, B., Dahlen, S. E., De Meulder, B., Djukanovic, R., Erpenbeck, V. J., Erzen, D., Fichtner, K., Fitch, N., Fleming, L. J., Formaggio, E., Fowler, S. J., Frey, U., Gahlemann, M., Geiser, T., Goss, V., Guo, Y. K., Hashimoto, S., Haughney, J., Hedlin, G., Hekking, P. W., Higenbottam, T., Hohlfeld, J. M., Holweg, C., Horvath, I., Howarth, P., James, A. J., Knowles, R. G., Knox, A. J., Krug, N., Lefaudeux, D., Loza, M. J., Lutter, R., Manta, A., Masefield, S., Matthews, J. G., Mazein, A., Meiser, A., Middelveld, R. J. M., Miralpeix, M., Montuschi, P., Mores, N., Murray, C. S., Musial, J., Myles, D., Pahus, L., Pandis, I., Pavlidis, S., Postle, A., Powel, P., Pratico, G., Puig Valls, M., Rao, N., Roberts, A., Roberts, G., Rowe, A., Sandstrom, T., Schofield, J. P. R., Seibold, W., Selby, A., Shaw, D. E., Sigmund, R., Singer, F., Skipp, P. J., Sousa, A. R., Sterk, P. J., Sun, K., Thornton, B., Van Aalderen, W. M., Van Geest, M., Vestbo, J., Vissing, N. H., Wagener, A. H., Wagers, S. S., Weiszhart, Z., Wheelock, C. E. and Wilson, S. J. | Respiratory Medicine | https://dx.doi.org/10.1016/j.rmed.2019.105817 |
| Asthma similarities across ProAR (Brazil) and U-BIOPRED (Europe) adult cohorts of contrasting locations, ethnicity and socioeconomic status | 2020 | Riley, J. H., Bansal, A. T., Davis, M., Bates, S., Adcock, I. M., Chung, K. F., Alcantara-Neves, N., Almeida, P. C. A., Amorim, L., Araujo, M. I., Barnes, K. C., Barreto, M. L., Belitardo, E., Biao-Lima, V., Cardoso, L., Camargos, P. A., Chatkin, J. M., Costa, R. S., Coelho, A. C. C., Cooper, P. J., Cruz, A. A., Cruz, C. S., Cunha, J., De Jesus, J. V., Fernandes, J., Franco, R. A., Gomes-Filho, I., Lima-Matos, A., Figueiredo, C. A., Lessa, M. A., Lins, L., Mello, L. M., Moura-Santos, P., Muniz, I. S., Paixao-Araujo, I., Pinheiro, G. P., Ponte, E. V., Rodrigues, L. C., Santana, C. V. N., Santos-Lima, G., Souza, T. M. O., Souza-Machado, A., Souza-Machado, C., Stelmach, R., Vasquez, V. S., Ahmed, H., Auffray, C., Bakke, P., Baribaud, F., Bel, E. H., Bigler, J., Bisgaard, H., Boedigheimer, M. J., Bonnelykke, K., Brandsma, J., Brinkman, P., Bucchioni, E., Burg, D., Bush, A., Caruso, M., Chaiboonchoe, A., Chanez, P., Compton, C. H., Corfield, J., D'amico, A., Dahlen, B., Dahlen, S. E., De Meulder, B., Djukanovic, R., Erpenbeck, V. J., Erzen, D., Fichtner, K., Fitch, N., Fleming, L. J., Formaggio, E., Fowler, S. J., Frey, U., Gahlemann, M., Geiser, T., Goss, V., Guo, Y. K., Hashimoto, S., Haughney, J., Hedlin, G., Hekking, P. W., Higenbottam, T., Hohlfeld, J. M., Holweg, C., Horvath, I., Howarth, P., James, A. J., Knowles, R. G., Knox, A. J., Krug, N., Lefaudeux, D., Loza, M. J., Lutter, R., Manta, A., Masefield, S., Matthews, J. G., Mazein, A., Meiser, A., Middelveld, R. J. M., Miralpeix, M., Montuschi, P., Mores, N., Murray, C. S., Musial, J., Myles, D., Pahus, L., Pandis, I., Pavlidis, S., Postle, A., Powel, P., Pratico, G., Puig Valls, M., Rao, N., Roberts, A., Roberts, G., Rowe, A., Sandstrom, T., Schofield, J. P. R., Seibold, W., Selby, A., Shaw, D. E., Sigmund, R., Singer, F., Skipp, P. J., Sousa, A. R., Sterk, P. J., Sun, K., Thornton, B., Van Aalderen, W. M., Van Geest, M., Vestbo, J., Vissing, N. H., Wagener, A. H., Wagers, S. S., Weiszhart, Z., Wheelock, C. E. and Wilson, S. J. | Respiratory Medicine | http://dx.doi.org/10.1016/j.rmed.2019.105817 |
| The frequency of visceral and phenotypic markers in patients with the combination of undifferentiated connective tissue disease and gastroesophageal reflux disease | 2020 | Romash, I. B. and Mishchuk, V. G. | Wiadomosci lekarskie (Warsaw, Poland : 1960) |  |
| Eosinophilic Esophagitis: Update on Diagnosis and Treatment in Pediatric Patients | 2020 | Rossetti, D., Isoldi, S. and Oliva, S. | Pediatric Drugs | http://dx.doi.org/10.1007/s40272-020-00398-z |
| Review - Helicobacter pylori and non-malignant upper gastro-intestinal diseases | 2020 | Schulz, C. and Kupcinskas, J. | Helicobacter | http://dx.doi.org/10.1111/hel.12738 |
| Complex Gastrointestinal and Endocrine Sources of Inflammation in Schizophrenia | 2020 | Severance, E. G., Dickerson, F. and Yolken, R. H. | Frontiers in Psychiatry | http://dx.doi.org/10.3389/fpsyt.2020.00549 |
| Bronchoalveolar bile acid and inflammatory markers to identify high-risk lung transplant recipients with reflux and microaspiration | 2020 | Zhang, C. Y. K., Ahmed, M., Huszti, E., Levy, L., Hunter, S. E., Boonstra, K. M., Moshkelgosha, S., Sage, A. T., Azad, S., Zamel, R., Ghany, R., Yeung, J. C., Crespin, O. M., Frankel, C., Budev, M., Shah, P., Reynolds, J. M., Snyder, L. D., Belperio, J. A., Singer, L. G., Weigt, S. S., Todd, J. L., Palmer, S. M., Keshavjee, S. and Martinu, T. | Journal of Heart and Lung Transplantation | http://dx.doi.org/10.1016/j.healun.2020.05.006 |
| Recurring Translocations in Barrett's Esophageal Adenocarcinoma | 2021 | Bajpai, M., Panda, A., Birudaraju, K., Van Gurp, J., Chak, A., Das, K. M., Javidian, P. and Aviv, H. | Frontiers in Genetics | http://dx.doi.org/10.3389/fgene.2021.674741 |
| Risk factors associated with the development of interstitial lung abnormalities | 2021 | Buendia-Roldan, I., Fernandez, R., Mejia, M., Juarez, F., Ramirez-Martinez, G., Montes, E., Pruneda, A. K. S., Martinez-Espinosa, K., Alarcon-Dionet, A., Herrera, I., Becerril, C., Chavez-Galan, L., Preciado, M., Pardo, A. and Selman, M. | European Respiratory Journal | http://dx.doi.org/10.1183/13993003.03005-2020 |
| Forkhead box F1 induces columnar phenotype and epithelial-to-mesenchymal transition in esophageal squamous cells to initiate Barrett's like metaplasia | 2021 | De, A., Zhou, J., Liu, P., Huang, M., Gunewardena, S., Mathur, S. C., Christenson, L. K., Sharma, M., Zhang, Q. and Bansal, A. | Laboratory Investigation | http://dx.doi.org/10.1038/s41374-021-00534-4 |
| Multi-omics of the esophageal microenvironment identifies signatures associated with progression of Barrett's esophagus | 2021 | Deshpande, N. P., Riordan, S. M., Gorman, C. J., Nielsen, S., Russell, T. L., Correa-Ospina, C., Fernando, B. S. M., Waters, S. A., Castano-Rodriguez, N., Man, S. M., Tedla, N., Wilkins, M. R. and Kaakoush, N. O. | Genome Medicine | http://dx.doi.org/10.1186/s13073-021-00951-6 |
| Limited clinical utility of lipid-laden macrophage index of induced sputum in predicting gastroesophageal reflux-related cough | 2021 | Dong, J., Huang, J., Liu, J., Tang, Y., Sivapalan, D., Lai, K., Zhong, N., Luo, W. and Chen, R. | Allergy, Asthma and Immunology Research | http://dx.doi.org/10.4168/AAIR.2021.13.5.799 |
| Salivary peptest for laryngopharyngeal reflux and gastroesophageal reflux disease: A systemic review and meta-analysis | 2021 | Guo, Z., Jiang, J., Wu, H., Zhu, J., Zhang, S. and Zhang, C. | Medicine | http://dx.doi.org/10.1097/MD.0000000000026756 |
| Evaluation of serum neuron specific enolase levels among patients with primary and secondary burning mouth syndrome | 2021 | Kishore, J., Shaikh, F., Zubairi, A. M., Mirza, S., Alqutub, M. N., Almubarak, A. M., Abduljabbar, T. and Vohra, F. | Cephalalgia. | http://dx.doi.org/10.1177/03331024211046613 |
| Gerd and its' therapeutic management by surgical methods | 2021 | Kumar, N., Mazumder, A. and Das, S. | International Journal of Pharmaceutical Research | http://dx.doi.org/10.31838/ijpr/2021.13.02.348 |
| Metabolomic profiling of extraesophageal reflux disease in children | 2021 | Mahoney, L. B., Esther, C. R., May, K. and Rosen, R. | Clinical and Translational Science | http://dx.doi.org/10.1111/cts.13064 |
| In vitro modelling of barrier impairment associated with gastro-oesophageal reflux disease (Gerd) | 2021 | Meloni, M., Buratti, P., Carriero, F. and Ceriotti, L. | Clinical and Experimental Gastroenterology | http://dx.doi.org/10.2147/CEG.S325346 |
| Association between Gastroesophageal Reflux Disease and Elastographic Parameters of Liver Steatosis and Fibrosis: Controlled Attenuation Parameter and Liver Stiffness Measurements | 2021 | Mikolasevic, I., Poropat, G., Filipec Kanizaj, T., Skenderevic, N., Zelic, M., Matasin, M., Vranic, L., Kresovic, A. and Hauser, G. | Canadian Journal of Gastroenterology and Hepatology | http://dx.doi.org/10.1155/2021/6670065 |
| Objective evidence of gastro-esophageal reflux disease is rare in patients with autoimmune gastritis | 2021 | Pilotto, V., Maddalo, G., Orlando, C., Fassan, M., Rugge, M., Farinati, F. and Savarino, E. | Journal of Gastrointestinal and Liver Diseases | http://dx.doi.org/10.15403/jgld-3033 |
| Comparative Cost Effectiveness of Reflux-Based and Reflux-Independent Strategies for Barrett's Esophagus Screening | 2021 | Sami, S. S., Moriarty, J. P., Rosedahl, J. K., Borah, B. J., Katzka, D. A., Wang, K. K., Kisiel, J. B., Ragunath, K., Rubenstein, J. H. and Iyer, P. G. | The American journal of gastroenterology | https://dx.doi.org/10.14309/ajg.0000000000001336 |
| Association of eosinophil-mediated inflammatory biomarkers with the presence of the Schatzki ring | 2021 | Sarbinowska, J., Wiatrak, B. and Wasko-Czopnik, D. | Advances in Medical Sciences | http://dx.doi.org/10.1016/j.advms.2021.05.004 |
| Sex differences in severe asthma: Results from severe asthma network in Italy-SANI | 2021 | Senna, G., Latorre, M., Bugiani, M., Caminati, M., Heffler, E., Morrone, D., Paoletti, G., Parronchi, P., Puggioni, F., Blasi, F., Canonica, G. W. and Paggiaro, P. | Allergy, Asthma and Immunology Research | https://dx.doi.org/10.4168/AAIR.2021.13.2.219 |
| Systemic Sclerosis in Zimbabwe: Autoantibody Biomarkers, Clinical, and Laboratory Correlates | 2021 | Sibanda, E. N., Dube, Y., Chakawa, M., Mduluza, T. and Mutapi, F. | Frontiers in Immunology | https://dx.doi.org/10.3389/fimmu.2021.679531 |
| Prenatal Ultrasound Measurement of Fetal Stomach Size Is Predictive of Postnatal Development of GERD in Isolated Cleft Lip and/or Palate | 2021 | Toscano, M., Burhans, K., Mack, L. M., Henderson, S., Koltz, P. F., Girotto, J. A. and Thornburg, L. L. | Cleft Palate-Craniofacial Journal | http://dx.doi.org/10.1177/1055665620968717 |
| Blood mRNA levels of T cells and IgE receptors are novel non-invasive biomarkers for eosinophilic esophagitis (EoE) | 2021 | Upparahalli Venkateshaiah, S., Rayapudi, M., Kandikattu, H. K., Yadavalli, C. S. and Mishra, A. | Clinical Immunology | http://dx.doi.org/10.1016/j.clim.2021.108752 |
| A urine and serum metabolomics study of gastroesophageal reflux disease in TCM syndrome differentiation using UPLC-Q-TOF/MS | 2021 | Ye, X., Wang, X., Wang, Y., Sun, W., Chen, Y., Wang, D. and Li, Z. | Journal of Pharmaceutical and Biomedical Analysis | http://dx.doi.org/10.1016/j.jpba.2021.114369 |
| JianpiQinghua granule reduced PPI dosage in patients with nonerosive reflux disease: A multicenter, randomized, double-blind, double-dummy, noninferiority study | 2021 | Zhang, J., Che, H., Zhang, B., Zhang, C., Zhou, B., Ji, H., Xie, J., Shi, X., Li, X., Wang, F. and Tang, X. | Phytomedicine | http://dx.doi.org/10.1016/j.phymed.2021.153584 |
| Identification of unique transcriptomic signatures and hub genes through rna sequencing and integrated wgcna and ppi network analysis in nonerosive reflux disease | 2021 | Zhao, Y., Ma, T. and Zou, D. | Journal of Inflammation Research | http://dx.doi.org/10.2147/JIR.S340452 |
| **Total Filtered: 150** | | | | |
