## Supplemental Table 3 for "Non-Invasive, MultiOmic and MultiCompartmental Biomarkers of Reflux Disease: A Systematic Review"

| **Supplemental Table 3. PUBMED Identified Manuscripts (N= 90)** | | | | |
| --- | --- | --- | --- | --- |
| **TITLE** | **YEAR** | **AUTHOR** | **JOURNAL** | **DOI** |
| Effect of Acid Suppression on Peripheral T-Lymphocyte Subsets and Immunohistochemical Esophageal Mucosal Changes in Patients With Gastroesophageal Reflux Disease | 2019 | Ahmed Osman, H., Aly, S. S., Mahmoud, H. S., Ahmed, E. H., Salah Eldin, E. M., Abdelrahim, E. A., El Masry, M. A., Herdan, R. A. and Hassan, M. H. | J Clin Gastroenterol | 10.1097/mcg.0000000000001098 |
| Chlorophyllin-stained macrophages as markers of pulmonary aspiration | 2013 | Alves, L. R., Soares, E. G., Aprile, L. R., Elias-Júnior, J., Vilas Boas, P. P. and Baddini-Martinez, J. | Am J Respir Crit Care Med | 10.1164/rccm.201307-1225LE |
| Does positioning affect tracheal aspiration of gastric content in ventilated infants? | 2015 | Aly, H., Soliman, R. M., El-Dib, M., Said, R. N., Abdellatif, M. A., Sibaii, H. and Elwakkad, A. | J Pediatr Gastroenterol Nutr | 10.1097/mpg.0000000000000601 |
| Mucosal impedance discriminates GERD from non-GERD conditions | 2015 | Ates, F., Yuksel, E. S., Higginbotham, T., Slaughter, J. C., Mabary, J., Kavitt, R. T., Garrett, C. G., Francis, D. and Vaezi, M. F. | Gastroenterology | 10.1053/j.gastro.2014.10.010 |
| A preliminary report on the use of Midodrine in treating refractory gastroesophageal disease: Randomized Double-Blind Controlled Trial | 2020 | Bagheri Lankarani, K., Sivandzadeh, G. R., Zare, M., Nejati, M., Niknam, R., Taghavi, A. R., Ejtehadi, F., Naini, M. A., Moini, M., Anbardar, M. H. and Peymani, P. | Acta Biomed | 10.23750/abm.v91i1.8486 |
| Relationship between disease characteristics and orofacial manifestations in systemic sclerosis: Canadian Systemic Sclerosis Oral Health Study III | 2015 | Baron, M., Hudson, M., Tatibouet, S., Steele, R., Lo, E., Gravel, S., Gyger, G., El Sayegh, T., Pope, J., Fontaine, A., Masetto, A., Matthews, D., Sutton, E., Thie, N., Jones, N., Copete, M., Kolbinson, D., Markland, J., Nogueira, G., Robinson, D., Fritzler, M. and Gornitsky, M. | Arthritis Care Res (Hoboken) | 10.1002/acr.22490 |
| Interleukin-6 as a potential indicator for prevention of high-risk adenoma recurrence by dietary flavonols in the polyp prevention trial | 2010 | Bobe, G., Albert, P. S., Sansbury, L. B., Lanza, E., Schatzkin, A., Colburn, N. H. and Cross, A. J. | Cancer Prev Res (Phila) | 10.1158/1940-6207.Capr-09-0161 |
| A pilot randomized clinical trial assessing the effect of cricoid pressure on risk of aspiration | 2018 | Bohman, J. K., Kashyap, R., Lee, A., He, Z., Soundar, S., Bolling, L. L. and Kor, D. J. | Clin Respir J | 10.1111/crj.12508 |
| Validation of Peptest™ in Patients with Gastro-Esophageal Reflux Disease and Laryngopharyngeal Reflux Undergoing Impedance Testing | 2019 | Bor, S., Capanoglu, D., Vardar, R., Woodcock, A. D., Fisher, J. and Dettmar, P. W. | J Gastrointestin Liver Dis | 10.15403/jgld-335 |
| Body mass index and comorbidities in adult severe asthmatics | 2014 | Bruno, A., Pace, E., Cibella, F. and Chanez, P. | Biomed Res Int | 10.1155/2014/607192 |
| Efficacy and safety of mavrilimumab in subjects with rheumatoid arthritis | 2013 | Burmester, G. R., Weinblatt, M. E., Mcinnes, I. B., Porter, D., Barbarash, O., Vatutin, M., Szombati, I., Esfandiari, E., Sleeman, M. A., Kane, C. D., Cavet, G., Wang, B., Godwood, A. and Magrini, F. | Ann Rheum Dis | 10.1136/annrheumdis-2012-202450 |
| Methylation panel is a diagnostic biomarker for Barrett's oesophagus in endoscopic biopsies and non-endoscopic cytology specimens | 2018 | Chettouh, H., Mowforth, O., Galeano-Dalmau, N., Bezawada, N., Ross-Innes, C., Macrae, S., Debiram-Beecham, I., O'donovan, M. and Fitzgerald, R. C. | Gut | 10.1136/gutjnl-2017-314026 |
| Evaluation of Barrett's esophagus with CK7, CK20, p53, Ki67, and COX2 expressions using chromoendoscopical examination | 2013 | Çoban, Ş., Örmeci, N., Savaş, B., Ekiz, F., Ensari, A., Kuzu, I. and Palabıyıkoğlu, M. | Dis Esophagus | 10.1111/j.1442-2050.2012.01352.x |
| Influence of proton pump inhibitor therapy on occurrence of voice prosthesis complications | 2020 | Danic Hadzibegovic, A., Kozmar, A., Hadzibegovic, I., Prgomet, D. and Danic, D. | Eur Arch Otorhinolaryngol | 10.1007/s00405-020-05784-4 |
| Impact of bile acids on the severity of laryngo-pharyngeal reflux | 2021 | De Corso, E., Baroni, S., Salonna, G., Marchese, M., Graziadio, M., Di Cintio, G., Paludetti, G., Costamagna, G. and Galli, J. | Clin Otolaryngol | 10.1111/coa.13643 |
| A single biopsy is valid for genetic diagnosis of eosinophilic esophagitis regardless of tissue preservation or location in the esophagus | 2015 | Dellon, E. S., Yellore, V., Andreatta, M. and Stover, J. | J Gastrointestin Liver Dis | 10.15403/jgld.2014.1121.242.bsy |
| Inflammatory and Comorbid Features of Patients with Severe Asthma and Frequent Exacerbations | 2017 | Denlinger, L. C., Phillips, B. R., Ramratnam, S., Ross, K., Bhakta, N. R., Cardet, J. C., Castro, M., Peters, S. P., Phipatanakul, W., Aujla, S., Bacharier, L. B., Bleecker, E. R., Comhair, S. A., Coverstone, A., Deboer, M., Erzurum, S. C., Fain, S. B., Fajt, M., Fitzpatrick, A. M., Gaffin, J., Gaston, B., Hastie, A. T., Hawkins, G. A., Holguin, F., Irani, A. M., Israel, E., Levy, B. D., Ly, N., Meyers, D. A., Moore, W. C., Myers, R., Opina, M. T., Peters, M. C., Schiebler, M. L., Sorkness, R. L., Teague, W. G., Wenzel, S. E., Woodruff, P. G., Mauger, D. T., Fahy, J. V. and Jarjour, N. N. | Am J Respir Crit Care Med | 10.1164/rccm.201602-0419OC |
| Cytosponge-trefoil factor 3 versus usual care to identify Barrett's oesophagus in a primary care setting: a multicentre, pragmatic, randomised controlled trial | 2020 | Fitzgerald, R. C., Di Pietro, M., O'donovan, M., Maroni, R., Muldrew, B., Debiram-Beecham, I., Gehrung, M., Offman, J., Tripathi, M., Smith, S. G., Aigret, B., Walter, F. M., Rubin, G. and Sasieni, P. | Lancet | 10.1016/s0140-6736(20)31099-0 |
| Exhaled breath condensate pH does not discriminate asymptomatic gastroesophageal reflux or the response to lansoprazole treatment in children with poorly controlled asthma | 2014 | Fitzpatrick, A. M., Holbrook, J. T., Wei, C. Y., Brown, M. S., Wise, R. A. and Teague, W. G. | J Allergy Clin Immunol Pract | 10.1016/j.jaip.2014.04.006 |
| Is histologic esophagitis associated with dental erosion: a cross-sectional observational study? | 2017 | Friesen, L. R., Bohaty, B., Onikul, R., Walker, M. P., Abraham, C., Williams, K. B., Cocjin, J. T., Cocjin, E. L. and Friesen, C. A. | BMC Oral Health | 10.1186/s12903-017-0408-z |
| Investigation of pretreatment prediction of proton pump inhibitor (PPI)-resistant patients with gastroesophageal reflux disease and the dose escalation challenge of PPIs-TORNADO study: a multicenter prospective study by the Acid-Related Symptom Research Group in Japan | 2011 | Furuta, T., Shimatani, T., Sugimoto, M., Ishihara, S., Fujiwara, Y., Kusano, M., Koike, T., Hongo, M., Chiba, T. and Kinoshita, Y. | J Gastroenterol | 10.1007/s00535-011-0446-2 |
| Association between tracheal pepsin, a reliable marker of gastric aspiration, and head of bed elevation among ventilated neonates | 2014 | Garland, J. S., Alex, C. P., Johnston, N., Yan, J. C. and Werlin, S. L. | J Neonatal Perinatal Med | 10.3233/npm-14814020 |
| Minor contribution of cytochrome P450 3A activity on fentanyl exposure in palliative care cancer patients | 2019 | Geist, M. J. P., Ziesenitz, V. C., Bardenheuer, H. J., Burhenne, J., Skopp, G. and Mikus, G. | Sci Rep | 10.1038/s41598-019-51279-6 |
| Aspiration and allograft injury secondary to gastroesophageal reflux occur in the immediate post-lung transplantation period (prospective clinical trial) | 2013 | Griffin, S. M., Robertson, A. G., Bredenoord, A. J., Brownlee, I. A., Stovold, R., Brodlie, M., Forrest, I., Dark, J. H., Pearson, J. P. and Ward, C. | Ann Surg | 10.1097/SLA.0b013e3182a6589b |
| Electroacupuncture to treat gastroesophageal reflux disease: study protocol for a randomized controlled trial | 2016 | Han, G., Leem, J., Lee, H. and Lee, J. | Trials | 10.1186/s13063-016-1371-8 |
| Randomised clinical trial to determine the safety of quercetin supplementation in patients with chronic obstructive pulmonary disease | 2020 | Han, M. K., Barreto, T. A., Martinez, F. J., Comstock, A. T. and Sajjan, U. S. | BMJ Open Respir Res | 10.1136/bmjresp-2018-000392 |
| Controlled exposure to particulate matter from urban street air is associated with decreased vasodilation and heart rate variability in overweight and older adults | 2015 | Hemmingsen, J. G., Rissler, J., Lykkesfeldt, J., Sallsten, G., Kristiansen, J., Møller, P. P. and Loft, S. | Part Fibre Toxicol | 10.1186/s12989-015-0081-9 |
| Effect of right lateral position with head elevation on tracheal aspirate pepsin in ventilated preterm neonates: randomized controlled trial | 2019 | Imam, S. S., Shinkar, D. M., Mohamed, N. A. and Mansour, H. E. | J Matern Fetal Neonatal Med | 10.1080/14767058.2018.1471674 |
| Impact of tracheal cuff shape on microaspiration of gastric contents in intubated critically ill patients: study protocol for a randomized controlled trial | 2015 | Jaillette, E., Brunin, G., Girault, C., Zerimech, F., Chiche, A., Broucqsault-Dedrie, C., Fayolle, C., Minacori, F., Alves, I., Barrailler, S., Robriquet, L., Tamion, F., Delaporte, E., Thellier, D., Delcourte, C., Duhamel, A. and Nseir, S. | Trials | 10.1186/s13063-015-0955-z |
| Impact of tapered-cuff tracheal tube on microaspiration of gastric contents in intubated critically ill patients: a multicenter cluster-randomized cross-over controlled trial | 2017 | Jaillette, E., Girault, C., Brunin, G., Zerimech, F., Behal, H., Chiche, A., Broucqsault-Dedrie, C., Fayolle, C., Minacori, F., Alves, I., Barrailler, S., Labreuche, J., Robriquet, L., Tamion, F., Delaporte, E., Thellier, D., Delcourte, C., Duhamel, A. and Nseir, S. | Intensive Care Med | 10.1007/s00134-017-4736-x |
| A Novel Susceptibility Locus Near GRIK2 Associated With Erosive Esophagitis in a Korean Cohort | 2020 | Jin, E. H., Park, B., Kim, Y. S., Choe, E. K., Choi, S. H., Kim, J. S. and Jung, S. A. | Clin Transl Gastroenterol | 10.14309/ctg.0000000000000145 |
| Disintegrin and metalloproteinases (ADAMs) expression in gastroesophageal reflux disease and in esophageal adenocarcinoma | 2017 | Kauttu, T., Mustonen, H., Vainionpää, S., Krogerus, L., Ilonen, I., Räsänen, J., Salo, J. and Puolakkainen, P. | Clin Transl Oncol | 10.1007/s12094-016-1503-3 |
| Biomarkers Predictive of Exacerbations in the SPIROMICS and COPDGene Cohorts | 2017 | Keene, J. D., Jacobson, S., Kechris, K., Kinney, G. L., Foreman, M. G., Doerschuk, C. M., Make, B. J., Curtis, J. L., Rennard, S. I., Barr, R. G., Bleecker, E. R., Kanner, R. E., Kleerup, E. C., Hansel, N. N., Woodruff, P. G., Han, M. K., Paine, R., 3rd, Martinez, F. J., Bowler, R. P. and O'neal, W. K. | Am J Respir Crit Care Med | 10.1164/rccm.201607-1330OC |
| Multilayered epithelium at the gastroesophageal junction is a marker of gastroesophageal reflux disease: data from a prospective Central European multicenter study (histoGERD trial) | 2014 | Langner, C., Wolf, E. M., Plieschnegger, W., Geppert, M., Wigginghaus, B., Höss, G. M., Eherer, A., Schneider, N. I., Rehak, P. and Vieth, M. | Virchows Arch | 10.1007/s00428-014-1550-5 |
| H2 receptor antagonists and right ventricular morphology: the MESA right ventricle study | 2014 | Leary, P. J., Barr, R. G., Bluemke, D. A., Bristow, M. R., Kronmal, R. A., Lima, J. A., Ralph, D. D., Ventetuolo, C. E. and Kawut, S. M. | Ann Am Thorac Soc | 10.1513/AnnalsATS.201407-344OC |
| Squamous tissue lymphocytes in the esophagus of controls and patients with reflux esophagitis and Barrett's esophagus are characterized by a non-inflammatory phenotype | 2014 | Lind, A., Koenderman, L., Kusters, J. G. and Siersema, P. D. | PLoS One | 10.1371/journal.pone.0106261 |
| Determinants of exhaled breath condensate pH in a large population with asthma | 2011 | Liu, L., Teague, W. G., Erzurum, S., Fitzpatrick, A., Mantri, S., Dweik, R. A., Bleecker, E. R., Meyers, D., Busse, W. W., Calhoun, W. J., Castro, M., Chung, K. F., Curran-Everett, D., Israel, E., Jarjour, W. N., Moore, W., Peters, S. P., Wenzel, S., Hunt, J. F. and Gaston, B. | Chest | 10.1378/chest.10-0163 |
| Measurement of mucosal conductivity by MII is a potential marker of mucosal integrity restored in infants on acid-suppression therapy | 2011 | Loots, C. M., Van Wijk, M. P., Smits, M. J., Wenzl, T. G., Benninga, M. A. and Omari, T. I. | J Pediatr Gastroenterol Nutr | 10.1097/MPG.0b013e318214c3cc |
| Hematological indices as indicators of silent inflammation in achalasia patients: A cross-sectional study | 2020 | López-Verdugo, F., Furuzawa-Carballeda, J., Romero-Hernández, F., Coss-Adame, E., Valdovinos, M. A., Priego-Ranero, A., Olvera-Prado, H., Narváez-Chavez, S., Peralta-Figueroa, J. and Torres-Villalobos, G. | Medicine (Baltimore) | 10.1097/md.0000000000019326 |
| Squamous Cellular Carcinoma Antigen Serum Determination as a Biomarker of Barrett Esophagus and Esophageal Cancer: A Phase III Study | 2018 | Maddalo, G., Fassan, M., Cardin, R., Piciocchi, M., Marafatto, F., Rugge, M., Zaninotto, G., Pozzan, C., Castoro, C., Ruol, A., Biasiolo, A. and Farinati, F. | J Clin Gastroenterol | 10.1097/mcg.0000000000000790 |
| Ectopic fat accumulation in patients with COPD: an ECLIPSE substudy | 2017 | Martin, M., Almeras, N., Després, J. P., Coxson, H. O., Washko, G. R., Vivodtzev, I., Wouters, E. F., Rutten, E., Williams, M. C., Murchison, J. T., Macnee, W., Sin, D. D. and Maltais, F. | Int J Chron Obstruct Pulmon Dis | 10.2147/copd.S124750 |
| Impact of self-reported gastroesophageal reflux disease in subjects from COPDGene cohort | 2014 | Martinez, C. H., Okajima, Y., Murray, S., Washko, G. R., Martinez, F. J., Silverman, E. K., Lee, J. H., Regan, E. A., Crapo, J. D., Curtis, J. L., Hatabu, H. and Han, M. K. | Respir Res | 10.1186/1465-9921-15-62 |
| Long-term follow-up of malignancy biomarkers in patients with Barrett's esophagus undergoing medical or surgical treatment | 2012 | Martinez De Haro, L. F., Ortiz, A., Parrilla, P., Munitiz, V., Martinez, C. M., Revilla, B., Ruiz De Angulo, D., Bermejo, J., Yélamos, J. and Molina, J. | Ann Surg | 10.1097/SLA.0b013e31824e6c6a |
| Serum albumin, total bilirubin, and patient age are independent confounders of hepatobiliary-phase gadoxetate parenchymal liver enhancement | 2019 | Matoori, S., Froehlich, J. M., Breitenstein, S., Pozdniakova, V., Reischauer, C., Kolokythas, O., Koh, D. M. and Gutzeit, A. | Eur Radiol | 10.1007/s00330-019-06179-8 |
| High yield reproducible rat model recapitulating human Barrett's carcinogenesis | 2017 | Matsui, D., Omstead, A. N., Kosovec, J. E., Komatsu, Y., Lloyd, E. J., Raphael, H., Kelly, R. J., Zaidi, A. H. and Jobe, B. A. | World J Gastroenterol | 10.3748/wjg.v23.i33.6077 |
| Clinical outcome of a randomized controlled blinded trial of open versus laparoscopic Nissen fundoplication in infants and children | 2011 | Mchoney, M., Wade, A. M., Eaton, S., Howard, R. F., Kiely, E. M., Drake, D. P., Curry, J. I. and Pierro, A. | Ann Surg | 10.1097/SLA.0b013e318226727f |
| Histology of symptomatic gastroesophageal reflux disease: is it predictive of response to proton pump inhibitors? | 2013 | Miwa, H., Takubo, K., Shimatani, T., Furuta, T., Oshima, T., Tanaka, J., Aida, J., Ito, M., Kurosawa, S., Joh, T., Wada, T., Habu, Y., Watanabe, Y., Hongo, M., Chiba, T. and Kinoshita, Y. | J Gastroenterol Hepatol | 10.1111/j.1440-1746.2012.07266.x |
| Randomized controlled trial comparing aerosolized swallowed fluticasone to esomeprazole for esophageal eosinophilia | 2013 | Moawad, F. J., Veerappan, G. R., Dias, J. A., Baker, T. P., Maydonovitch, C. L. and Wong, R. K. | Am J Gastroenterol | 10.1038/ajg.2012.443 |
| The effect of proton pump inhibitors on the CYP2C19 enzyme activity evaluated by the pantoprazole-(13)C breath test in GERD patients: clinical relevance for personalized medicine | 2016 | Modak, A. S., Klyarytska, I., Kriviy, V., Tsapyak, T. and Rabotyagova, Y. | J Breath Res | 10.1088/1752-7163/10/4/046017 |
| Response to therapy among neonates with gastro-esophageal reflux is associated with esophageal clearance | 2021 | Nobile, S., Meneghin, F., Marchionni, P., Noviello, C., Salvatore, S., Lista, G., Carnielli, V. P. and Vento, G. | Early Hum Dev | 10.1016/j.earlhumdev.2020.105248 |
| Barrett's oESophagus trial 3 (BEST3): study protocol for a randomised controlled trial comparing the Cytosponge-TFF3 test with usual care to facilitate the diagnosis of oesophageal pre-cancer in primary care patients with chronic acid reflux | 2018 | Offman, J., Muldrew, B., O'donovan, M., Debiram-Beecham, I., Pesola, F., Kaimi, I., Smith, S. G., Wilson, A., Khan, Z., Lao-Sirieix, P., Aigret, B., Walter, F. M., Rubin, G., Morris, S., Jackson, C., Sasieni, P. and Fitzgerald, R. C. | BMC Cancer | 10.1186/s12885-018-4664-3 |
| Treatment of non-erosive reflux disease and dynamics of the esophageal microbiome: a prospective multicenter study | 2020 | Park, C. H., Seo, S. I., Kim, J. S., Kang, S. H., Kim, B. J., Choi, Y. J., Byun, H. J., Yoon, J. H. and Lee, S. K. | Sci Rep | 10.1038/s41598-020-72082-8 |
| Distal mean nocturnal baseline impedance on pH-impedance monitoring predicts reflux burden and symptomatic outcome in gastro-oesophageal reflux disease | 2016 | Patel, A., Wang, D., Sainani, N., Sayuk, G. S. and Gyawali, C. P. | Aliment Pharmacol Ther | 10.1111/apt.13777 |
| Correlation of serum pepsinogens and gastrin-17 with atrophic gastritis in gastroesophageal reflux patients: a matched-pairs study | 2011 | Peitz, U., Wex, T., Vieth, M., Stolte, M., Willich, S., Labenz, J., Jaspersen, D., Lind, T. and Malfertheiner, P. | J Gastroenterol Hepatol | 10.1111/j.1440-1746.2010.06413.x |
| Objective Evidence of Gastro-Esophageal Reflux Disease is Rare in Patients with Autoimmune Gastritis | 2021 | Pilotto, V., Maddalo, G., Orlando, C., Fassan, M., Rugge, M., Farinati, F. and Savarino, E. V. | J Gastrointestin Liver Dis | 10.15403/jgld-3033 |
| Carditis: a relevant marker of gastroesophageal reflux disease. Data from a prospective central European multicenter study on histological and endoscopic diagnosis of esophagitis (histoGERD trial) | 2019 | Pinto, D., Plieschnegger, W., Schneider, N. I., Geppert, M., Bordel, H., Höss, G. M., Eherer, A., Wolf, E. M., Vieth, M. and Langner, C. | Dis Esophagus | 10.1093/dote/doy073 |
| Filaggrin and Periostin Expression Is Altered in Eosinophilic Esophagitis and Normalized With Treatment | 2017 | Politi, E., Angelakopoulou, A., Grapsa, D., Zande, M., Stefanaki, K., Panagiotou, I., Roma, E. and Syrigou, E. | J Pediatr Gastroenterol Nutr | 10.1097/mpg.0000000000001419 |
| Studies of salivary pepsin in patients with gastro-oesophageal reflux disease | 2019 | Race, C., Chowdry, J., Russell, J. M., Corfe, B. M. and Riley, S. A. | Aliment Pharmacol Ther | 10.1111/apt.15138 |
| A prospective evaluation of the effect of chronic proton pump inhibitor use on plasma biomarker levels in humans | 2012 | Raines, D., Chester, M., Diebold, A. E., Mamikunian, P., Anthony, C. T., Mamikunian, G. and Woltering, E. A. | Pancreas | 10.1097/MPA.0b013e318243a0b6 |
| Sustained benefit from intravenous immunoglobulin therapy for gastrointestinal involvement in systemic sclerosis | 2016 | Raja, J., Nihtyanova, S. I., Murray, C. D., Denton, C. P. and Ong, V. H. | Rheumatology (Oxford) | 10.1093/rheumatology/kev318 |
| Evaluation of a minimally invasive cell sampling device coupled with assessment of trefoil factor 3 expression for diagnosing Barrett's esophagus: a multi-center case-control study | 2015 | Ross-Innes, C. S., Debiram-Beecham, I., O'donovan, M., Walker, E., Varghese, S., Lao-Sirieix, P., Lovat, L., Griffin, M., Ragunath, K., Haidry, R., Sami, S. S., Kaye, P., Novelli, M., Disep, B., Ostler, R., Aigret, B., North, B. V., Bhandari, P., Haycock, A., Morris, D., Attwood, S., Dhar, A., Rees, C., Rutter, M. D., Sasieni, P. D. and Fitzgerald, R. C. | PLoS Med | 10.1371/journal.pmed.1001780 |
| Use of direct, endoscopic-guided measurements of mucosal impedance in diagnosis of gastroesophageal reflux disease | 2012 | Saritas Yuksel, E., Higginbotham, T., Slaughter, J. C., Mabary, J., Kavitt, R. T., Garrett, C. G. and Vaezi, M. F. | Clin Gastroenterol Hepatol | 10.1016/j.cgh.2012.05.018 |
| Rapid salivary pepsin test: blinded assessment of test performance in gastroesophageal reflux disease | 2012 | Saritas Yuksel, E., Hong, S. K., Strugala, V., Slaughter, J. C., Goutte, M., Garrett, C. G., Dettmar, P. W. and Vaezi, M. F. | Laryngoscope | 10.1002/lary.23252 |
| Use of the PEPTEST™ tool for the diagnosis of GERD in the Emergency Department | 2019 | Saviano, A., Petruzziello, C., Brigida, M., Tersigni, I., Migneco, A., Piccioni, A., Saviano, L., Covino, M., Franceschi, F. and Ojetti, V. | Am J Emerg Med | 10.1016/j.ajem.2019.06.047 |
| Predisposing factors for positive D-Xylose breath test for evaluation of small intestinal bacterial overgrowth: a retrospective study of 932 patients | 2015 | Schatz, R. A., Zhang, Q., Lodhia, N., Shuster, J., Toskes, P. P. and Moshiree, B. | World J Gastroenterol | 10.3748/wjg.v21.i15.4574 |
| A prospective randomized study of systemic inflammation and immune response after laparoscopic nissen fundoplication performed with standard and low-pressure pneumoperitoneum | 2013 | Schietroma, M., Carlei, F., Cecilia, E. M., Piccione, F., Sista, F., De Vita, F. and Amicucci, G. | Surg Laparosc Endosc Percutan Tech | 10.1097/SLE.0b013e3182827e51 |
| Vorinostat in refractory soft tissue sarcomas - Results of a multi-centre phase II trial of the German Soft Tissue Sarcoma and Bone Tumour Working Group (AIO) | 2016 | Schmitt, T., Mayer-Steinacker, R., Mayer, F., Grünwald, V., Schütte, J., Hartmann, J. T., Kasper, B., Hüsing, J., Hajda, J., Ottawa, G., Mechtersheimer, G., Mikus, G., Burhenne, J., Lehmann, L., Heilig, C. E., Ho, A. D. and Egerer, G. | Eur J Cancer | 10.1016/j.ejca.2016.05.018 |
| Capsaicin and evodiamine ingestion does not augment energy expenditure and fat oxidation at rest or after moderately-intense exercise | 2013 | Schwarz, N. A., Spillane, M., La Bounty, P., Grandjean, P. W., Leutholtz, B. and Willoughby, D. S. | Nutr Res | 10.1016/j.nutres.2013.08.007 |
| Gastrointestinal symptoms in idiopathic pulmonary fibrosis patients treated with pirfenidone and herbal medicine | 2014 | Shimizu, Y., Shimoyama, Y., Kawada, A., Kusano, M., Hosomi, Y., Sekiguchi, M., Kawata, T., Horie, T., Ishii, Y., Yamada, M., Dobashi, K. and Takise, A. | J Biol Regul Homeost Agents |  |
| Polymorphisms of Genes Related to Function and Metabolism of Vitamin D in Esophageal Adenocarcinoma | 2019 | Singhal, S., Kapoor, H., Subramanian, S., Agrawal, D. K. and Mittal, S. K. | J Gastrointest Cancer | 10.1007/s12029-018-0164-6 |
| Gastric adenocarcinoma with chief cell differentiation: a proposal for reclassification as oxyntic gland polyp/adenoma | 2012 | Singhi, A. D., Lazenby, A. J. and Montgomery, E. A. | Am J Surg Pathol | 10.1097/PAS.0b013e31825033e7 |
| Is electrical brain activity a reliable biomarker for opioid analgesia in the gut? | 2011 | Staahl, C., Krarup, A. L., Olesen, A. E., Brock, C., Graversen, C. and Drewes, A. M. | Basic Clin Pharmacol Toxicol | 10.1111/j.1742-7843.2011.00727.x |
| Prevalence, characteristics and outcome of non-cardiac chest pain and elevated copeptin levels | 2014 | Stallone, F., Twerenbold, R., Wildi, K., Reichlin, T., Rubini Gimenez, M., Haaf, P., Fuechslin, N., Hillinger, P., Jaeger, C., Kreutzinger, P., Puelacher, C., Radosavac, M., Moreno Weidmann, Z., Moehring, B., Honegger, U., Schumacher, C., Denhaerynck, K., Arnold, C., Bingisser, R., Vollert, J. O., Osswald, S. and Mueller, C. | Heart | 10.1136/heartjnl-2014-305583 |
| A prospective cohort study on overweight, smoking, alcohol consumption, and risk of Barrett's esophagus | 2011 | Steevens, J., Schouten, L. J., Driessen, A. L., Huysentruyt, C. J., Keulemans, Y. C., Goldbohm, R. A. and Van Den Brandt, P. A. | Cancer Epidemiol Biomarkers Prev | 10.1158/1055-9965.Epi-10-0636 |
| Improved Cough and Cough-Specific Quality of Life in Patients Treated for Scleroderma-Related Interstitial Lung Disease: Results of Scleroderma Lung Study II | 2017 | Tashkin, D. P., Volkmann, E. R., Tseng, C. H., Roth, M. D., Khanna, D., Furst, D. E., Clements, P. J., Theodore, A., Kafaja, S., Kim, G. H., Goldin, J., Ariolla, E. and Elashoff, R. M. | Chest | 10.1016/j.chest.2016.11.052 |
| Changes in obesity-related diseases and biochemical variables after laparoscopic sleeve gastrectomy: a two-year follow-up study | 2014 | Våge, V., Sande, V. A., Mellgren, G., Laukeland, C., Behme, J. and Andersen, J. R. | BMC Surg | 10.1186/1471-2482-14-8 |
| Iron deficiency in worsening heart failure is associated with reduced estimated protein intake, fluid retention, inflammation, and antiplatelet use | 2019 | Van Der Wal, H. H., Grote Beverborg, N., Dickstein, K., Anker, S. D., Lang, C. C., Ng, L. L., Van Veldhuisen, D. J., Voors, A. A. and Van Der Meer, P. | Eur Heart J | 10.1093/eurheartj/ehz680 |
| Clinical symptom tool that raises the index of suspicion for eosinophilic oesophagitis in adults and drives earlier biopsy for definitive diagnosis | 2017 | Von Arnim, U., Röhl, F. W., Miehlke, S., Jechorek, D., Reinhold, D., Wex, T. and Malfertheiner, P. | Aliment Pharmacol Ther | 10.1111/apt.13869 |
| Dose-response effect of Bifidobacterium lactis HN019 on whole gut transit time and functional gastrointestinal symptoms in adults | 2011 | Waller, P. A., Gopal, P. K., Leyer, G. J., Ouwehand, A. C., Reifer, C., Stewart, M. E. and Miller, L. E. | Scand J Gastroenterol | 10.3109/00365521.2011.584895 |
| Validation in China of a non-invasive salivary pepsin biomarker containing two unique human pepsin monoclonal antibodies to diagnose gastroesophageal reflux disease | 2019 | Wang, Y. F., Yang, C. Q., Chen, Y. X., Cao, A. P., Yu, X. F., Yu, Y., Zhang, Z. Y., Shen, X. Z., Liu, F., Zhong, L., Wang, Y. X., Liu, Z. J., Shi, Y. H., Zhong, J., Li, J. N., Lan, Y., Lenham, R. K., Woodcock, A. D., Dettmar, P. W. and Fang, J. Y. | J Dig Dis | 10.1111/1751-2980.12783 |
| Two Years Remission of Type 2 Diabetes Mellitus after Bariatric Surgery | 2019 | Wazir, N., Arshad, M. F., Finney, J., Kirk, K. and Dewan, S. | J Coll Physicians Surg Pak | 10.29271/jcpsp.2019.10.967 |
| Leukocyte telomere length in relation to the risk of Barrett's esophagus and esophageal adenocarcinoma | 2016 | Wennerström, E. C., Risques, R. A., Prunkard, D., Giffen, C., Corley, D. A., Murray, L. J., Whiteman, D. C., Wu, A. H., Bernstein, L., Ye, W., Chow, W. H., Vaughan, T. L. and Liao, L. M. | Cancer Med | 10.1002/cam4.810 |
| Reflux-associated oxygen desaturations: usefulness in diagnosing reflux-related respiratory symptoms | 2013 | Wilshire, C. L., Salvador, R., Sepesi, B., Niebisch, S., Watson, T. J., Litle, V. R., Peyre, C. G., Jones, C. E. and Peters, J. H. | J Gastrointest Surg | 10.1007/s11605-012-2065-5 |
| Exercise and the Prevention of Oesophageal Cancer (EPOC) study protocol: a randomized controlled trial of exercise versus stretching in males with Barrett's oesophagus | 2010 | Winzer, B. M., Paratz, J. D., Reeves, M. M. and Whiteman, D. C. | BMC Cancer | 10.1186/1471-2407-10-292 |
| Superficial Esophageal Mucosal Afferent Nerves May Contribute to Reflux Hypersensitivity in Nonerosive Reflux Disease | 2017 | Woodland, P., Shen Ooi, J. L., Grassi, F., Nikaki, K., Lee, C., Evans, J. A., Koukias, N., Triantos, C., Mcdonald, S. A., Peiris, M., Aktar, R., Blackshaw, L. A. and Sifrim, D. | Gastroenterology | 10.1053/j.gastro.2017.07.017 |
| Lymphocytic Esophagitis With CD4 T-cell-predominant Intraepithelial Lymphocytes and Primary Esophageal Motility Abnormalities: A Potential Novel Clinicopathologic Entity | 2015 | Xue, Y., Suriawinata, A., Liu, X., Li, Z., Gabbard, S., Rothstein, R., Lacy, B. and Lisovsky, M. | Am J Surg Pathol | 10.1097/pas.0000000000000493 |
| Shar Pei Larynx: Supraglottic and Postcricoid Mucosal Redundancy and Its Association With Medical Comorbidities | 2019 | Yiu, Y., Tibbetts, K. M., Simpson, C. B. and Matrka, L. A. | Ann Otol Rhinol Laryngol | 10.1177/0003489418810893 |
| The Association of Depressive Symptoms With Rates of Acute Exacerbations in Patients With COPD: Results From a 3-year Longitudinal Follow-up of the ECLIPSE Cohort | 2017 | Yohannes, A. M., Mülerová, H., Lavoie, K., Vestbo, J., Rennard, S. I., Wouters, E. and Hanania, N. A. | J Am Med Dir Assoc | 10.1016/j.jamda.2017.05.024 |
| Targeting the COX1/2-Driven thromboxane A2 pathway suppresses Barrett's esophagus and esophageal adenocarcinoma development | 2019 | Zhang, T., Wang, Q., Ma, W. Y., Wang, K., Chang, X., Johnson, M. L., Bai, R., Bode, A. M., Foster, N. R., Falk, G. W., Limburg, P. J., Iyer, P. G. and Dong, Z. | EBioMedicine | 10.1016/j.ebiom.2019.10.038 |
| Oesophageal intrasquamous IgG4 deposits: an adjunctive marker to distinguish eosinophilic oesophagitis from reflux oesophagitis | 2016 | Zukerberg, L., Mahadevan, K., Selig, M. and Deshpande, V. | Histopathology | 10.1111/his.12892 |
| **Total Filtered: 90** | | | | |
