## Supplemental Table 5 for "Non-Invasive, MultiOmic and MultiCompartmental Biomarkers of Reflux Disease: A Systematic Review"

| **Supplemental Table 5. Excluded Manuscripts (N=208)** | | | | | | |
| --- | --- | --- | --- | --- | --- | --- |
| **Reasons for Exclusion** | **TITLE** | **YEAR** | **AUTHOR** | **JOURNAL** | **DOI** | **DATABASE** |
| Unrelated | Chlorophyllin-stained macrophages as markers of pulmonary aspiration | 2013 | Alves, L. R., Soares, E. G., Aprile, L. R., Elias-Júnior, J., Vilas Boas, P. P. and Baddini-Martinez, J. | Am J Respir Crit Care Med | 10.1164/rccm.201307-1225LEA | PubMed |
| Unrelated | Mucosal impedance discriminates GERD from non-GERD conditions | 2015 | Ates, F., Yuksel, E. S., Higginbotham, T., Slaughter, J. C., Mabary, J., Kavitt, R. T., Garrett, C. G., Francis, D. and Vaezi, M. F. | Gastroenterology | 10.1053/j.gastro.2014.10.010 | PubMed |
| Unrelated | A preliminary report on the use of Midodrine in treating refractory gastroesophageal disease: Randomized Double-Blind Controlled Trial | 2020 | Bagheri Lankarani, K., Sivandzadeh, G. R., Zare, M., Nejati, M., Niknam, R., Taghavi, A. R., Ejtehadi, F., Naini, M. A., Moini, M., Anbardar, M. H. and Peymani, P. | Acta Biomed | 10.23750/abm.v91i1.8486 | PubMed |
| Unrelated | Prevalence and predictors of columnar lined esophagus in Gastroesophageal Reflux Disease (GERD) patients undergoing upper endoscopy | 2012 | Balasubramanian, G., Singh, M., Gupta, N., Gaddam, S., Giacchino, M., Wani, S. B., Moloney, B., Higbee, A. D., Rastogi, A., Bansal, A. and Sharma, P. | American Journal of Gastroenterology | http://dx.doi.org/10.1038/ajg.2012.299 | Embase |
| Unrelated | Relationship between disease characteristics and orofacial manifestations in systemic sclerosis: Canadian Systemic Sclerosis Oral Health Study III | 2015 | Baron, M., Hudson, M., Tatibouet, S., Steele, R., Lo, E., Gravel, S., Gyger, G., El Sayegh, T., Pope, J., Fontaine, A., Masetto, A., Matthews, D., Sutton, E., Thie, N., Jones, N., Copete, M., Kolbinson, D., Markland, J., Nogueira, G., Robinson, D., Fritzler, M. and Gornitsky, M. | Arthritis Care Res (Hoboken) | 10.1002/acr.22490 | PubMed |
| Unrelated | Exhaled Breath Condensate pH in Lung Cancer, the Impact of Clinical Factors | 2015 | Bikov, A., Lazar, Z., Gyulai, N., Szentkereszty, M., Losonczy, G., Horvath, I. and Galffy, G. | Lung | http://dx.doi.org/10.1007/s00408-015-9778-7 | Embase |
| Unrelated | A striking local esophageal cytokine expression profile in eosinophilic esophagitis | 2011 | Blanchard, C., Stucke, E. M., Rodriguez-Jimenez, B., Burwinkel, K., Collins, M. H., Ahrens, A., Alexander, E. S., Buckmeier Butz, B. K., Jameson, S. C., Kaul, A., Franciosi, J. P., Kushner, J. P., Putnam, P. E., Abonia, J. P. and Rothenberg, M. E. | Journal of Allergy and Clinical Immunology | http://dx.doi.org/10.1016/j.jaci.2010.10.039 | Embase |
| Unrelated | Interleukin-6 as a potential indicator for prevention of high-risk adenoma recurrence by dietary flavonols in the polyp prevention trial | 2010 | Bobe, G., Albert, P. S., Sansbury, L. B., Lanza, E., Schatzkin, A., Colburn, N. H. and Cross, A. J. | Cancer Prev Res (Phila) | 10.1158/1940-6207.Capr-09-0161 | PubMed |
| Unrelated | Body mass index and comorbidities in adult severe asthmatics | 2014 | Bruno, A., Pace, E., Cibella, F. and Chanez, P. | Biomed Res Int | 10.1155/2014/607192 | PubMed |
| Unrelated | Candidate serum metabolite biomarkers for differentiating gastroesophageal reflux disease, Barrett's esophagus, and high-grade dysplasia/esophageal adenocarcinoma | 2017 | Buas, M. F., Gu, H., Djukovic, D., Zhu, J., Onstad, L., Reid, B. J., Raftery, D. and Vaughan, T. L. | Metabolomics | http://dx.doi.org/10.1007/s11306-016-1154-y | Embase |
| Unrelated | Risk factors associated with the development of interstitial lung abnormalities | 2021 | Buendia-Roldan, I., Fernandez, R., Mejia, M., Juarez, F., Ramirez-Martinez, G., Montes, E., Pruneda, A. K. S., Martinez-Espinosa, K., Alarcon-Dionet, A., Herrera, I., Becerril, C., Chavez-Galan, L., Preciado, M., Pardo, A. and Selman, M. | European Respiratory Journal | http://dx.doi.org/10.1183/13993003.03005-2020 | Embase |
| Unrelated | Efficacy and safety of mavrilimumab in subjects with rheumatoid arthritis | 2013 | Burmester, G. R., Weinblatt, M. E., Mcinnes, I. B., Porter, D., Barbarash, O., Vatutin, M., Szombati, I., Esfandiari, E., Sleeman, M. A., Kane, C. D., Cavet, G., Wang, B., Godwood, A. and Magrini, F. | Ann Rheum Dis | 10.1136/annrheumdis-2012-202450 | PubMed |
| Unrelated | Central obesity and other factors associated with uncontrolled asthma in women | 2015 | Capelo, A. V., De Fonseca, V. M., Peixoto, M. V. M., De Carvalho, S. R. and Guerino, L. G. | Allergy, Asthma and Clinical Immunology | http://dx.doi.org/10.1186/s13223-015-0076-y | Embase |
| Unrelated | OPCML is hypermethylated in a subset of patients with metaplastic changes in their esophagus | 2018 | Castano-Rodriguez, N., Popple, G. L., Porras-Hurtado, G. L., Cardona-Deazza, J. L., Montoya-Martinez, J. J., Cadavid-Velez, A. J., Toro-Hidalgo, H. W., Cobo-Alvarado, A. R., Del Socorro Hincapie-Rincon, O., Riordan, S. M. and Kaakoush, N. O. | Biomarker Research | http://dx.doi.org/10.1186/s40364-018-0150-y | Embase |
| Unrelated | The association between depression and gastroesophageal reflux based on phylogenetic analysis of mirna biomarkers | 2020 | Chen, Y. H. and Wang, H. | Current Medicinal Chemistry | http://dx.doi.org/10.2174/0929867327666200425214906 | Embase |
| Unrelated | Functional dyspepsia susceptibility is associated with TGFB1 gene polymorphisms (RS4803455, RS1800469) in H pylori-negative Chinese population | 2019 | Cheung, C. K. Y., Lan, L. L., Chan, Y., Yuen, K., Cheong, P. K., Fang, F. and Wu, J. C. Y. | Neurogastroenterology and Motility | http://dx.doi.org/10.1111/nmo.13681 | Embase |
| Unrelated | Plasma levels of TNF-alpha, IL-6, IFN-gamma, IL-12, IL-17, IL-22, and IL-23 in achalasia, eosinophilic esophagitis (EoE), and gastroesophageal reflux disease (GERD) | 2019 | Clayton, S., Cauble, E., Kumar, A., Patil, N., Ledford, D., Kolliputi, N., Lopes-Virella, M. F., Castell, D. and Richter, J. | BMC Gastroenterology | http://dx.doi.org/10.1186/s12876-019-0937-9 | Embase |
| Unrelated | Eosinophilic Esophagitis Histology Remission Score: Significant Relations to Measures of Disease Activity and Symptoms | 2020 | Collins, M. H., Martin, L. J., Wen, T., Abonia, J. P., Putnam, P. E., Mukkada, V. A. and Rothenberg, M. E. | Journal of pediatric gastroenterology and nutrition | http://dx.doi.org/10.1097/MPG.0000000000002637 | Embase |
| Unrelated | Influence of proton pump inhibitor therapy on occurrence of voice prosthesis complications | 2020 | Danic Hadzibegovic, A., Kozmar, A., Hadzibegovic, I., Prgomet, D. and Danic, D. | Eur Arch Otorhinolaryngol | 10.1007/s00405-020-05784-4 | PubMed |
| Unrelated | Diagnostic utility of major basic protein, eotaxin-3, and leukotriene enzyme staining in eosinophilic esophagitis | 2012 | Dellon, E. S., Chen, X., Miller, C. R., Woosley, J. T. and Shaheen, N. J. | American Journal of Gastroenterology | http://dx.doi.org/10.1038/ajg.2012.202 | Embase |
| Unrelated | Prospective assessment of serum periostin as a biomarker for diagnosis and monitoring of eosinophilic oesophagitis | 2016 | Dellon, E. S., Higgins, L. L., Beitia, R., Rusin, S., Woosley, J. T., Veerappan, R., Selitsky, S. R., Parker, J. S., Genta, R. M., Lash, R. H., Aranda, R., Peach, R. J. and Grimm, M. | Alimentary Pharmacology and Therapeutics | http://dx.doi.org/10.1111/apt.13672 | Embase |
| Unrelated | Utility of a noninvasive serum biomarker panel for diagnosis and monitoring of eosinophilic esophagitis: A prospective study | 2015 | Dellon, E. S., Rusin, S., Gebhart, J. H., Covey, S., Higgins, L. L., Beitia, R., Speck, O., Woodward, K., Woosley, J. T. and Shaheen, N. J. | American Journal of Gastroenterology | http://dx.doi.org/10.1038/ajg.2015.57 | Embase |
| Unrelated | A single biopsy is valid for genetic diagnosis of eosinophilic esophagitis regardless of tissue preservation or location in the esophagus | 2015 | Dellon, E. S., Yellore, V., Andreatta, M. and Stover, J. | J Gastrointestin Liver Dis | 10.15403/jgld.2014.1121.242.bsy | PubMed |
| Unrelated | Inflammatory and Comorbid Features of Patients with Severe Asthma and Frequent Exacerbations | 2017 | Denlinger, L. C., Phillips, B. R., Ramratnam, S., Ross, K., Bhakta, N. R., Cardet, J. C., Castro, M., Peters, S. P., Phipatanakul, W., Aujla, S., Bacharier, L. B., Bleecker, E. R., Comhair, S. A., Coverstone, A., Deboer, M., Erzurum, S. C., Fain, S. B., Fajt, M., Fitzpatrick, A. M., Gaffin, J., Gaston, B., Hastie, A. T., Hawkins, G. A., Holguin, F., Irani, A. M., Israel, E., Levy, B. D., Ly, N., Meyers, D. A., Moore, W. C., Myers, R., Opina, M. T., Peters, M. C., Schiebler, M. L., Sorkness, R. L., Teague, W. G., Wenzel, S. E., Woodruff, P. G., Mauger, D. T., Fahy, J. V. and Jarjour, N. N. | Am J Respir Crit Care Med | 10.1164/rccm.201602-0419OC | PubMed |
| Unrelated | Multi-omics of the esophageal microenvironment identifies signatures associated with progression of Barrett's esophagus | 2021 | Deshpande, N. P., Riordan, S. M., Gorman, C. J., Nielsen, S., Russell, T. L., Correa-Ospina, C., Fernando, B. S. M., Waters, S. A., Castano-Rodriguez, N., Man, S. M., Tedla, N., Wilkins, M. R. and Kaakoush, N. O. | Genome Medicine | http://dx.doi.org/10.1186/s13073-021-00951-6 | Embase |
| Unrelated | Limited clinical utility of lipid-laden macrophage index of induced sputum in predicting gastroesophageal reflux-related cough | 2021 | Dong, J., Huang, J., Liu, J., Tang, Y., Sivapalan, D., Lai, K., Zhong, N., Luo, W. and Chen, R. | Allergy, Asthma and Immunology Research | http://dx.doi.org/10.4168/AAIR.2021.13.5.799 | Embase |
| Unrelated | Cytosponge-trefoil factor 3 versus usual care to identify Barrett's oesophagus in a primary care setting: a multicentre, pragmatic, randomised controlled trial | 2020 | Fitzgerald, R. C., Di Pietro, M., O'donovan, M., Maroni, R., Muldrew, B., Debiram-Beecham, I., Gehrung, M., Offman, J., Tripathi, M., Smith, S. G., Aigret, B., Walter, F. M., Rubin, G. and Sasieni, P. | Lancet | 10.1016/s0140-6736(20)31099-0 | PubMed |
| Unrelated | The oesophageal string test: A novel, minimally invasive method measures mucosal inflammation in eosinophilic oesophagitis | 2013 | Furuta, G. T., Kagalwalla, A. F., Lee, J. J., Alumkal, P., Maybruck, B. T., Fillon, S., Masterson, J. C., Ochkur, S., Protheroe, C., Moore, W., Pan, Z., Amsden, K., Robinson, Z., Capocelli, K., Mukkada, V., Atkins, D., Fleischer, D., Hosford, L., Kwatia, M. A., Schroeder, S., Kelly, C., Lovell, M., Melin-Aldana, H. and Ackerman, S. J. | Gut | http://dx.doi.org/10.1136/gutjnl-2012-303171 | Embase |
| Unrelated | Investigation of pretreatment prediction of proton pump inhibitor (PPI)-resistant patients with gastroesophageal reflux disease and the dose escalation challenge of PPIs-TORNADO study: a multicenter prospective study by the Acid-Related Symptom Research Group in Japan | 2011 | Furuta, T., Shimatani, T., Sugimoto, M., Ishihara, S., Fujiwara, Y., Kusano, M., Koike, T., Hongo, M., Chiba, T. and Kinoshita, Y. | J Gastroenterol | 10.1007/s00535-011-0446-2 | PubMed |
| Unrelated | Minor contribution of cytochrome P450 3A activity on fentanyl exposure in palliative care cancer patients | 2019 | Geist, M. J. P., Ziesenitz, V. C., Bardenheuer, H. J., Burhenne, J., Skopp, G. and Mikus, G. | Sci Rep | 10.1038/s41598-019-51279-6 | PubMed |
| Unrelated | Assessment of cricopharyngeal muscle aging with apoptotic markers | 2014 | Gor, A. P., Alkan, Z., Yigit, O., Dursun, N., Engin, A., Akin, S. M. and Sam, B. | The Journal of craniofacial surgery | http://dx.doi.org/10.1097/SCS.0000000000000940 | Embase |
| Unrelated | Cost-effectiveness of endoscopic surveillance of non-dysplastic Barrett's esophagus | 2014 | Gordon, L. G., Mayne, G. C., Hirst, N. G., Bright, T., Whiteman, D. C. and Watson, D. I. | Gastrointestinal Endoscopy | http://dx.doi.org/10.1016/j.gie.2013.07.046 | Embase |
| Unrelated | Interchangeable Use of Proton Pump Inhibitors Based on Relative Potency | 2018 | Graham, D. Y. and Tansel, A. | Clinical Gastroenterology and Hepatology | http://dx.doi.org/10.1016/j.cgh.2017.09.033 | Embase |
| Unrelated | Aspiration and allograft injury secondary to gastroesophageal reflux occur in the immediate post-lung transplantation period (Prospective Clinical Trial) | 2013 | Griffin, S. M., Robertson, A. G. N., Bredenoord, A. J., Brownlee, I. A., Stovold, R., Brodlie, M., Forrest, I., Dark, J. H., Pearson, J. P. and Ward, C. | Annals of Surgery | http://dx.doi.org/10.1097/SLA.0b013e3182a6589b | Embase |
| Unrelated | Developing a blood-based gene mutation assay as a novel biomarker for oesophageal adenocarcinoma | 2019 | Haboubi, H. N., Lawrence, R. L., Rees, B., Williams, L., Manson, J. M., Al-Mossawi, N., Bodger, O., Griffiths, P., Thornton, C. and Jenkins, G. J. | Scientific reports | http://dx.doi.org/10.1038/s41598-019-41490-w | Embase |
| Unrelated | Identification of Individuals with Functional Dyspepsia With a Simple, Minimally Invasive Test: A Single Center Cohort Study of the Oral Capsaicin Test | 2018 | Hammer, J. | American Journal of Gastroenterology | http://dx.doi.org/10.1038/ajg.2018.16 | Embase |
| Unrelated | Randomised clinical trial to determine the safety of quercetin supplementation in patients with chronic obstructive pulmonary disease | 2020 | Han, M. K., Barreto, T. A., Martinez, F. J., Comstock, A. T. and Sajjan, U. S. | BMJ Open Respir Res | 10.1136/bmjresp-2018-000392 | PubMed |
| Unrelated | Controlled exposure to particulate matter from urban street air is associated with decreased vasodilation and heart rate variability in overweight and older adults | 2015 | Hemmingsen, J. G., Rissler, J., Lykkesfeldt, J., Sallsten, G., Kristiansen, J., Møller, P. P. and Loft, S. | Part Fibre Toxicol | 10.1186/s12989-015-0081-9 | PubMed |
| Unrelated | Urease and Helicobacter spp. Antigens in Pulmonary Granuloma | 2013 | Herndon, B., Quinn, T., Wasson, N., Nzabi, M. and Molteni, A. | Journal of Comparative Pathology | http://dx.doi.org/10.1016/j.jcpa.2012.06.011 | Embase |
| Unrelated | Promising Modalities to Identify and Monitor Eosinophilic Esophagitis | 2017 | Hiremath, G. and Gupta, S. K. | Clinical Gastroenterology and Hepatology | http://dx.doi.org/10.1016/j.cgh.2017.05.004 | Embase |
| Unrelated | Oil red O stain of alveolar macrophages is an effective screening test for gastroesophageal reflux disease in lung transplant recipients | 2010 | Hopkins, P. M., Kermeen, F., Duhig, E., Fletcher, L., Gradwell, J., Whitfield, L., Godinez, C., Musk, M., Chambers, D., Gotley, D. and Mcneil, K. | Journal of Heart and Lung Transplantation | http://dx.doi.org/10.1016/j.healun.2010.03.015 | Embase |
| Unrelated | Correlation between the serum pepsinogen I level and the symptom degree in proton pump inhibitor-users administered with a probiotic | 2014 | Igarashi, M., Nagano, J., Tsuda, A., Suzuki, T., Koike, J., Uchida, T., Matsushima, M., Mine, T. and Koga, Y. | Pharmaceuticals | http://dx.doi.org/10.3390/ph7070754 | Embase |
| Unrelated | Impact of tapered-cuff tracheal tube on microaspiration of gastric contents in intubated critically ill patients: a multicenter cluster-randomized cross-over controlled trial | 2017 | Jaillette, E., Girault, C., Brunin, G., Zerimech, F., Behal, H., Chiche, A., Broucqsault-Dedrie, C., Fayolle, C., Minacori, F., Alves, I., Barrailler, S., Labreuche, J., Robriquet, L., Tamion, F., Delaporte, E., Thellier, D., Delcourte, C., Duhamel, A. and Nseir, S. | Intensive Care Med | 10.1007/s00134-017-4736-x | PubMed |
| Unrelated | Fragments of e-Cadherin as Biomarkers of Non-erosive Reflux Disease | 2018 | Jovov, B., Reed, C. C., Shaheen, N. J., Pruitt, A., Ferrell, K., Orlando, G. S., Djukic, Z. and Orlando, R. C. | Digestive Diseases and Sciences | http://dx.doi.org/10.1007/s10620-017-4815-4 | Embase |
| Unrelated | Eosinophilic esophagitis: Epithelial mesenchymal transition contributes to esophageal remodeling and reverses with treatment | 2012 | Kagalwalla, A. F., Akhtar, N., Woodruff, S. A., Rea, B. A., Masterson, J. C., Mukkada, V., Parashette, K. R., Du, J., Fillon, S., Protheroe, C. A., Lee, J. J., Amsden, K., Melin-Aldana, H., Capocelli, K. E., Furuta, G. T. and Ackerman, S. J. | Journal of Allergy and Clinical Immunology | http://dx.doi.org/10.1016/j.jaci.2012.03.005 | Embase |
| Unrelated | Biomarkers Predictive of Exacerbations in the SPIROMICS and COPDGene Cohorts | 2017 | Keene, J. D., Jacobson, S., Kechris, K., Kinney, G. L., Foreman, M. G., Doerschuk, C. M., Make, B. J., Curtis, J. L., Rennard, S. I., Barr, R. G., Bleecker, E. R., Kanner, R. E., Kleerup, E. C., Hansel, N. N., Woodruff, P. G., Han, M. K., Paine, R., 3rd, Martinez, F. J., Bowler, R. P. and O'neal, W. K. | Am J Respir Crit Care Med | 10.1164/rccm.201607-1330OC | PubMed |
| Unrelated | Association between skeletal muscle attenuation and gastroesophageal reflux disease: A health check-up cohort study | 2019 | Kim, Y. M., Kim, J. H., Baik, S. J., Jung, D. H., Park, J. J., Youn, Y. H. and Park, H. | Scientific reports | http://dx.doi.org/10.1038/s41598-019-56702-6 | Embase |
| Unrelated | Phenotypic characteristics and asthma severity in an East African cohort of adults and adolescents with asthma: Findings from the African severe asthma project | 2020 | Kirenga, B., Chakaya, J., Yimer, G., Nyale, G., Haile, T., Muttamba, W., Mugenyi, L., Katagira, W., Worodria, W., Aanyu-Tukamuhebwa, H., Lugogo, N., Joloba, M., Bekele, A., Makumbi, F., Green, C., De Jong, C., Kamya, M. and Van Der Molen, T. | BMJ Open Respiratory Research | http://dx.doi.org/10.1136/bmjresp-2019-000484 | Embase |
| Unrelated | Evaluation of serum neuron specific enolase levels among patients with primary and secondary burning mouth syndrome | 2021 | Kishore, J., Shaikh, F., Zubairi, A. M., Mirza, S., Alqutub, M. N., Almubarak, A. M., Abduljabbar, T. and Vohra, F. | Cephalalgia. | http://dx.doi.org/10.1177/03331024211046613 | Embase |
| Unrelated | Gerd and its' therapeutic management by surgical methods | 2021 | Kumar, N., Mazumder, A. and Das, S. | International Journal of Pharmaceutical Research | http://dx.doi.org/10.31838/ijpr/2021.13.02.348 | Embase |
| Unrelated | Markers of antigen presentation and activation on eosinophils and T cells in the esophageal tissue of patients with eosinophilic esophagitis | 2013 | Le-Carlson, M., Seki, S., Abarbanel, D., Quiros, A., Cox, K. and Nadeau, K. C. | Journal of Pediatric Gastroenterology and Nutrition | http://dx.doi.org/10.1097/MPG.0b013e3182758d49 | Embase |
| Unrelated | H2 receptor antagonists and right ventricular morphology: the MESA right ventricle study | 2014 | Leary, P. J., Barr, R. G., Bluemke, D. A., Bristow, M. R., Kronmal, R. A., Lima, J. A., Ralph, D. D., Ventetuolo, C. E. and Kawut, S. M. | Ann Am Thorac Soc | 10.1513/AnnalsATS.201407-344OC | PubMed |
| Unrelated | Differential protein expression and oncogenic gene network link tyrosine kinase ephrin B4 receptor to aggressive gastric and gastroesophageal junction cancers | 2016 | Liersch-Lohn, B., Slavova, N., Buhr, H. J. and Bennani-Baiti, I. M. | International Journal of Cancer | http://dx.doi.org/10.1002/ijc.29865 | Embase |
| Unrelated | Determinants of exhaled breath condensate pH in a large population with asthma | 2011 | Liu, L., Teague, W. G., Erzurum, S., Fitzpatrick, A., Mantri, S., Dweik, R. A., Bleecker, E. R., Meyers, D., Busse, W. W., Calhoun, W. J., Castro, M., Chung, K. F., Curran-Everett, D., Israel, E., Jarjour, W. N., Moore, W., Peters, S. P., Wenzel, S., Hunt, J. F. and Gaston, B. | Chest | 10.1378/chest.10-0163 | PubMed |
| Unrelated | Hematological indices as indicators of silent inflammation in achalasia patients: A cross-sectional study | 2020 | Lopez-Verdugo, F., Furuzawa-Carballeda, J., Romero-Hernandez, F., Coss-Adame, E., Valdovinos, M. A., Priego-Ranero, A., Olvera-Prado, H., Narvaez-Chavez, S., Peralta-Figueroa, J. and Torres-Villalobos, G. | Medicine (United States) | http://dx.doi.org/10.1097/MD.0000000000019326 | Embase |
| Unrelated | Ectopic fat accumulation in patients with COPD: an ECLIPSE substudy | 2017 | Martin, M., Almeras, N., Després, J. P., Coxson, H. O., Washko, G. R., Vivodtzev, I., Wouters, E. F., Rutten, E., Williams, M. C., Murchison, J. T., Macnee, W., Sin, D. D. and Maltais, F. | Int J Chron Obstruct Pulmon Dis | 10.2147/copd.S124750 | PubMed |
| Unrelated | Impact of self-reported gastroesophageal reflux disease in subjects from COPDGene cohort | 2014 | Martinez, C. H., Okajima, Y., Murray, S., Washko, G. R., Martinez, F. J., Silverman, E. K., Lee, J. H., Regan, E. A., Crapo, J. D., Curtis, J. L., Hatabu, H. and Han, M. K. | Respir Res | 10.1186/1465-9921-15-62 | PubMed |
| Unrelated | Long-term follow-up of malignancy biomarkers in patients with Barrett's esophagus undergoing medical or surgical treatment | 2012 | Martinez De Haro, L. F., Ortiz, A., Parrilla, P., Munitiz, V., Martinez, C. M., Revilla, B., Ruiz De Angulo, D., Bermejo, J., Yélamos, J. and Molina, J. | Ann Surg | 10.1097/SLA.0b013e31824e6c6a | PubMed |
| Unrelated | Serum albumin, total bilirubin, and patient age are independent confounders of hepatobiliary-phase gadoxetate parenchymal liver enhancement | 2019 | Matoori, S., Froehlich, J. M., Breitenstein, S., Pozdniakova, V., Reischauer, C., Kolokythas, O., Koh, D. M. and Gutzeit, A. | Eur Radiol | 10.1007/s00330-019-06179-8 | PubMed |
| Unrelated | COX-2 mRNA is increased in oesophageal mucosal cells by a proton pump inhibitor | 2012 | Mayne, G. C., Watson, D. I. and Hussey, D. J. | ANZ Journal of Surgery | http://dx.doi.org/10.1111/j.1445-2197.2012.06124.x | Embase |
| Unrelated | Cross validated serum small extracellular vesicle microRNAs for the detection of oropharyngeal squamous cell carcinoma | 2020 | Mayne, G. C., Woods, C. M., Dharmawardana, N., Wang, T., Krishnan, S., Hodge, J. C., Foreman, A., Boase, S., Carney, A. S., Sigston, E. a. W., Watson, D. I., Ooi, E. H. and Hussey, D. J. | Journal of Translational Medicine | http://dx.doi.org/10.1186/s12967-020-02446-1 | Embase |
| Unrelated | Association between Gastroesophageal Reflux Disease and Elastographic Parameters of Liver Steatosis and Fibrosis: Controlled Attenuation Parameter and Liver Stiffness Measurements | 2021 | Mikolasevic, I., Poropat, G., Filipec Kanizaj, T., Skenderevic, N., Zelic, M., Matasin, M., Vranic, L., Kresovic, A. and Hauser, G. | Canadian Journal of Gastroenterology and Hepatology | http://dx.doi.org/10.1155/2021/6670065 | Embase |
| Unrelated | Randomized controlled trial comparing aerosolized swallowed fluticasone to esomeprazole for esophageal eosinophilia | 2013 | Moawad, F. J., Veerappan, G. R., Dias, J. A., Baker, T. P., Maydonovitch, C. L. and Wong, R. K. | Am J Gastroenterol | 10.1038/ajg.2012.443 | PubMed |
| Unrelated | Comparison of eotaxin-3 biomarker in patients with eosinophilic oesophagitis, proton pump inhibitor-responsive oesophageal eosinophilia and gastro-oesophageal reflux disease | 2015 | Moawad, F. J., Wells, J. M., Johnson, R. L., Reinhardt, B. J., Maydonovitch, C. L. and Baker, T. P. | Alimentary Pharmacology and Therapeutics | http://dx.doi.org/10.1111/apt.13258 | Embase |
| Unrelated | Gastroesophageal reflux and metabolic syndrome | 2013 | Mocanu, M. A., Diculescu, M. and Dumitrescu, M. | Revista medico-chirurgicala a Societatii de Medici si Naturalisti din Iasi |  | Embase |
| Unrelated | The effect of proton pump inhibitors on the CYP2C19 enzyme activity evaluated by the pantoprazole-(13)C breath test in GERD patients: clinical relevance for personalized medicine | 2016 | Modak, A. S., Klyarytska, I., Kriviy, V., Tsapyak, T. and Rabotyagova, Y. | J Breath Res | 10.1088/1752-7163/10/4/046017 | PubMed |
| Unrelated | A PCR blood test outperforms chromogranin A in carcinoid detection and is unaffected by proton pump inhibitors | 2014 | Modlin, I. M., Aslanian, H., Bodei, L., Drozdov, I. and Kidd, M. | Endocrine Connections | http://dx.doi.org/10.1530/EC-14-0100 | Embase |
| Unrelated | Implications of historical height loss for prevalent vertebral fracture, spinal osteoarthritis, and gastroesophageal reflux disease | 2020 | Nakano, M., Nakamura, Y., Suzuki, T., Kobayashi, T., Takahashi, J. and Shiraki, M. | Scientific reports | https://dx.doi.org/10.1038/s41598-020-76074-6 | Embase |
| Unrelated | Epidemiology | 2016 | Navaratnam, V., Forrester, D. L. and Hubbard, R. B. | ERS Monograph | http://dx.doi.org/10.1183/2312508X.10004615 | Embase |
| Unrelated | Comorbidities are associated with different features of severe asthma 11 Medical and Health Sciences 1102 Cardiorespiratory Medicine and Haematology | 2018 | Novelli, F., Bacci, E., Latorre, M., Seccia, V., Bartoli, M. L., Cianchetti, S., Dente, F. L., Franco, A. D., Celi, A. and Paggiaro, P. | Clinical and Molecular Allergy | http://dx.doi.org/10.1186/s12948-018-0103-x | Embase |
| Unrelated | Treatment of non-erosive reflux disease and dynamics of the esophageal microbiome: a prospective multicenter study | 2020 | Park, C. H., Seo, S. I., Kim, J. S., Kang, S. H., Kim, B. J., Choi, Y. J., Byun, H. J., Yoon, J. H. and Lee, S. K. | Sci Rep | 10.1038/s41598-020-72082-8 | PubMed |
| Unrelated | Nissen fundoplication in Cornelia de Lange syndrome spectrum: Who are the potential candidates? | 2020 | Parma, B., Cianci, P., Mariani, M., Cereda, A., Panceri, R., Fossati, C., Maestri, L., Macchini, F., Onesimo, R., Zampino, G., Betalli, P., Cheli, M. and Selicorni, A. | American Journal of Medical Genetics, Part A | http://dx.doi.org/10.1002/ajmg.a.61625 | Embase |
| Unrelated | Distal mean nocturnal baseline impedance on pH-impedance monitoring predicts reflux burden and symptomatic outcome in gastro-oesophageal reflux disease | 2016 | Patel, A., Wang, D., Sainani, N., Sayuk, G. S. and Gyawali, C. P. | Alimentary Pharmacology and Therapeutics | http://dx.doi.org/10.1111/apt.13777 | Embase |
| Unrelated | Correlation of serum pepsinogens and gastrin-17 with atrophic gastritis in gastroesophageal reflux patients: A matched-pairs study | 2011 | Peitz, U., Wex, T., Vieth, M., Stolte, M., Willich, S., Labenz, J., Jaspersen, D., Lind, T. and Malfertheiner, P. | Journal of Gastroenterology and Hepatology (Australia) | http://dx.doi.org/10.1111/j.1440-1746.2010.06413.x | Embase |
| Unrelated | Objective evidence of gastro-esophageal reflux disease is rare in patients with autoimmune gastritis | 2021 | Pilotto, V., Maddalo, G., Orlando, C., Fassan, M., Rugge, M., Farinati, F. and Savarino, E. | Journal of Gastrointestinal and Liver Diseases | http://dx.doi.org/10.15403/jgld-3033 | Embase |
| Unrelated | Filaggrin and Periostin Expression Is Altered in Eosinophilic Esophagitis and Normalized With Treatment | 2017 | Politi, E., Angelakopoulou, A., Grapsa, D., Zande, M., Stefanaki, K., Panagiotou, I., Roma, E. and Syrigou, E. | J Pediatr Gastroenterol Nutr | 10.1097/mpg.0000000000001419 | PubMed |
| Unrelated | Lymphocytic Esophagitis in Nonachalasia Primary Esophageal Motility Disorders: Improved Criteria, Prevalence, Strength of Association, and Natural History | 2016 | Putra, J., Muller, K. E., Hussain, Z. H., Parker, S., Gabbard, S., Brickley, E. B., Lacy, B. E., Rothstein, R. and Lisovsky, M. | The American journal of surgical pathology |  | Embase |
| Unrelated | Studies of salivary pepsin in patients with gastro-oesophageal reflux disease | 2019 | Race, C., Chowdry, J., Russell, J. M., Corfe, B. M. and Riley, S. A. | Aliment Pharmacol Ther | 10.1111/apt.15138 | PubMed |
| Unrelated | A prospective evaluation of the effect of chronic proton pump inhibitor use on plasma biomarker levels in humans | 2012 | Raines, D., Chester, M., Diebold, A. E., Mamikunian, P., Anthony, C. T., Mamikunian, G. and Woltering, E. A. | Pancreas | 10.1097/MPA.0b013e318243a0b6 | PubMed |
| Unrelated | Sustained benefit from intravenous immunoglobulin therapy for gastrointestinal involvement in systemic sclerosis | 2016 | Raja, J., Nihtyanova, S. I., Murray, C. D., Denton, C. P. and Ong, V. H. | Rheumatology (Oxford) | 10.1093/rheumatology/kev318 | PubMed |
| Unrelated | Routine Preoperative Nutritional Screening in All Primary Total Joint Arthroplasty Patients Has Little Utility | 2020 | Rao, S. S., Chaudhry, Y. P., Solano, M. A., Sterling, R. S., Oni, J. K. and Khanuja, H. S. | Journal of Arthroplasty |  | PubMed |
| Unrelated | Sensiprobe - A miniature thermal device incorporating Peltier technology as a diagnostic tool for studying human oesophageal sensitivity | 2014 | Reeves, J. W., Al-Zinaty, M., Woodland, P., Sifrim, D., Aziz, Q. and Birch, M. J. | Physiological Measurement | http://dx.doi.org/10.1088/0967-3334/35/7/1265 | Embase |
| Unrelated | Exhaled nitric oxide in relation to asthma control: A real-life survey | 2016 | Ricciardolo, F. L. M., Sorbello, V., Bellezza Fontana, R., Schiavetti, I. and Ciprandi, G. | Allergologia et Immunopathologia | http://dx.doi.org/10.1016/j.aller.2015.05.012 | Embase |
| Unrelated | Asthma similarities across ProAR (Brazil) and U-BIOPRED (Europe) adult cohorts of contrasting locations, ethnicity and socioeconomic status | 2020 | Riley, J. H., Bansal, A. T., Davis, M., Bates, S., Adcock, I. M., Chung, K. F., Alcantara-Neves, N., Almeida, P. C. A., Amorim, L., Araujo, M. I., Barnes, K. C., Barreto, M. L., Belitardo, E., Biao-Lima, V., Cardoso, L., Camargos, P. A., Chatkin, J. M., Costa, R. S., Coelho, A. C. C., Cooper, P. J., Cruz, A. A., Cruz, C. S., Cunha, J., De Jesus, J. V., Fernandes, J., Franco, R. A., Gomes-Filho, I., Lima-Matos, A., Figueiredo, C. A., Lessa, M. A., Lins, L., Mello, L. M., Moura-Santos, P., Muniz, I. S., Paixao-Araujo, I., Pinheiro, G. P., Ponte, E. V., Rodrigues, L. C., Santana, C. V. N., Santos-Lima, G., Souza, T. M. O., Souza-Machado, A., Souza-Machado, C., Stelmach, R., Vasquez, V. S., Ahmed, H., Auffray, C., Bakke, P., Baribaud, F., Bel, E. H., Bigler, J., Bisgaard, H., Boedigheimer, M. J., Bonnelykke, K., Brandsma, J., Brinkman, P., Bucchioni, E., Burg, D., Bush, A., Caruso, M., Chaiboonchoe, A., Chanez, P., Compton, C. H., Corfield, J., D'amico, A., Dahlen, B., Dahlen, S. E., De Meulder, B., Djukanovic, R., Erpenbeck, V. J., Erzen, D., Fichtner, K., Fitch, N., Fleming, L. J., Formaggio, E., Fowler, S. J., Frey, U., Gahlemann, M., Geiser, T., Goss, V., Guo, Y. K., Hashimoto, S., Haughney, J., Hedlin, G., Hekking, P. W., Higenbottam, T., Hohlfeld, J. M., Holweg, C., Horvath, I., Howarth, P., James, A. J., Knowles, R. G., Knox, A. J., Krug, N., Lefaudeux, D., Loza, M. J., Lutter, R., Manta, A., Masefield, S., Matthews, J. G., Mazein, A., Meiser, A., Middelveld, R. J. M., Miralpeix, M., Montuschi, P., Mores, N., Murray, C. S., Musial, J., Myles, D., Pahus, L., Pandis, I., Pavlidis, S., Postle, A., Powel, P., Pratico, G., Puig Valls, M., Rao, N., Roberts, A., Roberts, G., Rowe, A., Sandstrom, T., Schofield, J. P. R., Seibold, W., Selby, A., Shaw, D. E., Sigmund, R., Singer, F., Skipp, P. J., Sousa, A. R., Sterk, P. J., Sun, K., Thornton, B., Van Aalderen, W. M., Van Geest, M., Vestbo, J., Vissing, N. H., Wagener, A. H., Wagers, S. S., Weiszhart, Z., Wheelock, C. E. and Wilson, S. J. | Respiratory Medicine | http://dx.doi.org/10.1016/j.rmed.2019.105817 | Embase |
| Unrelated | The Cologne-Bonn cohort: lessons learned | 2015 | Rockstroh, J. K. | Infection. | http://dx.doi.org/10.1007/s15010-015-0745-2 | Embase |
| Unrelated | The frequency of visceral and phenotypic markers in patients with the combination of undifferentiated connective tissue disease and gastroesophageal reflux disease | 2020 | Romash, I. B. and Mishchuk, V. G. | Wiadomosci lekarskie (Warsaw, Poland : 1960) |  | Embase |
| Unrelated | Evaluation of a minimally invasive cell sampling device coupled with assessment of trefoil factor 3 expression for diagnosing Barrett's esophagus: a multi-center case-control study | 2015 | Ross-Innes, C. S., Debiram-Beecham, I., O'donovan, M., Walker, E., Varghese, S., Lao-Sirieix, P., Lovat, L., Griffin, M., Ragunath, K., Haidry, R., Sami, S. S., Kaye, P., Novelli, M., Disep, B., Ostler, R., Aigret, B., North, B. V., Bhandari, P., Haycock, A., Morris, D., Attwood, S., Dhar, A., Rees, C., Rutter, M. D., Sasieni, P. D. and Fitzgerald, R. C. | PLoS Med | 10.1371/journal.pmed.1001780 | PubMed |
| Unrelated | Comparative Cost Effectiveness of Reflux-Based and Reflux-Independent Strategies for Barrett's Esophagus Screening | 2021 | Sami, S. S., Moriarty, J. P., Rosedahl, J. K., Borah, B. J., Katzka, D. A., Wang, K. K., Kisiel, J. B., Ragunath, K., Rubenstein, J. H. and Iyer, P. G. | The American journal of gastroenterology | https://dx.doi.org/10.14309/ajg.0000000000001336 | Embase |
| Unrelated | Association of eosinophil-mediated inflammatory biomarkers with the presence of the Schatzki ring | 2021 | Sarbinowska, J., Wiatrak, B. and Wasko-Czopnik, D. | Advances in Medical Sciences | http://dx.doi.org/10.1016/j.advms.2021.05.004 | Embase |
| Unrelated | Predisposing factors for positive D-Xylose breath test for evaluation of small intestinal bacterial overgrowth: a retrospective study of 932 patients | 2015 | Schatz, R. A., Zhang, Q., Lodhia, N., Shuster, J., Toskes, P. P. and Moshiree, B. | World J Gastroenterol | 10.3748/wjg.v21.i15.4574 | PubMed |
| Unrelated | A prospective randomized study of systemic inflammation and immune response after laparoscopic nissen fundoplication performed with standard and low-pressure pneumoperitoneum | 2013 | Schietroma, M., Carlei, F., Cecilia, E. M., Piccione, F., Sista, F., De Vita, F. and Amicucci, G. | Surg Laparosc Endosc Percutan Tech | 10.1097/SLE.0b013e3182827e51 | PubMed |
| Unrelated | Vorinostat in refractory soft tissue sarcomas - Results of a multi-centre phase II trial of the German Soft Tissue Sarcoma and Bone Tumour Working Group (AIO) | 2016 | Schmitt, T., Mayer-Steinacker, R., Mayer, F., Grünwald, V., Schütte, J., Hartmann, J. T., Kasper, B., Hüsing, J., Hajda, J., Ottawa, G., Mechtersheimer, G., Mikus, G., Burhenne, J., Lehmann, L., Heilig, C. E., Ho, A. D. and Egerer, G. | Eur J Cancer | 10.1016/j.ejca.2016.05.018 | PubMed |
| Unrelated | Capsaicin and evodiamine ingestion does not augment energy expenditure and fat oxidation at rest or after moderately-intense exercise | 2013 | Schwarz, N. A., Spillane, M., La Bounty, P., Grandjean, P. W., Leutholtz, B. and Willoughby, D. S. | Nutr Res | 10.1016/j.nutres.2013.08.007 | PubMed |
| Unrelated | Sex differences in severe asthma: Results from severe asthma network in Italy-SANI | 2021 | Senna, G., Latorre, M., Bugiani, M., Caminati, M., Heffler, E., Morrone, D., Paoletti, G., Parronchi, P., Puggioni, F., Blasi, F., Canonica, G. W. and Paggiaro, P. | Allergy, Asthma and Immunology Research | https://dx.doi.org/10.4168/AAIR.2021.13.2.219 | Embase |
| Unrelated | Complex Gastrointestinal and Endocrine Sources of Inflammation in Schizophrenia | 2020 | Severance, E. G., Dickerson, F. and Yolken, R. H. | Frontiers in Psychiatry | http://dx.doi.org/10.3389/fpsyt.2020.00549 | Embase |
| Unrelated | Gastrointestinal symptoms in idiopathic pulmonary fibrosis patients treated with pirfenidone and herbal medicine | 2014 | Shimizu, Y., Shimoyama, Y., Kawada, A., Kusano, M., Hosomi, Y., Sekiguchi, M., Kawata, T., Horie, T., Ishii, Y., Yamada, M., Dobashi, K. and Takise, A. | J Biol Regul Homeost Agents |  | PubMed |
| Unrelated | Systemic Sclerosis in Zimbabwe: Autoantibody Biomarkers, Clinical, and Laboratory Correlates | 2021 | Sibanda, E. N., Dube, Y., Chakawa, M., Mduluza, T. and Mutapi, F. | Frontiers in Immunology | https://dx.doi.org/10.3389/fimmu.2021.679531 | Embase |
| Unrelated | Gastric adenocarcinoma with chief cell differentiation: a proposal for reclassification as oxyntic gland polyp/adenoma | 2012 | Singhi, A. D., Lazenby, A. J. and Montgomery, E. A. | Am J Surg Pathol | 10.1097/PAS.0b013e31825033e7 | PubMed |
| Unrelated | Prospective single arm study on the effect of ilaprazole in patients with heartburn but no reflux esophagitis | 2018 | Song, I. J., Kim, H. K., Lee, N. K. and Lee, S. K. | Yonsei Medical Journal | http://dx.doi.org/10.3349/ymj.2018.59.8.951 | Embase |
| Unrelated | Is electrical brain activity a reliable biomarker for opioid analgesia in the gut? | 2011 | Staahl, C., Krarup, A. L., Olesen, A. E., Brock, C., Graversen, C. and Drewes, A. M. | Basic Clin Pharmacol Toxicol | 10.1111/j.1742-7843.2011.00727.x | PubMed |
| Unrelated | Prevalence, characteristics and outcome of non-cardiac chest pain and elevated copeptin levels | 2014 | Stallone, F., Twerenbold, R., Wildi, K., Reichlin, T., Rubini Gimenez, M., Haaf, P., Fuechslin, N., Hillinger, P., Jaeger, C., Kreutzinger, P., Puelacher, C., Radosavac, M., Moreno Weidmann, Z., Moehring, B., Honegger, U., Schumacher, C., Denhaerynck, K., Arnold, C., Bingisser, R., Vollert, J. O., Osswald, S. and Mueller, C. | Heart | 10.1136/heartjnl-2014-305583 | PubMed |
| Unrelated | A prospective cohort study on overweight, smoking, alcohol consumption, and risk of Barrett's esophagus | 2011 | Steevens, J., Schouten, L. J., Driessen, A. L., Huysentruyt, C. J., Keulemans, Y. C., Goldbohm, R. A. and Van Den Brandt, P. A. | Cancer Epidemiol Biomarkers Prev | 10.1158/1055-9965.Epi-10-0636 | PubMed |
| Unrelated | Clinical and autoantibody profile in systemic sclerosis: Baseline characteristics from a West Malaysian cohort | 2015 | Sujau, I., Ng, C. T., Sthaneshwar, P., Sockalingam, S., Cheah, T. E., Yahya, F. and Jasmin, R. | International Journal of Rheumatic Diseases | http://dx.doi.org/10.1111/1756-185X.12322 | Embase |
| Unrelated | Improved Cough and Cough-Specific Quality of Life in Patients Treated for Scleroderma-Related Interstitial Lung Disease: Results of Scleroderma Lung Study II | 2017 | Tashkin, D. P., Volkmann, E. R., Tseng, C. H., Roth, M. D., Khanna, D., Furst, D. E., Clements, P. J., Theodore, A., Kafaja, S., Kim, G. H., Goldin, J., Ariolla, E. and Elashoff, R. M. | Chest | http://dx.doi.org/10.1016/j.chest.2016.11.052 | Embase |
| Unrelated | Blood mRNA levels of T cells and IgE receptors are novel non-invasive biomarkers for eosinophilic esophagitis (EoE) | 2021 | Upparahalli Venkateshaiah, S., Rayapudi, M., Kandikattu, H. K., Yadavalli, C. S. and Mishra, A. | Clinical Immunology | http://dx.doi.org/10.1016/j.clim.2021.108752 | Embase |
| Unrelated | Changes in obesity-related diseases and biochemical variables after laparoscopic sleeve gastrectomy: a two-year follow-up study | 2014 | Vage, V., Sande, V. A., Mellgren, G., Laukeland, C., Behme, J. and Andersen, J. R. | BMC surgery | http://dx.doi.org/10.1186/1471-2482-14-8 | Embase |
| Unrelated | Iron deficiency in worsening heart failure is associated with reduced estimated protein intake, fluid retention, inflammation, and antiplatelet use | 2019 | Van Der Wal, H. H., Grote Beverborg, N., Dickstein, K., Anker, S. D., Lang, C. C., Ng, L. L., Van Veldhuisen, D. J., Voors, A. A. and Van Der Meer, P. | Eur Heart J | 10.1093/eurheartj/ehz680 | PubMed |
| Unrelated | Clinical symptom tool that raises the index of suspicion for eosinophilic oesophagitis in adults and drives earlier biopsy for definitive diagnosis | 2017 | Von Arnim, U., Rohl, F. W., Miehlke, S., Jechorek, D., Reinhold, D., Wex, T. and Malfertheiner, P. | Alimentary Pharmacology and Therapeutics | http://dx.doi.org/10.1111/apt.13869 | Embase |
| Unrelated | Identification of clinical and laboratory markers for predicting eosinophilic esophagitis in adults | 2011 | Von Arnim, U., Wex, T., Rohl, F. W., Neumann, H., Kuster, D., Weigt, J., Monkemuller, K. and Malfertheiner, P. | Digestion | http://dx.doi.org/10.1159/000331142 | Embase |
| Unrelated | Bronchoalveolar pH and inflammatory biomarkers in newly diagnosed IPF and GERD patients: A case-control study | 2014 | Vukovac, E. L., Lozo, M., Mise, K., Gudelj, I., Puljiz, Z., Jurcev-Savicevic, A., Bradaric, A., Kokeza, J. and Mise, J. | Medical Science Monitor | http://dx.doi.org/10.12659/MSM.889800 | Embase |
| Unrelated | Dose-response effect of Bifidobacterium lactis HN019 on whole gut transit time and functional gastrointestinal symptoms in adults | 2011 | Waller, P. A., Gopal, P. K., Leyer, G. J., Ouwehand, A. C., Reifer, C., Stewart, M. E. and Miller, L. E. | Scand J Gastroenterol | 10.3109/00365521.2011.584895 | PubMed |
| Unrelated | Two Years Remission of Type 2 Diabetes Mellitus after Bariatric Surgery | 2019 | Wazir, N., Arshad, M. F., Finney, J., Kirk, K. and Dewan, S. | J Coll Physicians Surg Pak | 10.29271/jcpsp.2019.10.967 | PubMed |
| Unrelated | Leukocyte telomere length in relation to the risk of Barrett's esophagus and esophageal adenocarcinoma | 2016 | Wennerstrom, E. C. M., Risques, R. A., Prunkard, D., Giffen, C., Corley, D. A., Murray, L. J., Whiteman, D. C., Wu, A. H., Bernstein, L., Ye, W., Chow, W., Vaughan, T. L. and Liao, L. M. | Cancer Medicine | http://dx.doi.org/10.1002/cam4.810 | Embase |
| Unrelated | Reflux-Associated Oxygen Desaturations: Usefulness in Diagnosing Reflux-Related Respiratory Symptoms | 2013 | Wilshire, C. L., Salvador, R., Sepesi, B., Niebisch, S., Watson, T. J., Litle, V. R., Peyre, C. G., Jones, C. E. and Peters, J. H. | Journal of Gastrointestinal Surgery | http://dx.doi.org/10.1007/s11605-012-2065-5 | Embase |
| Unrelated | Lymphocytic Esophagitis With CD4 T-cell-predominant Intraepithelial Lymphocytes and Primary Esophageal Motility Abnormalities: A Potential Novel Clinicopathologic Entity | 2015 | Xue, Y., Suriawinata, A., Liu, X., Li, Z., Gabbard, S., Rothstein, R., Lacy, B. and Lisovsky, M. | Am J Surg Pathol | 10.1097/pas.0000000000000493 | PubMed |
| Unrelated | A urine and serum metabolomics study of gastroesophageal reflux disease in TCM syndrome differentiation using UPLC-Q-TOF/MS | 2021 | Ye, X., Wang, X., Wang, Y., Sun, W., Chen, Y., Wang, D. and Li, Z. | Journal of Pharmaceutical and Biomedical Analysis | http://dx.doi.org/10.1016/j.jpba.2021.114369 | Embase |
| Unrelated | Shar Pei Larynx: Supraglottic and Postcricoid Mucosal Redundancy and Its Association With Medical Comorbidities | 2019 | Yiu, Y., Tibbetts, K. M., Simpson, C. B. and Matrka, L. A. | Ann Otol Rhinol Laryngol | 10.1177/0003489418810893 | PubMed |
| Unrelated | The Association of Depressive Symptoms With Rates of Acute Exacerbations in Patients With COPD: Results From a 3-year Longitudinal Follow-up of the ECLIPSE Cohort | 2017 | Yohannes, A. M., Mülerová, H., Lavoie, K., Vestbo, J., Rennard, S. I., Wouters, E. and Hanania, N. A. | J Am Med Dir Assoc | 10.1016/j.jamda.2017.05.024 | PubMed |
| Unrelated | Bronchoalveolar bile acid and inflammatory markers to identify high-risk lung transplant recipients with reflux and microaspiration | 2020 | Zhang, C. Y. K., Ahmed, M., Huszti, E., Levy, L., Hunter, S. E., Boonstra, K. M., Moshkelgosha, S., Sage, A. T., Azad, S., Zamel, R., Ghany, R., Yeung, J. C., Crespin, O. M., Frankel, C., Budev, M., Shah, P., Reynolds, J. M., Snyder, L. D., Belperio, J. A., Singer, L. G., Weigt, S. S., Todd, J. L., Palmer, S. M., Keshavjee, S. and Martinu, T. | Journal of Heart and Lung Transplantation | http://dx.doi.org/10.1016/j.healun.2020.05.006 | Embase |
| Unrelated | JianpiQinghua granule reduced PPI dosage in patients with nonerosive reflux disease: A multicenter, randomized, double-blind, double-dummy, noninferiority study | 2021 | Zhang, J., Che, H., Zhang, B., Zhang, C., Zhou, B., Ji, H., Xie, J., Shi, X., Li, X., Wang, F. and Tang, X. | Phytomedicine | http://dx.doi.org/10.1016/j.phymed.2021.153584 | Embase |
| Unrelated | Targeting the COX1/2-Driven thromboxane A2 pathway suppresses Barrett's esophagus and esophageal adenocarcinoma development | 2019 | Zhang, T., Wang, Q., Ma, W. Y., Wang, K., Chang, X., Johnson, M. L., Bai, R., Bode, A. M., Foster, N. R., Falk, G. W., Limburg, P. J., Iyer, P. G. and Dong, Z. | EBioMedicine | http://dx.doi.org/10.1016/j.ebiom.2019.10.038 | Embase |
| Unrelated | Oesophageal intrasquamous IgG4 deposits: an adjunctive marker to distinguish eosinophilic oesophagitis from reflux oesophagitis | 2016 | Zukerberg, L., Mahadevan, K., Selig, M. and Deshpande, V. | Histopathology | 10.1111/his.12892 | PubMed |
| Pediatric Study | Eosinophilic oesophagitis: A paediatric update | 2015 | Allen, K. J. and Heine, R. G. | Journal of Paediatrics and Child Health | http://dx.doi.org/10.1111/jpc.12757 | Embase |
| Pediatric Study | Does positioning affect tracheal aspiration of gastric content in ventilated infants? | 2015 | Aly, H., Soliman, R. M., El-Dib, M., Said, R. N., Abdellatif, M. A., Sibaii, H. and Elwakkad, A. | J Pediatr Gastroenterol Nutr | 10.1097/mpg.0000000000000601 | PubMed |
| Pediatric Study | Pepsinogen identification in the middle ear fluid of children with otitis media with effusion | 2016 | Buyruk, A., Osma, U., Yilmaz, M. D. and Eyigor, H. | Kulak burun bogaz ihtisas dergisi : KBB = Journal of ear, nose, and throat | http://dx.doi.org/10.5606/kbbihtisas.2016.83669 | Embase |
| Pediatric Study | Short-Term Neurodevelopmental Outcome in Children Born With High-Risk Congenital Lung Lesions | 2018 | Danzer, E., Hoffman, C., D'agostino, J. A., Boelig, M. M., Gerdes, M., Bernbaum, J. C., Rosenthal, H., Waqar, L. N., Rintoul, N. E., Herkert, L. M., Kallan, M. J., Peranteau, W. H., Flake, A. W., Adzick, N. S. and Hedrick, H. L. | Annals of Thoracic Surgery | http://dx.doi.org/10.1016/j.athoracsur.2018.01.033 | Embase |
| Pediatric Study | Autism spectrum disorder and neurodevelopmental delays in children with giant omphalocele | 2019 | Danzer, E., Hoffman, C., Miller, J. S., D'agostino, J. A., Schindewolf, E. M., Gerdes, M., Bernbaum, J., Adams, S. E., Rintoul, N. E., Herkert, L. M., Taylor, L., Schreiber, J., Peranteau, W. H., Flake, A. W., Adzick, N. S. and Hedrick, H. L. | Journal of Pediatric Surgery | http://dx.doi.org/10.1016/j.jpedsurg.2019.05.017 | Embase |
| Pediatric Study | Immunoreactivity of p53 and Ki-67 for dysplastic changes in children with eosinophilic esophagitis | 2013 | Denning, K. L., Al-Subu, A. and Elitsur, Y. | Pediatric and Developmental Pathology | http://dx.doi.org/10.2350/13-03-1306-OA.1 | Embase |
| Pediatric Study | May failure to thrive in infants be a clinical marker for the early diagnosis of cow's milk allergy? | 2020 | Diaferio, L., Caimmi, D., Verga, M. C., Palladino, V., Trove, L., Giordano, P., Verduci, E. and Miniello, V. L. | Nutrients | http://dx.doi.org/10.3390/nu12020466 | Embase |
| Pediatric Study | Esomeprazole FDA Approval in Children with GERD: Exposure-Matching and Exposure-Response | 2017 | Earp, J. C., Mehrotra, N., Peters, K. E., Fiorentino, R. P., Griebel, D., Lee, S. C., Mulberg, A., Rohss, K., Sandstrom, M., Taylor, A., Tornoe, C. W., Wynn, E. L., Van Der Walt, J. S. and Garnett, C. | Journal of Pediatric Gastroenterology and Nutrition | http://dx.doi.org/10.1097/MPG.0000000000001467 | Embase |
| Pediatric Study | Exhaled breath condensate pH does not discriminate asymptomatic gastroesophageal reflux or the response to lansoprazole treatment in children with poorly controlled asthma | 2014 | Fitzpatrick, A. M., Holbrook, J. T., Wei, C. Y., Brown, M. S., Wise, R. A. and Teague, W. G. | J Allergy Clin Immunol Pract | 10.1016/j.jaip.2014.04.006 | PubMed |
| Pediatric Study | Pepsin in saliva as a biomarker for oropharyngeal reflux compared with 24-hour esophageal impedance/pH monitoring in pediatric patients | 2017 | Fortunato, J. E., D'agostino, R. B. and Lively, M. O. | Neurogastroenterology and Motility | http://dx.doi.org/10.1111/nmo.12936 | Embase |
| Pediatric Study | Is histologic esophagitis associated with dental erosion: a cross-sectional observational study? | 2017 | Friesen, L. R., Bohaty, B., Onikul, R., Walker, M. P., Abraham, C., Williams, K. B., Cocjin, J. T., Cocjin, E. L. and Friesen, C. A. | BMC Oral Health | 10.1186/s12903-017-0408-z | PubMed |
| Pediatric Study | Association between tracheal pepsin, a reliable marker of gastric aspiration, and head of bed elevation among ventilated neonates | 2014 | Garland, J. S., Alex, C. P., Johnston, N., Yan, J. C. and Werlin, S. L. | J Neonatal Perinatal Med | 10.3233/npm-14814020 | PubMed |
| Pediatric Study | Salivary pepsin A detection related to gastro-oesophageal reflux episodes in children undergoing impedance probe monitoring | 2020 | Haddad, H. A., He, Z., Shaffer, S. E. and Molle-Rios, Z. L. | Acta Paediatrica, International Journal of Paediatrics | http://dx.doi.org/10.1111/apa.15276 | Embase |
| Pediatric Study | Effect of right lateral position with head elevation on tracheal aspirate pepsin in ventilated preterm neonates: randomized controlled trial | 2019 | Imam, S. S., Shinkar, D. M., Mohamed, N. A. and Mansour, H. E. | J Matern Fetal Neonatal Med | 10.1080/14767058.2018.1471674 | PubMed |
| Pediatric Study | Measurement of mucosal conductivity by MII is a potential marker of mucosal integrity restored in infants on acid-suppression therapy | 2011 | Loots, C. M., Van Wijk, M. P., Smits, M. J., Wenzl, T. G., Benninga, M. A. and Omari, T. I. | J Pediatr Gastroenterol Nutr | 10.1097/MPG.0b013e318214c3cc | PubMed |
| Pediatric Study | Metabolomic profiling of extraesophageal reflux disease in children | 2021 | Mahoney, L. B., Esther, C. R., May, K. and Rosen, R. | Clinical and Translational Science | http://dx.doi.org/10.1111/cts.13064 | Embase |
| Pediatric Study | Diagnostic value of a pattern of exhaled breath condensate biomarkers in asthmatic children | 2017 | Maloca Vuljanko, I., Turkalj, M., Nogalo, B., Bulat Lokas, S. and Plavec, D. | Allergologia et Immunopathologia |  | PubMed |
| Pediatric Study | Molecular abnormalities in pediatric barrett esophagus: Can we test for potential of neoplastic progression? | 2010 | Maltby, E. L., Dyson, M. J., Wheeler, M. R., Thomson, M., Sethuraman, C. and Cohen, M. C. | Pediatric and Developmental Pathology | http://dx.doi.org/10.2350/09-08-0700-OA.1 | Embase |
| Pediatric Study | Pediatric Eosinophilic Esophagitis Symptom Scores (PEESS v2.0) identify histologic and molecular correlates of the key clinical features of disease | 2015 | Martin, L. J., Franciosi, J. P., Collins, M. H., Abonia, J. P., Lee, J. J., Hommel, K. A., Varni, J. W., Grotjan, J. T., Eby, M., He, H., Marsolo, K., Putnam, P. E., Garza, J. M., Kaul, A., Wen, T. and Rothenberg, M. E. | Journal of Allergy and Clinical Immunology | http://dx.doi.org/10.1016/j.jaci.2015.03.004 | PubMed |
| Pediatric Study | Correlation of ALOX15 expression with eosinophilic or reflux esophagitis in a cohort of pediatric patients with esophageal eosinophilia | 2014 | Matoso, A., Allen, D., Herzlinger, M., Ferreira, J., Chen, S., Lu, S., Fabre, V., Monahan, R., Yang, D., Noble, L., Mangray, S. and Resnick, M. B. | Human Pathology | http://dx.doi.org/10.1016/j.humpath.2014.01.021 | PubMed |
| Pediatric Study | Clinical outcome of a randomized controlled blinded trial of open versus laparoscopic Nissen fundoplication in infants and children | 2011 | Mchoney, M., Wade, A. M., Eaton, S., Howard, R. F., Kiely, E. M., Drake, D. P., Curry, J. I. and Pierro, A. | Ann Surg | 10.1097/SLA.0b013e318226727f | PubMed |
| Pediatric Study | Nutritional State and Feeding Behaviors of Children with Eosinophilic Esophagitis and Gastroesophageal Reflux Disease | 2018 | Mehta, P., Furuta, G. T., Brennan, T., Henry, M. L., Maune, N. C., Sundaram, S. S., Menard-Katcher, C., Atkins, D., Takurukura, F., Giffen, S., Pan, Z. and Haas, A. M. | Journal of Pediatric Gastroenterology and Nutrition | http://dx.doi.org/10.1097/MPG.0000000000001741 | Embase |
| Pediatric Study | Response to therapy among neonates with gastro-esophageal reflux is associated with esophageal clearance | 2021 | Nobile, S., Meneghin, F., Marchionni, P., Noviello, C., Salvatore, S., Lista, G., Carnielli, V. P. and Vento, G. | Early Hum Dev | 10.1016/j.earlhumdev.2020.105248 | PubMed |
| Pediatric Study | Pediatric Eosinophilic Esophagitis | 2018 | Posten, S., Adamiak, T. and Jensen, M. | South Dakota medicine: the journal of the South Dakota State Medical Association |  | Embase |
| Pediatric Study | The presence of pepsin in the lung and its relationship to pathologic gastro-esophageal reflux | 2012 | Rosen, R., Johnston, N., Hart, K., Khatwa, U. and Nurko, S. | Neurogastroenterology and Motility | http://dx.doi.org/10.1111/j.1365-2982.2011.01826.x | Embase |
| Pediatric Study | Eosinophilic Esophagitis: Update on Diagnosis and Treatment in Pediatric Patients | 2020 | Rossetti, D., Isoldi, S. and Oliva, S. | Pediatric Drugs | http://dx.doi.org/10.1007/s40272-020-00398-z | Embase |
| Pediatric Study | Mean platelet volume and red cell distribution width as potential new biomarkers in children with gastroesophageal reflux disease | 2019 | Sevencan, N. O., Cesur, O., Cakar, M., Dogan, E., Ozkan, A. E. and Benli, A. R. | American Journal of Translational Research |  | Embase |
| Pediatric Study | Blind esophageal brushing offers a safe and accurate method to monitor inflammation in children and young adults with eosinophilic esophagitis | 2018 | Smadi, Y., Deb, C., Bornstein, J., Safder, S., Horvath, K. and Mehta, D. | Diseases of the Esophagus | http://dx.doi.org/10.1093/dote/doy056 | Embase |
| Pediatric Study | Prenatal Ultrasound Measurement of Fetal Stomach Size Is Predictive of Postnatal Development of GERD in Isolated Cleft Lip and/or Palate | 2021 | Toscano, M., Burhans, K., Mack, L. M., Henderson, S., Koltz, P. F., Girotto, J. A. and Thornburg, L. L. | Cleft Palate-Craniofacial Journal | http://dx.doi.org/10.1177/1055665620968717 | Embase |
| Invasive/Histochemical Analysis | Effect of Acid Suppression on Peripheral T-Lymphocyte Subsets and Immunohistochemical Esophageal Mucosal Changes in Patients With Gastroesophageal Reflux Disease | 2019 | Ahmed Osman, H., Aly, S. S., Mahmoud, H. S., Ahmed, E. H., Salah Eldin, E. M., Abdelrahim, E. A., El Masry, M. A., Herdan, R. A. and Hassan, M. H. | J Clin Gastroenterol | 10.1097/mcg.0000000000001098 | PubMed |
| Invasive/Histochemical Analysis | Immunohistochemical analysis of Ki-67, p53 and Bcl-2 expression related to histological features in gastroesophageal reflux disease | 2010 | Ayhan, S., Ak Nalbant, O., Isisag, A., Turkel Kucukmetin, N. and Temiz, P. | Turkish Journal of Gastroenterology | http://dx.doi.org/10.4318/tjg.2010.0088 | Embase |
| Invasive/Histochemical Analysis | Highlighted role of "IL17 signaling pathway" in gastroesophageal reflux disease | 2020 | Azodi, M. Z., Razzaghi, M., Malekpour, H., Heidari, M. H. and Rezaei-Tavirani, M. | Gastroenterology and Hepatology from Bed to Bench |  | Embase |
| Invasive/Histochemical Analysis | Prolonged exposure to acid and bile induces chromosome abnormalities that precede malignant transformation of benign Barrett's epithelium | 2012 | Bajpai, M., Aviv, H. and Das, K. M. | Molecular Cytogenetics | http://dx.doi.org/10.1186/1755-8166-5-43 | Embase |
| Invasive/Histochemical Analysis | Recurring Translocations in Barrett's Esophageal Adenocarcinoma | 2021 | Bajpai, M., Panda, A., Birudaraju, K., Van Gurp, J., Chak, A., Das, K. M., Javidian, P. and Aviv, H. | Frontiers in Genetics | http://dx.doi.org/10.3389/fgene.2021.674741 | Embase |
| Invasive/Histochemical Analysis | Immunohistochemistry for CDX2 expression in non-goblet-cell Barrett's oesophagus | 2014 | Behan, M., Gledhill, A. and Hayes, S. | British journal of biomedical science |  | Embase |
| Invasive/Histochemical Analysis | Foveolar type dysplasia in Barrett esophagus | 2010 | Brown, I. S., Whiteman, D. C. and Lauwers, G. Y. | Modern Pathology | http://dx.doi.org/10.1038/modpathol.2010.59 | Embase |
| Invasive/Histochemical Analysis | Distinct proteomic profiles characterise non-erosive from erosive reflux disease | 2011 | Calabrese, C., Marzano, V., Urbani, A., Lazzarini, G., Valerii, M. C., Liguori, G., Di Molfetta, S., Rizzello, F., Gionchetti, P., Campieri, M. and Spisni, E. | Alimentary Pharmacology and Therapeutics | http://dx.doi.org/10.1111/j.1365-2036.2011.04801.x | Embase |
| Invasive/Histochemical Analysis | Activation of the BMP4 Pathway and Early Expression of CDX2 Characterize Non-specialized Columnar Metaplasia in a Human Model of Barrett's Esophagus | 2012 | Castillo, D., Puig, S., Iglesias, M., Seoane, A., De Bolos, C., Munitiz, V., Parrilla, P., Comerma, L., Poulsom, R., Krishnadath, K. K., Grande, L. and Pera, M. | Journal of Gastrointestinal Surgery | http://dx.doi.org/10.1007/s11605-011-1758-5 | Embase |
| Invasive/Histochemical Analysis | Evaluation of Barrett's esophagus with CK7, CK20, p53, Ki67, and COX2 expressions using chromoendoscopical examination | 2013 | Çoban, Ş., Örmeci, N., Savaş, B., Ekiz, F., Ensari, A., Kuzu, I. and Palabıyıkoğlu, M. | Dis Esophagus | 10.1111/j.1442-2050.2012.01352.x | PubMed |
| Invasive/Histochemical Analysis | Differential MicroRNA signatures in the pathogenesis of barrett's esophagus | 2020 | Craig, M. P., Rajakaruna, S., Paliy, O., Sajjad, M., Madhavan, S., Reddy, N., Zhang, J., Bottomley, M., Agrawal, S. and Kadakia, M. P. | Clinical and Translational Gastroenterology | http://dx.doi.org/10.14309/ctg.0000000000000125 | Embase |
| Invasive/Histochemical Analysis | Proteomic Analysis in Esophageal Eosinophilia Reveals Differential Galectin-3 Expression and S-Nitrosylation | 2016 | Davis, C. M., Hiremath, G., Wiktorowicz, J. E., Soman, K. V., Straub, C., Nance, C., Quintanilla, N., Pazdrak, K., Thakkar, K., Olive, A. P. and Kurosky, A. | Digestion | http://dx.doi.org/10.1159/000444675 | Embase |
| Invasive/Histochemical Analysis | Characterization of oncocytes in deep esophageal glands | 2016 | Gonzalez, G., Huang, Q. and Mashimo, H. | Diseases of the Esophagus | http://dx.doi.org/10.1111/dote.12382 | Embase |
| Invasive/Histochemical Analysis | A Novel Susceptibility Locus Near GRIK2 Associated With Erosive Esophagitis in a Korean Cohort | 2020 | Jin, E. H., Park, B., Kim, Y. S., Choe, E. K., Choi, S. H., Kim, J. S. and Jung, S. A. | Clin Transl Gastroenterol | 10.14309/ctg.0000000000000145 | PubMed |
| Invasive/Histochemical Analysis | Role of e-cadherin in the pathogenesis of gastroesophageal reflux disease | 2011 | Jovov, B., Que, J., Tobey, N. A., Djukic, Z., Hogan, B. L. M. and Orlando, R. C. | American Journal of Gastroenterology |  | PubMed |
| Invasive/Histochemical Analysis | Disintegrin and metalloproteinases (ADAMs) expression in gastroesophageal reflux disease and in esophageal adenocarcinoma | 2017 | Kauttu, T., Mustonen, H., Vainionpää, S., Krogerus, L., Ilonen, I., Räsänen, J., Salo, J. and Puolakkainen, P. | Clin Transl Oncol | 10.1007/s12094-016-1503-3 | PubMed |
| Invasive/Histochemical Analysis | Multilayered epithelium at the gastroesophageal junction is a marker of gastroesophageal reflux disease: data from a prospective Central European multicenter study (histoGERD trial) | 2014 | Langner, C., Wolf, E. M., Plieschnegger, W., Geppert, M., Wigginghaus, B., Höss, G. M., Eherer, A., Schneider, N. I., Rehak, P. and Vieth, M. | Virchows Arch | 10.1007/s00428-014-1550-5 | PubMed |
| Invasive/Histochemical Analysis | Squamous tissue lymphocytes in the esophagus of controls and patients with reflux esophagitis and Barrett's esophagus are characterized by a non-inflammatory phenotype | 2014 | Lind, A., Koenderman, L., Kusters, J. G. and Siersema, P. D. | PLoS One | 10.1371/journal.pone.0106261 | PubMed |
| Invasive/Histochemical Analysis | MUC2 is a highly specific marker of goblet cell metaplasia in the distal esophagus and gastroesophageal junction | 2011 | Mcintire, M. G., Soucy, G., Vaughan, T. L., Shahsafaei, A. and Odze, R. D. | American Journal of Surgical Pathology | http://dx.doi.org/10.1097/PAS.0b013e318218940d | Embase |
| Invasive/Histochemical Analysis | Histology of symptomatic gastroesophageal reflux disease: is it predictive of response to proton pump inhibitors? | 2013 | Miwa, H., Takubo, K., Shimatani, T., Furuta, T., Oshima, T., Tanaka, J., Aida, J., Ito, M., Kurosawa, S., Joh, T., Wada, T., Habu, Y., Watanabe, Y., Hongo, M., Chiba, T. and Kinoshita, Y. | J Gastroenterol Hepatol | 10.1111/j.1440-1746.2012.07266.x | PubMed |
| Invasive/Histochemical Analysis | The histological and immunohistochemical aspects of bile reflux in patients with gastroesophageal reflux disease | 2011 | Nakos, A., Kouklakis, G., Pitiakoudis, M., Zezos, P., Efraimidou, E., Giatromanolaki, A., Polychronidis, A., Liratzopoulos, N., Sivridis, E. and Simopoulos, K. | Gastroenterology Research and Practice | http://dx.doi.org/10.1155/2011/905872 | Embase |
| Invasive/Histochemical Analysis | Carditis: a relevant marker of gastroesophageal reflux disease. Data from a prospective central European multicenter study on histological and endoscopic diagnosis of esophagitis (histoGERD trial) | 2019 | Pinto, D., Plieschnegger, W., Schneider, N. I., Geppert, M., Bordel, H., Höss, G. M., Eherer, A., Wolf, E. M., Vieth, M. and Langner, C. | Dis Esophagus | 10.1093/dote/doy073 | PubMed |
| Invasive/Histochemical Analysis | Immunohistochemical assessment of Survivin and Bcl3 expression as potential biomarkers for NF-kappaB activation in the Barrett metaplasia-dysplasia-adenocarcinoma sequence | 2018 | Puccio, I., Khan, S., Butt, A., Graham, D., Sehgal, V., Patel, D., Novelli, M., Lovat, L. B., Rodriguez-Justo, M. and Hamoudi, R. A. | International Journal of Experimental Pathology | http://dx.doi.org/10.1111/iep.12260 | Embase |
| Invasive/Histochemical Analysis | Use of direct, endoscopic-guided measurements of mucosal impedance in diagnosis of gastroesophageal reflux disease | 2012 | Saritas Yuksel, E., Higginbotham, T., Slaughter, J. C., Mabary, J., Kavitt, R. T., Garrett, C. G. and Vaezi, M. F. | Clin Gastroenterol Hepatol | 10.1016/j.cgh.2012.05.018 | PubMed |
| Invasive/Histochemical Analysis | Polymorphisms of Genes Related to Function and Metabolism of Vitamin D in Esophageal Adenocarcinoma | 2019 | Singhal, S., Kapoor, H., Subramanian, S., Agrawal, D. K. and Mittal, S. K. | J Gastrointest Cancer | 10.1007/s12029-018-0164-6 | PubMed |
| Invasive/Histochemical Analysis | Identification of unique transcriptomic signatures and hub genes through rna sequencing and integrated wgcna and ppi network analysis in nonerosive reflux disease | 2021 | Zhao, Y., Ma, T. and Zou, D. | Journal of Inflammation Research | http://dx.doi.org/10.2147/JIR.S340452 | Embase |
| Systematic Review/ Meta-Analysis | Current hurdles in the management of eosinophilic oesophagitis: The next steps | 2013 | Attwood, S. E. A. and Wilson, M. S. J. | World Journal of Gastroenterology | https://dx.doi.org/10.3748/wjg.v19.i6.790 | Embase |
| Systematic Review/ Meta-Analysis | Review: Helicobacter pylori and non-malignant upper gastrointestinal diseases | 2019 | Boltin, D., Niv, Y., Schutte, K. and Schulz, C. | Helicobacter | http://dx.doi.org/10.1111/hel.12637 | Embase |
| Systematic Review/ Meta-Analysis | Barrett's esophagus in 2012: Updates in pathogenesis, treatment, and surveillance | 2013 | Chandra, S., Gorospe, E. C., Leggett, C. L. and Wang, K. K. | Current Gastroenterology Reports | http://dx.doi.org/10.1007/s11894-013-0322-8 | Embase |
| Systematic Review/ Meta-Analysis | Research advances in esophageal diseases: Bench to bedside | 2013 | Di Pietro, M. and Fitzgerald, R. C. | F1000Prime Reports | http://dx.doi.org/10.12703/P5-44 | Embase |
| Systematic Review/ Meta-Analysis | Salivary peptest for laryngopharyngeal reflux and gastroesophageal reflux disease: A systemic review and meta-analysis | 2021 | Guo, Z., Jiang, J., Wu, H., Zhu, J., Zhang, S. and Zhang, C. | Medicine | http://dx.doi.org/10.1097/MD.0000000000026756 | Embase |
| Systematic Review/ Meta-Analysis | Pepsin in saliva as a diagnostic marker for gastroesophageal reflux disease: A meta-analysis | 2018 | Guo, Z., Wu, H., Jiang, J. and Zhang, C. | Medical Science Monitor | http://dx.doi.org/10.12659/MSM.913978 | Embase |
| Systematic Review/ Meta-Analysis | Pepsin: Biomarker, mediator, and therapeutic target for reflux and aspiration | 2018 | Johnston, N., Dettmar, P. W., Ondrey, F. G., Nanchal, R., Lee, S. H. and Bock, J. M. | Annals of the New York Academy of Sciences. | http://dx.doi.org/10.1111/nyas.13729 | Embase |
| Systematic Review/ Meta-Analysis | Biomarkers of Reflux Disease | 2016 | Kia, L., Pandolfino, J. E. and Kahrilas, P. J. | Clinical Gastroenterology and Hepatology | http://dx.doi.org/10.1016/j.cgh.2015.09.014 | Embase |
| Systematic Review/ Meta-Analysis | Oxidative stress in inflammation-based gastrointestinal tract diseases: Challenges and opportunities | 2012 | Kim, Y. J., Kim, E. H. and Hahm, K. B. | Journal of Gastroenterology and Hepatology (Australia) | http://dx.doi.org/10.1111/j.1440-1746.2012.07108.x | Embase |
| Systematic Review/ Meta-Analysis | A Systematic Review of Esophageal MicroRNA Markers for Diagnosis and Monitoring of Barrett's Esophagus | 2016 | Mallick, R., Patnaik, S. K., Wani, S. and Bansal, A. | Digestive Diseases and Sciences | http://dx.doi.org/10.1007/s10620-015-3959-3 | Embase |
| Systematic Review/ Meta-Analysis | Current therapies for gastro-oesophageal reflux in the setting of chronic lung disease: State of the art review | 2020 | Mcdonnell, M. J., Hunt, E. B., Ward, C., Pearson, J. P., O'toole, D., Laffey, J. G., Murphy, D. M. and Rutherford, R. M. | ERJ Open Research | http://dx.doi.org/10.1183/23120541.00190-2019 | Embase |
| Systematic Review/ Meta-Analysis | Early events during neoplastic progression in Barrett's esophagus | 2011 | Reid, B. J. | Cancer Biomarkers | http://dx.doi.org/10.3233/CBM-2011-0162 | Embase |
| Systematic Review/ Meta-Analysis | Review - Helicobacter pylori and non-malignant upper gastro-intestinal diseases | 2020 | Schulz, C. and Kupcinskas, J. | Helicobacter | http://dx.doi.org/10.1111/hel.12738 | PubMed |
| Systematic Review/ Meta-Analysis | Influence of genetics on tumoral pathologies: The example of the adenocarcinoma arising in barrett's esophagus | 2012 | Villanacci, V., Bassotti, G., Salemme, M. and Rossi, E. | Revista Espanola de Enfermedades Digestivas | http://dx.doi.org/10.4321/s1130-01082012001100007 | Embase |
| Study Protocol/ Pilot Study | MicroRNA expression can be a promising strategy for the detection of barrett's esophagus: A pilot study | 2014 | Bansal, A., Hong, X., Lee, I. H., Krishnadath, K. K., Mathur, S. C., Gunewardena, S., Rastogi, A., Sharma, P. and Christenson, L. K. | Clinical and Translational Gastroenterology | http://dx.doi.org/10.1038/ctg.2014.17 | Embase |
| Study Protocol/ Pilot Study | A pilot randomized clinical trial assessing the effect of cricoid pressure on risk of aspiration | 2018 | Bohman, J. K., Kashyap, R., Lee, A., He, Z., Soundar, S., Bolling, L. L. and Kor, D. J. | Clinical Respiratory Journal | http://dx.doi.org/10.1111/crj.12508 | Embase |
| Study Protocol/ Pilot Study | The diagnostic value of pepsin detection in saliva for gastro-esophageal reflux disease: A preliminary study from China | 2017 | Du, X., Wang, F., Hu, Z., Wu, J., Wang, Z., Yan, C., Zhang, C. and Tang, J. | BMC Gastroenterology | http://dx.doi.org/10.1186/s12876-017-0667-9 | Embase |
| Study Protocol/ Pilot Study | Electroacupuncture to treat gastroesophageal reflux disease: study protocol for a randomized controlled trial | 2016 | Han, G., Leem, J., Lee, H. and Lee, J. | Trials | 10.1186/s13063-016-1371-8 | PubMed |
| Study Protocol/ Pilot Study | Impact of tracheal cuff shape on microaspiration of gastric contents in intubated critically ill patients: study protocol for a randomized controlled trial | 2015 | Jaillette, E., Brunin, G., Girault, C., Zerimech, F., Chiche, A., Broucqsault-Dedrie, C., Fayolle, C., Minacori, F., Alves, I., Barrailler, S., Robriquet, L., Tamion, F., Delaporte, E., Thellier, D., Delcourte, C., Duhamel, A. and Nseir, S. | Trials | 10.1186/s13063-015-0955-z | PubMed |
| Study Protocol/ Pilot Study | Barrett's oESophagus trial 3 (BEST3): study protocol for a randomised controlled trial comparing the Cytosponge-TFF3 test with usual care to facilitate the diagnosis of oesophageal pre-cancer in primary care patients with chronic acid reflux | 2018 | Offman, J., Muldrew, B., O'donovan, M., Debiram-Beecham, I., Pesola, F., Kaimi, I., Smith, S. G., Wilson, A., Khan, Z., Lao-Sirieix, P., Aigret, B., Walter, F. M., Rubin, G., Morris, S., Jackson, C., Sasieni, P. and Fitzgerald, R. C. | BMC Cancer | 10.1186/s12885-018-4664-3 | PubMed |
| Study Protocol/ Pilot Study | Exercise and the Prevention of Oesophageal Cancer (EPOC) study protocol: A randomized controlled trial of exercise versus stretching in males with Barrett's oesophagus | 2010 | Winzer, B. M., Paratz, J. D., Reeves, M. M. and Whiteman, D. C. | BMC Cancer | http://dx.doi.org/10.1186/1471-2407-10-292 | Embase |
| In vitro/ cell study | Methylation panel is a diagnostic biomarker for Barrett's oesophagus in endoscopic biopsies and non-endoscopic cytology specimens | 2018 | Chettouh, H., Mowforth, O., Galeano-Dalmau, N., Bezawada, N., Ross-Innes, C., Macrae, S., Debiram-Beecham, I., O'donovan, M. and Fitzgerald, R. C. | Gut | http://dx.doi.org/10.1136/gutjnl-2017-314026 | Embase |
| In vitro/ cell study | Overexpression of CCN1 in Het1A cells attenuates bile-induced esophageal metaplasia through suppressing non-canonical NFkappaB activation | 2019 | Dang, T., Meng, X., Modak, C., Wu, J., Chang, Z., Che, N., Narvaez, R. and Chai, J. | Cytokine | http://dx.doi.org/10.1016/j.cyto.2018.12.020 | Embase |
| In vitro/ cell study | Forkhead box F1 induces columnar phenotype and epithelial-to-mesenchymal transition in esophageal squamous cells to initiate Barrett's like metaplasia | 2021 | De, A., Zhou, J., Liu, P., Huang, M., Gunewardena, S., Mathur, S. C., Christenson, L. K., Sharma, M., Zhang, Q. and Bansal, A. | Laboratory Investigation | http://dx.doi.org/10.1038/s41374-021-00534-4 | Embase |
| In vitro/ cell study | In vitro modelling of barrier impairment associated with gastro-oesophageal reflux disease (Gerd) | 2021 | Meloni, M., Buratti, P., Carriero, F. and Ceriotti, L. | Clinical and Experimental Gastroenterology | http://dx.doi.org/10.2147/CEG.S325346 | Embase |
| Expert opinion/letter to the editor | Barrett's esophagus: Progression to adenocarcinoma and markers | 2011 | Fang, D., Das, K. M., Cao, W., Malhotra, U., Triadafilopoulos, G., Najarian, R. M., Hardie, L. J., Lightdale, C. J., Beales, I. L., Felix, V. N., Schneider, P. M. and Bellizzi, A. M. | Annals of the New York Academy of Sciences | http://dx.doi.org/10.1111/j.1749-6632.2011.06053.x | Embase |
| Expert opinion/letter to the editor | Use of the PEPTEST™ tool for the diagnosis of GERD in the Emergency Department | 2019 | Saviano, A., Petruzziello, C., Brigida, M., Tersigni, I., Migneco, A., Piccioni, A., Saviano, L., Covino, M., Franceschi, F. and Ojetti, V. | Am J Emerg Med | 10.1016/j.ajem.2019.06.047 | PubMed |
| Expert opinion/letter to the editor | Barrett's esophagus: Genetic and cell changes | 2011 | Souza, R. F., Freschi, G., Taddei, A., Ringressi, M. N., Bechi, P., Castiglione, F., Degl'innocenti, D. R., Triadafilopoulos, G., Wang, J. S., Chang, A. C., Barr, H., Bajpai, M., Das, K. M., Schneider, P. M., Krishnadath, K. K., Malhotra, U. and Lynch, J. P. | Annals of the New York Academy of Sciences | http://dx.doi.org/10.1111/j.1749-6632.2011.06043.x | Embase |
| Conference abstract | Pharmacologic treatments for esophageal disorders | 2014 | Blackshaw, L. A., Bordin, D. S., Brock, C., Brokjaer, A., Drewes, A. M., Farmer, A. D., Krarup, A. L., Lottrup, C., Masharova, A. A., Moawad, F. J. and Olesen, A. E. | Annals of the New York Academy of Sciences | http://dx.doi.org/10.1111/nyas.12520 | Embase |
| Case report | Omeprazole induced increase in liver markers-a case report | 2019 | Elmahdy, M. F. and Almater, J. M. | Journal of Clinical and Diagnostic Research | http://dx.doi.org/10.7860/JCDR/2019/41848.13218 | Embase |
| Animal study | High yield reproducible rat model recapitulating human Barrett's carcinogenesis | 2017 | Matsui, D., Omstead, A. N., Kosovec, J. E., Komatsu, Y., Lloyd, E. J., Raphael, H., Kelly, R. J., Zaidi, A. H. and Jobe, B. A. | World J Gastroenterol | 10.3748/wjg.v23.i33.6077 | PubMed |
| **Total Cleaned: 208** | | | | | | |
