## Supplemental Table 7 for "Non-Invasive, MultiOmic and MultiCompartmental Biomarkers of Reflux Disease: A Systematic Review"

| **Supplemental Table­ 7A. Risk of bias assessment for cohort studies (N=6)** | | | | | | | | | |
| --- | --- | --- | --- | --- | --- | --- | --- | --- | --- |
| **Study** | **Selection ^a^** | | | | **Comparability ^b^** | **Outcome ^c^** | | | **Risk of bias** |
|  | **SL1** | **SL2** | **SL3** | **SL4** | **CP** | **OC1** | **^*^OC2** | **OC3** |  |
| **Bor,** 2019 | High | Unclear | Low | Low | Low | Low | N/A | Low | High |
| **Guo,** 2020 | Low | N/A | Low | Low | High | Low | N/A | High | High |
| **De Corso**, 2020 | Low | Low | Low | Low | High | Low | N/A | Low | High |
| **Buas**, 2017 | Low | Low | Low | Low | Low | Low | N/A | Low | Low |
| **Haider,** 2018 | Low | Low | Low | Low | Low | Low | Low | Low | Low |
| **Kim**, 2010 | Low | Low | Low | Low | High | Low | N/A | Unclear | High |
| **Abbreviations:** **SL** Selection ^a^; **CP** Comparability ^b^; **OC** Outcome ^c^. **N/A*** Not applicable.  **SL1**: Representativeness of the exposed cohort (i.e., representative of the average, somewhat representative, specific to a subgroup, or no description of how this was derived in the context of the community of GERD patients being studied)  **SL2**: Selection of the non-exposed cohort (Subjects without Reflux symptoms/ MII-pH event- drawn from the same community as the exposed cohort, drawn form a different source or no description of derivation)  **SL3**: Ascertainment of exposure (Secure record, structured interview or written self-report)  **SL4**: Demonstration that outcome of interest was not present at start of the study (ex. Yes/No if biomarker was present)  **CP**: Comparability of cohorts (on the basis of design or analysis)  **OC1**: Assessment of outcome (Independent blind assessment, record linkage, self-report or no description**)**;  **OC2**: Adequate follow up length (Studies quantified or showed the presence or absence of biomarkers, follow up was not required)  **OC3**: Adequacy of follow up (All subjects accounted for or not) | | | | | | | | | |

| **Supplemental Table­ 7B. Risk of bias assessment for case-control studies (N=9)** | | | | | | | | | |
| --- | --- | --- | --- | --- | --- | --- | --- | --- | --- |
| **Study** | **Selection** ^a^ | | | | **Comparability** ^b^ | **Exposure** ^c^ | | | **Risk of bias** |
|  | **SL1** | **SL2** | **SL3** | **SL4** | **CP** | **EP1** | **EP2** | **EP3** |  |
| **Yuske**l, 2012 | Low | Low | Low | Low | High | Low | Low | Low | High |
| **Wang**, 2019 | Low | Low | Low | Low | High | Low | Low | Low | High |
| **Dettmar**, 2020 | Low | High | Low | Low | High | Low | Low | Low | High |
| **Dryahina**, 2014 | Low | High | Low | Low | Low | Low | Low | Low | High |
| **Snider,** 2018 | Low | High | High | Low | Low | Low | Low | High | High |
| **Yan**, 2016 | Low | Low | Low | Low | Low | Low | Low | Low | Low |
| **Maddalo**, 2018 | Low | Low | Low | Low | High | Low | Low | Low | High |
| **Dosedelova**, 2014 | High | High | Low | High | Low | Low | Low | Low | High |
| **Thrift**, 2014 | Low | Low | High | High | Low | Low | Low | Low | High |
| **Abbreviations:** **SL** Selection ^a^; **CP** Comparability ^b^; **EP** Exposure ^c^  **SL1**: Adequate definition of cases (Yes with independent validation, record linkage or self-report or no description);  **SL2**: Representativeness of the cases (consecutive or obviously representative series of cases, potential for selection bias stated or not stated)  **SL3**: Selection of controls (community controls, hospital controls or no description;  **SL4**: Definition of controls (no history of disease or no description)  **CP**: Control or adjustment for important factors (comparability of cases and controls on the basis of design or analysis)  **EP1**: Ascertainment of exposure (secure record, structured interview blinded/not blinded, written self-reports or no description);  **EP2**: Same method of ascertainment for cases and controls; (Yes or No)  **EP3**: Non-response rate (same rate, non-respondents described, different rate or no designation). | | | | | | | | | |
